# Genomic analysis of intracranial and subcortical brain volumes yields polygenic scores accounting for variation across ancestries

**DOI:** 10.1101/2024.08.13.24311922

**Authors:** Luis M García-Marín, Adrian I Campos, Santiago Diaz-Torres, Jill A Rabinowitz, Zuriel Ceja, Brittany L Mitchell, Katrina L Grasby, Jackson G Thorp, Ingrid Agartz, Saud Alhusaini, David Ames, Philippe Amouyel, Ole A Andreassen, Konstantinos Arfanakis, Alejandro Arias Vasquez, Nicola J Armstrong, Lavinia Athanasiu, Mark E Bastin, Alexa S Beiser, David A Bennett, Joshua C Bis, Marco PM Boks, Dorret I Boomsma, Henry Brodaty, Rachel M Brouwer, Jan K Buitelaar, Ralph Burkhardt, Wiepke Cahn, Vince D. Calhoun, Owen T Carmichael, Mallar Chakravarty, Qiang Chen, Christopher R. K. Ching, Sven Cichon, Benedicto Crespo-Facorro, Fabrice Crivello, Anders M Dale, George Davey Smith, Eco JC de Geus, Philip L. De Jager, Greig I de Zubicaray, Stéphanie Debette, Charles DeCarli, Chantal Depondt, Sylvane Desrivières, Srdjan Djurovic, Stefan Ehrlich, Susanne Erk, Thomas Espeseth, Guillén Fernández, Irina Filippi, Simon E Fisher, Debra A Fleischman, Evan Fletcher, Myriam Fornage, Andreas J Forstner, Clyde Francks, Barbara Franke, Tian Ge, Aaron L Goldman, Hans J Grabe, Robert C Green, Oliver Grimm, Nynke A Groenewold, Oliver Gruber, Vilmundur Gudnason, Asta K Håberg, Unn K Haukvik, Andreas Heinz, Derrek P Hibar, Saima Hilal, Jayandra J Himali, Beng-Choon Ho, David F Hoehn, Pieter J Hoekstra, Edith Hofer, Wolfgang Hoffmann, Avram J Holmes, Georg Homuth, Norbert Hosten, M. Kamran Ikram, Jonathan C Ipser, Clifford R Jack, Neda Jahanshad, Erik G Jönsson, Rene S Kahn, Ryota Kanai, Marieke Klein, Maria J Knol, Lenore J Launer, Stephen M Lawrie, Stephanie Le Hellard, Phil H Lee, Hervé Lemaître, Shuo Li, David CM Liewald, Honghuang Lin, W T Longstreth, Oscar L Lopez, Michelle Luciano, Pauline Maillard, Andre F Marquand, Nicholas G Martin, Jean-Luc Martinot, Karen A Mather, Venkata S Mattay, Katie L McMahon, Patrizia Mecocci, Ingrid Melle, Andreas Meyer-Lindenberg, Nazanin Mirza-Schreiber, Yuri Milaneschi, Thomas H Mosley, Thomas W Mühleisen, Bertram Müller-Myhsok, Susana Muñoz Maniega, Matthias Nauck, Kwangsik Nho, Wiro J Niessen, Markus M Nöthen, Paul A Nyquist, Jaap Oosterlaan, Massimo Pandolfo, Tomas Paus, Zdenka Pausova, Brenda WJH Penninx, G. Bruce Pike, Bruce M Psaty, Benno Pütz, Simone Reppermund, Marcella D Rietschel, Shannon L Risacher, Nina Romanczuk-Seiferth, Rafael Romero-Garcia, Gennady V Roshchupkin, Jerome I Rotter, Perminder S Sachdev, Philipp G Sämann, Arvin Saremi, Muralidharan Sargurupremraj, Andrew J Saykin, Lianne Schmaal, Helena Schmidt, Reinhold Schmidt, Peter R Schofield, Markus Scholz, Gunter Schumann, Emanuel Schwarz, Li Shen, Jean Shin, Sanjay M Sisodiya, Albert V Smith, Jordan W Smoller, Hilkka S Soininen, Vidar M Steen, Dan J Stein, Jason L Stein, Sophia I Thomopoulos, Arthur W. Toga, Diana Tordesillas-Gutiérrez, Julian N Trollor, Maria C Valdes-Hernandez, Dennis van ’t Ent, Hans van Bokhoven, Dennis van der Meer, Nic JA van der Wee, Javier Vázquez-Bourgon, Dick J Veltman, Meike W Vernooij, Arno Villringer, Louis N Vinke, Henry Völzke, Henrik Walter, Joanna M Wardlaw, Daniel R Weinberger, Michael W Weiner, Wei Wen, Lars T Westlye, Eric Westman, Tonya White, A. Veronica Witte, Christiane Wolf, Jingyun Yang, Marcel P Zwiers, M Arfan Ikram, Sudha Seshadri, Paul M Thompson, Claudia L Satizabal, Sarah E Medland, Miguel E Rentería

## Abstract

Subcortical brain structures are involved in developmental, psychiatric and neurological disorders. We performed GWAS meta-analyses of intracranial and nine subcortical brain volumes (brainstem, caudate nucleus, putamen, hippocampus, globus pallidus, thalamus, nucleus accumbens, amygdala and, for the first time, the ventral diencephalon) in 74,898 participants of European ancestry. We identified 254 independent loci associated with these brain volumes, explaining up to 35% of phenotypic variance. We observed gene expression in specific neural cell types across differentiation time points, including genes involved in intracellular signalling and brain ageing-related processes. Polygenic scores for brain volumes showed predictive ability when applied to individuals of diverse ancestries. We observed causal genetic effects of brain volumes with Parkinson’s disease and ADHD. Findings implicate specific gene expression patterns in brain development and genetic variants in comorbid neuropsychiatric disorders, which could point to a brain substrate and region of action for risk genes implicated in brain diseases.

## Introduction

Subcortical brain structures are affected in most major neurological diseases, including psychiatric and developmental brain disorders^1^. These brain structures are involved in crucial daily functions, such as learning^2,3^, memory^3,4^, attention^3^, motor control^2,3^, and reward^5,6^. Likewise, intracranial volume (ICV) variation has been associated with neuropsychiatric phenotypes in observational^7,8^ and genetic^9–11^ studies. Notably, genome-wide association studies (GWAS) have revealed a shared genetic aetiology between brain structures and behavioural, neuropsychiatric, and other health-related phenotypes^2,12–15^.

While neuroimaging genetic studies have advanced our understanding of the genetic architecture of subcortical^2,16^ and cortical^13,17^ brain structures, the most highly powered studies have uncovered the genetic underpinnings of the global measures of the cortex and specific cortical brain structures^13,18,19^. Therefore, there is a need to leverage large and diverse datasets to uncover genetic variants that provide insights into the mechanistic pathways responsible for variation in the volumes of intracranial and subcortical brain volumes.

We coordinated a worldwide analysis of 49 study samples from 19 countries and conducted the largest international genetic analysis of human subcortical brain volumes and ICV. We analysed individual and summary-level genetic data from participants across four international sources to accomplish three goals. First, we sought to characterise the genetic and molecular underpinnings of intracranial and nine subcortical brain volumes (i.e. the brainstem, caudate nucleus, putamen, hippocampus, globus pallidus, thalamus, nucleus accumbens, amygdala and, for the first time, the ventral diencephalon). We performed GWAS meta-analyses including over 70,000 individuals, investigated the genetic overlap among these structural brain volumes, and conducted gene-based tests, eQTL mapping with transcriptome-wide association studies, and the integration of single-cell RNA sequencing data with GWAS summary statistics. Second, we evaluated the predictive utility of polygenic scores for these brain volumes in a diverse ancestral population. Finally, we investigated the overlap and potential causal genetic effects between the observed brain-associated genomic loci and genomic markers implicated in major neurological and psychiatric diseases to examine structure-specific genetic associations with major brain diseases.

This work is crucial, as it can point to a brain substrate and region of action for risk genes implicated in brain diseases.

## Results

### Genome-wide association analyses

We identified 529 genome-wide significant loci (*p*-value < 5×10^−8^) associated with human intracranial or subcortical brain volumes (**Table 1 and Supplementary Figures 1 - 20**), of which 367 survived a multiple testing correction for the total number of phenotypes (*p*-value < 6.25×10^−9^). Of the 529 genome-wide significant loci (**Supplementary Tables 1 and 2**), 254 were independent unique loci across structures (**Supplementary Table 3**). Brainstem volume showed the largest number of independent genetic associations, whereas the amygdala volume had the fewest (**Figure 1** and **Table 1**). SNP-based heritability estimates indicated that common genetic variants explained a substantial proportion of the phenotypic variation of intracranial and subcortical brain volumes, ranging from 17% for the volume of the amygdala to 35% for the volume of the brainstem (**Table 1**). LD score regression intercepts close to or equal to 1 suggested that the elevated lambdas and inflation in the quantile plots (**Supplementary Figures 1 - 20**) were most likely due to polygenicity rather than population stratification (**Table 1**). Attenuation ratios close to 0 indicated correct genomic control. Manhattan and QQ plots for GWAS in individual cohorts are available in **Supplementary Figures 21 - 60**.

**Table 1.** Summary of GWAS meta-analysis results per subcortical brain volume and intracranial volume (ICV).

| Brain volume | Number of genome-wide significant loci | h<sup>2</sup>SNP (SE) | Intercept (SE) | Attenuation ratio (SE) |
| --- | --- | --- | --- | --- |
| Brainstem | 96 | 0.35 (0.03) | 1.00 (0.01) | < 0 |
| ICV | 83 | 0.28 (0.02) | 1.00 (0.02) | 0.006 (0.04) |
| Caudate nucleus | 78 | 0.27 (0.02) | 1.00 (0.01) | 0.001 (0.03) |
| Putamen | 71 | 0.29 (0.03) | 1.00 (0.01) | 0.006 (0.03) |
| Hippocampus | 47 | 0.21 (0.02) | 1.00 (0.01) | 0.001 (0.04) |
| Ventral diencephalon | 36 | 0.33 (0.03) | 1.00 (0.01) | 0.01 (0.04) |
| Thalamus | 35 | 0.22 (0.01) | 1.00 (0.01) | < 0 |
| Globus pallidus | 32 | 0.22 (0.02) | 1.00 (0.01) | < 0 |
| Nucleus accumbens | 29 | 0.21 (0.01) | 1.00 (0.01) | 0.003 (0.03) |
| Amygdala | 22 | 0.17 (0.01) | 1.00 (0.01) | < 0 |
| Total | 529 | NA | NA | NA |
| Number of genome-wide loci is reported at the common genome-wide significance threshold ($p$-value < $5 \times 10^{-8}$, $r^2$ threshold to define independent significant loci $\geq 0.6$, second $r^2$ threshold to define lead loci $\geq 0.05$). A full list of independent significant loci is reported in Supplementary Table 1. | | | | |

**Figure 1.**
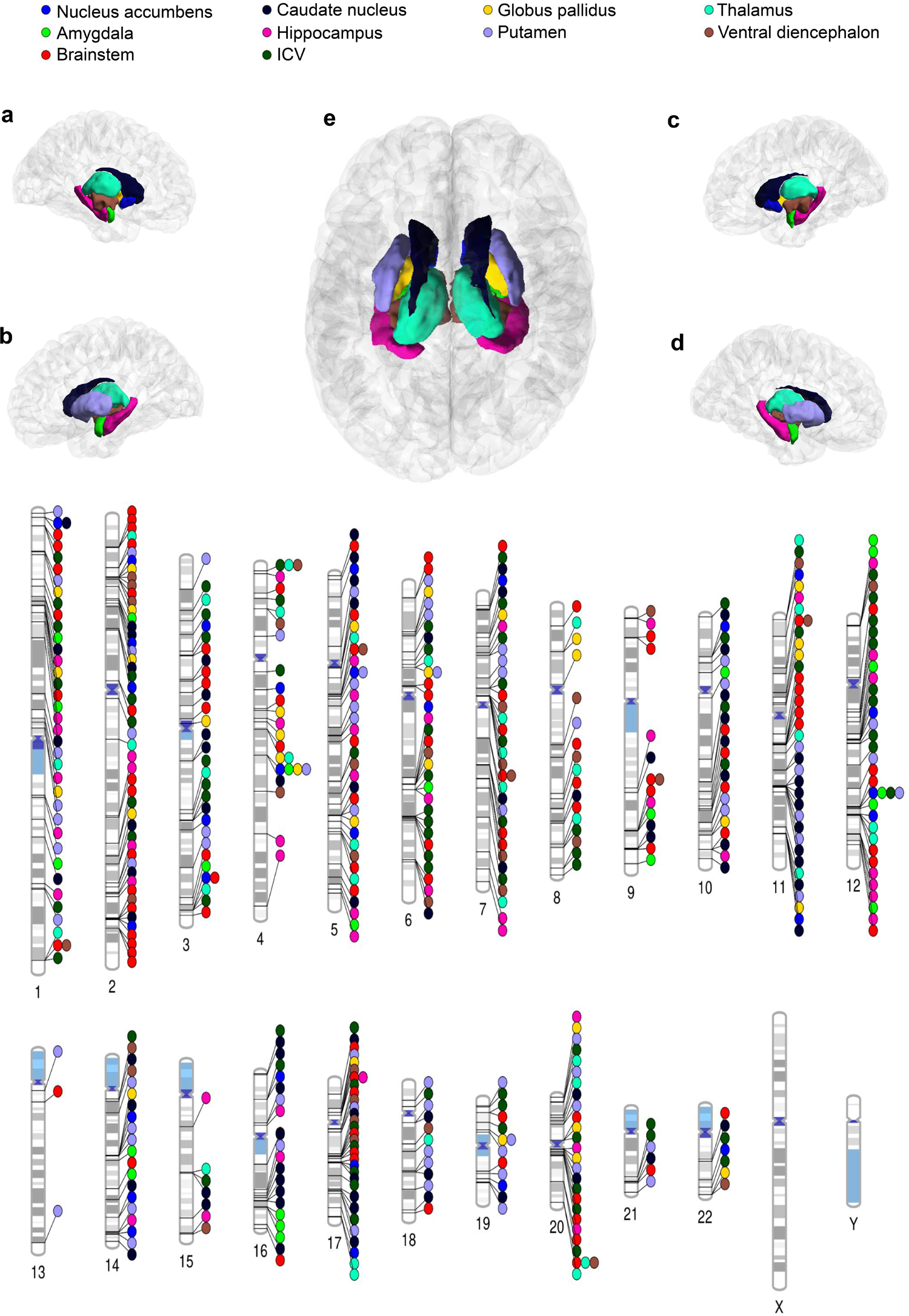
Meta-analyses results overview. Phenogram illustrating loci associated with each of the brain volumes under study at the common genome-wide significance threshold (*p*-value < 5×10^−8^). ICV = Intracranial volume, A = Left Hemisphere interior, B = Left Hemisphere exterior, C = Right hemisphere interior, D = Right hemisphere exterior, E = Both hemispheres upper. The p-values referenced here correspond to a two-tailed Z-test test as implemented in MTAG.

As a sensitivity analysis, we performed GWAS in the UK Biobank cohort for subcortical brain volumes without adjusting for ICV (**see Methods and Supplementary Figures 61 - 78**.). Direction and magnitude of SNP effect sizes were largely consistent as suggested by Pearson’s correlations using the SNP effect sizes for the same phenotype with and without the adjusting for ICV (correlations range = 0.81 - 0.92). Moreover, we split the UK Biobank sample into two randomised subsamples (N ∼ 18,047 each) in an attempt to investigate replicability for intracranial and subcortical brain volumes (**Supplementary Figures 79 - 118)**. Direction and magnitude of SNP effect sizes were for the most part consistent as suggested by Pearson’s correlations using the effect sizes for the same phenotype for both subsamples (correlations range = 0.67 - 0.84).

### Functional annotation and gene prioritisation

We used MAGMA (v1.08) to perform gene-based association analyses. GWAS meta-analysis for ICV and the volumes of the brainstem and caudate nucleus showed the largest number of genes associated with each structure, followed by the volumes of the putamen, hippocampus, ventral diencephalon, globus pallidus, thalamus, and nucleus accumbens (**Supplementary Table 4**). Amygdala volume was associated with the fewest genes. No single gene was associated with all intracranial or subcortical brain volumes, which reflects the correction for ICV. The Forkhead Box O3 (*FOXO3*) gene was associated with the volume of five brain structures. Similarly, the Geminin Coiled-Coil Domain Containing (*GMNC*), A-kinase anchoring protein 10 (*AKAP10*), Epidermal Growth Factor Receptor (*EGFR*), Microtubule Nucleation Factor (*TPX2*), and the Bcl-2-like Protein 1 (*BCL2L1*) were associated with the volume of four brain structures. Furthermore, genes from the *HOX* and *PAX* homeobox gene families were associated with the volume of the brainstem. In addition, genes from the *WNT* family were associated with brainstem, ventral diencephalon and intracranial volumes. Other genes associated with multiple subcortical brain volumes included *BIRC6, CRHR1, IGF1, MAPT, NUP37, NUP43, KTN1, FOXS1* and *COX4I2*, which have been previously reported to have roles in intracellular signalling^20^, autophagy^21–23^, and multiple brain ageing processes, such as vascular ageing, oxidative resistance, tau pathology, and apoptosis^24–27^. A full list of statistically significant gene-based test findings after Bonferroni multiple testing correction is available in **Supplementary Table 5**.

We integrated our GWAS results with expression quantitative trait loci (eQTL) data from the Genotype-Tissue Expression project (GTEx/v8) (**Supplementary Table 6**). We observed consistent findings with our gene-based tests. The genes *CRHR1, NUP43*, and *KTN1* were associated with subcortical brain volumes. Furthermore, we observed associations for the genes *UQCC1*, and *COX4I2*. Genes that may be linked to specific brain structures through changes in gene expression include, among others, *CRHR1* for the putamen, *FAIM* for the thalamus, and *MAPK3* as well as *ZNF786* for the hippocampus. We prioritised potential causal genes from the associated loci performing transcriptome-wide association studies (TWAS). Most genes were associated uniquely with the volume of a single brain structure (91%), while others were shared across the volumes of several brain structures. With this approach we observed associations of the genes *CRHR1, MAPT, NUP43, NUDT14, FAIM, MAPK3, and ZNF786* with subcortical brain volumes (**Supplementary Table 6**), even after correcting for multiple testing using a conservative approach (*p*-value < 3.06×10^−4^, **see Methods**). Likewise, we revised eQTLs in developmental datasets to identify genes involved in brain development (**Supplementary Table 7**) and observed associations of brainstem, caudate nucleus, putamen, thalamus, ventral diencephalon, and intracranial volumes with the genes *LRRC37A, LRRC37A2, KANSL1, RPS26, ARL17B, PILRB, PILRA*, and *EFCAB13* after correcting for multiple testing using a conservative approach (*p*-value < 1.26×10^−3^, **see Methods**).

We integrated single-cell RNA-sequencing data^28^ with GWAS summary statistics to identify critical cell types and cellular processes influencing intracranial and subcortical brain volumes variation. From the prioritised genes across MAGMA and TWAS analyses, we identified nine expressed genes (*TUFM, CRHR1, NUP43, MAPK3, LRRC37A2, FAIM, ZNF786, YIPF4* and *PSMC3*) across seven different cell types, including pluripotent floor progenitor plate cells (FPP), proliferating floor progenitor plate cells (P_FPP), dopaminergic neurons (DA), ependymal-like 1 (Epen1), serotonergic-like neurons (Serts), and astrocyte-like cells (Astro), influencing brain volume variation. Our gene expression findings in cell types mentioned previously cover up to 52 days of differentiation. Most of the expressed genes at day 11 were observed in FPP and P_FPP; at day 30 in FPP, DA, and ependymal-like 1 cells; and at day 52 in DA, serotonergic-like neurons, astrocyte-like, ependymal-like 1 cells. Full results surviving multiple testing correction (p-value < 1.19×10^−3^, **see Methods**) are available in **Supplementary Table 8**.

As a sensitivity analysis, we performed MAGMA analyses for subcortical brain volumes using data from the UK Biobank with and without adjusting for ICV (**Supplementary Tables 9 and 10**). Identified genes were consistent with and without the adjustment for ICV. However, these genes were associated with more subcortical brain volumes when GWAS were not adjusted for ICV.

### Polygenic scores predict phenotypic brain volumes

We tested the predictive capability of our genome-wide results by performing the meta-analyses leaving out the ABCD cohort (N = 5,267) to determine whether polygenic scores from European ancestry samples are associated with intracranial and subcortical brain volumes in the more diverse ABCD cohort. The polygenic scores for all brain volumes were strongly associated with intracranial and subcortical volumes in participants of European, African, and non-European ancestries, as well as across all ancestral groups (**Figure 2** and **Supplementary Figures 119-124**). Overall, results remained consistent with additional adjustments for cryptic relatedness. While polygenic prediction was most accurate for participants of European ancestry (variance explained ranging from 2.1 to 8.5%), we observed that the variance explained in non-European ancestry groups was also significant and ranged from 0.8 to 9.8% (**Figure 2** and **Supplementary Table 11**). Sensitivity analyses included linear regressions among participants of European ancestry for subcortical volumes using ICV as a covariate. The results were consistent and remained essentially unchanged. As expected, polygenic scores for ICV did not explain residual variance above phenotypic ICV (**Supplementary Table 12, Supplementary Table 13**, and **Supplementary Figure 125**).

**Figure 2.**
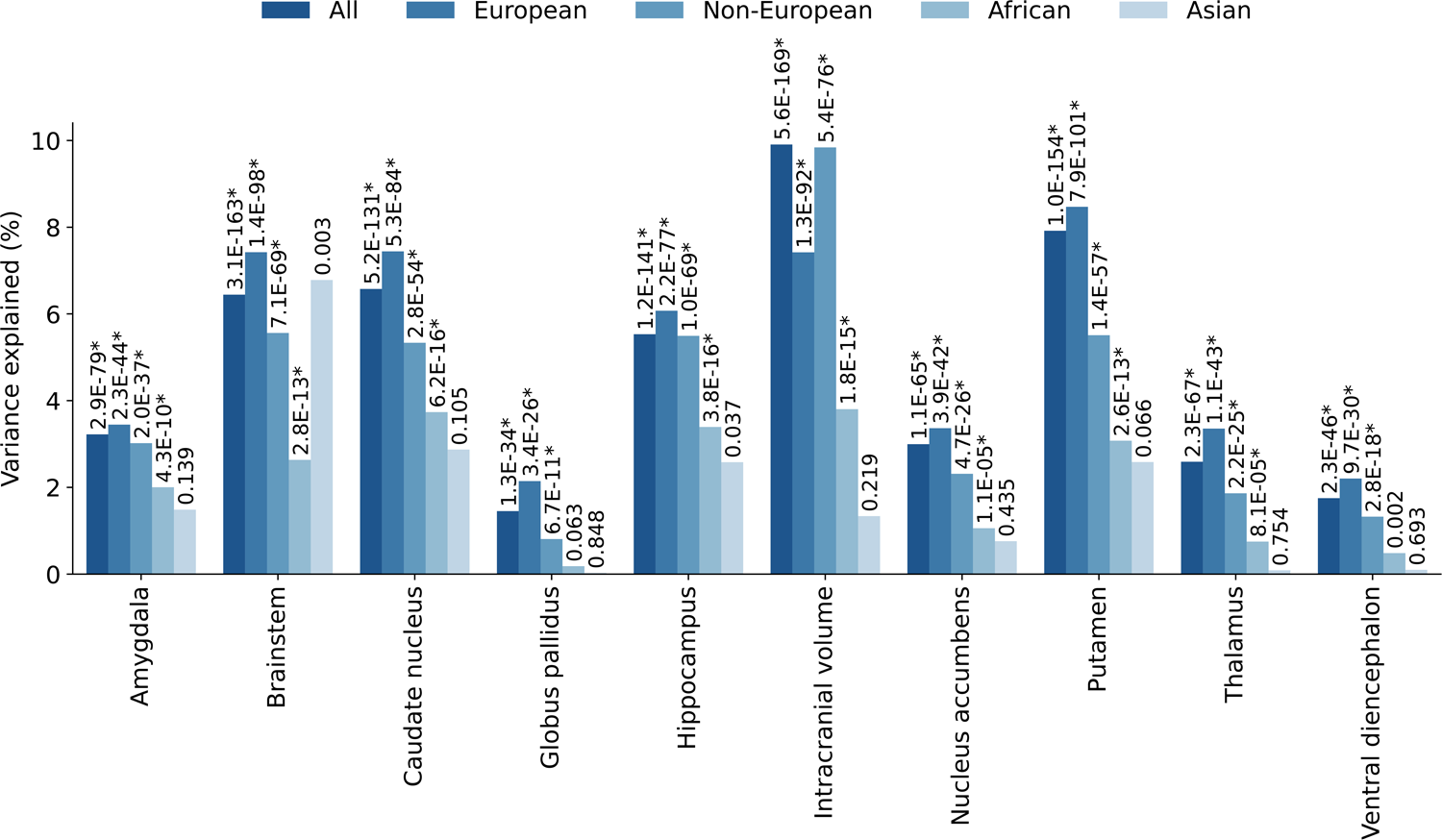
Polygenic prediction in the ABCD cohort. Barplots show the variance explained by intracranial and subcortical brain volume polygenic scores using the *SBayesR* approach with a linear mixed effects model implemented in GCTA for the whole sample (N = 10,440), and individuals of European (N = 5,267), Non-European (N = 5,173), African-only (N = 1,833), and Asian-only (N = 152) ancestry. The *p*-value of the association is shown at the top of each bar; those with an asterisk (*) were significant after Bonferroni multiple testing correction [0.05 / 50 [total number of tests] = 1×10^−3^]. Non-European ancestry individuals include, but are not limited to, African-only and Asian-only ancestries as individuals with admixed ancestry were also included. P-values in this figure correspond to wald-tests (2-sided) derived from the linear mixed model results.

### Genetic overlap between subcortical brain structures

Using LD score regression, we estimated genetic correlations among intracranial and the nine subcortical brain volumes under study. We adopted a conservative approach to multiple testing and corrected for the total number of genetic correlations, including those for other complex human phenotypes [0.05 / 320 [total number of genetic correlation tests] = 1.56×10^−4^]. We observed substantial genetic overlap among intracranial and subcortical brain volumes (**Figure 3** and **Supplementary Tables 14 - 15**). The thalamus volume showed genetic correlations with the other eight brain volumes. The volume of the brainstem, amygdala, and the caudate nucleus, with four significant genetic correlations, showed the fewest. Components of the striatum, including the caudate nucleus and putamen, were strongly correlated with the nucleus accumbens. Within-phenotype genetic correlations across cohorts were large (rg > 0.60) and statistically significant after multiple testing correction (**Supplementary Table 16**).

**Figure 3.**
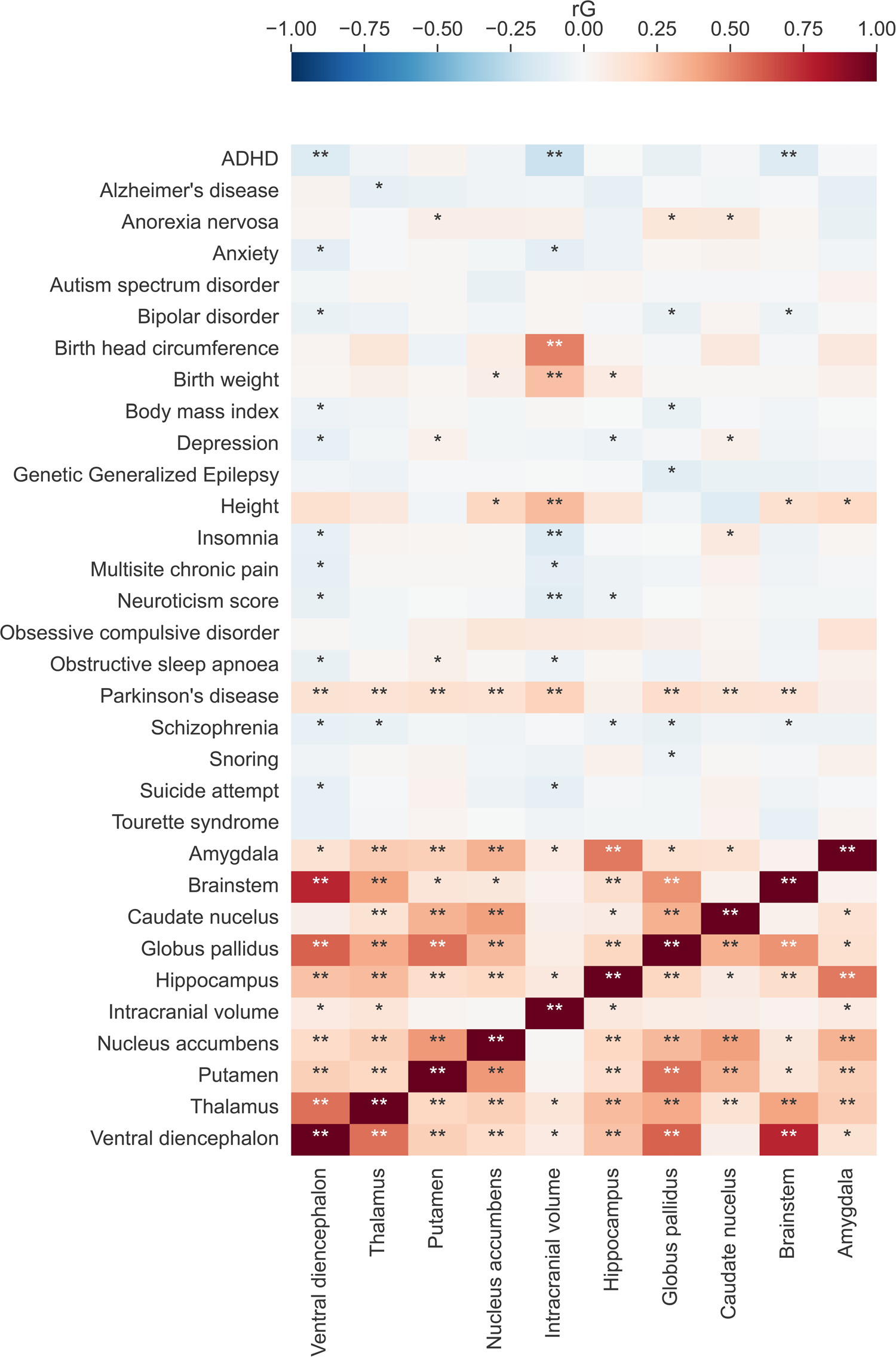
Genetic overlap with neuropsychiatric traits and disorders. Heatmap depicting genetic correlations (rG) of intracranial and subcortical brain volumes with complex human phenotypes. \**p*-value < 0.05; \*\**p*-value significant after Bonferroni multiple testing correction (0.05 / 320 [total number of genetic correlation tests] = 1.56×10^−4^). Genetic correlations were estimated using LD score regression. P-values correspond to chi-squared tests with one degree of freedom as implemented in LD score regression.

We further explored polygenic overlap between the GWAS summary statistics, including all cohorts, for the subcortical brain volumes using MiXeR. We estimated the number of causal variants influencing each subcortical brain volume (median n causal variants = 1.92K, **Supplementary Table 17**). The volume of the hippocampus was the least polygenic (1.000K causal variants, SE = 0.13K), whilst thalamus volume was the most polygenic (2.58K causal variants, SE = 0.15K). We then estimated the number of causal variants shared between subcortical brain volumes, finding substantial polygenic overlap between them (median n shared causal variants = 1.24K, **Supplementary Table 18**). The largest overlap was observed between the volumes of the thalamus and globus pallidus (2.08K variants, SE = 0.22K), while the smallest was between the thalamus and hippocampus (0.53K variants, SE = 0.04K). We identified polygenic overlap between three pairs of brain structures: brainstem-amygdala (0.97K variants, SE = 0.09K), brainstem-caudate nucleus (0.87K variants, SE = 0.10K), and caudate-ventral diencephalon (1.01K variants, SE = 0.12K), despite their genetic correlation being close to zero (**Figure 3**).

### Genetic clustering of subcortical brain structures

We used genomic structural equation modelling (SEM)^29^ to examine whether and how subcortical brain structures cluster together at a genetic level. We first tested a common factor model, which provided a poor fit to the data (CFI = 0.70, SRMR = 0.13, AIC = 828.03; **Supplementary Table 19**). To explore other possible factor structures underlying subcortical brain structures, we conducted genetic exploratory factor analyses (EFA) based on the genetic correlation matrix of the 9 subcortical structures (**Supplementary Table 20**). A two-factor model (**Supplementary Table 21**) and three-factor model (**Supplementary Table 22**) explained 43% and 53% of the total genetic variance, respectively. Follow up confirmatory factor analyses (CFA) were specified in genomic SEM (retaining standardised loadings greater than 0.25). While the two-factor model did not provide adequate fit (CFI = 0.84, SRMR = 0.09, AIC = 482.97), the three-factor model provided good fit to the data (CFI = 0.91, SRMR = 0.06, AIC = 299.33, **Figure 4**).

**Figure 4.**
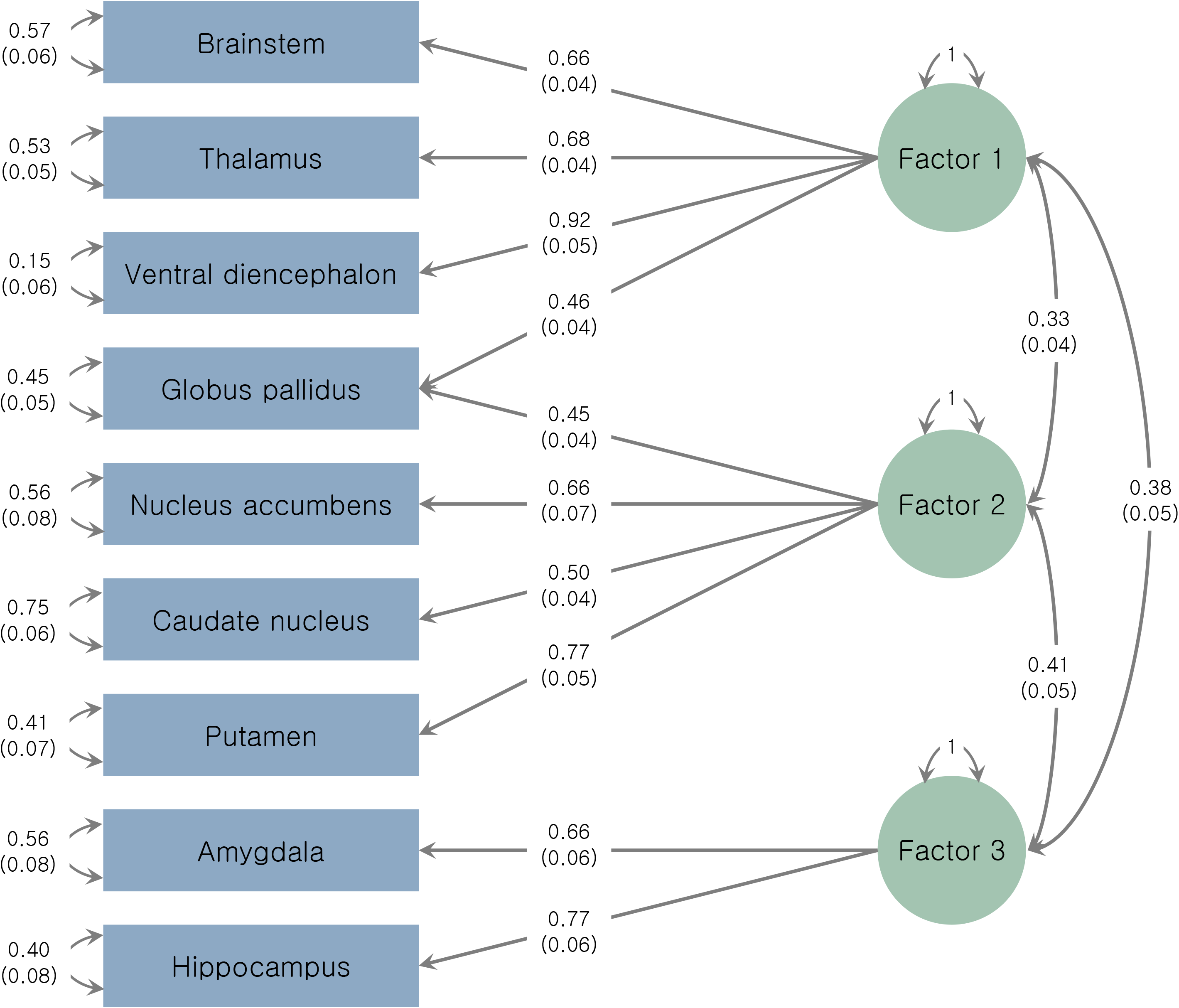
Genetic structure of subcortical brain volumes. Path diagram of a three-factor model estimated with genomic structural equation modelling. Blue rectangles represent the genetic component of each subcortical brain volume. Green circles represent latent factors. Standardised path coefficients are presented.

### Genetic correlations with brain disorders

We estimated genetic correlations between the brain volumes investigated and 22 complex human phenotypes (**Figure 3** and **Supplementary Tables 14 - 15**). Parkinson’s disease, attention-deficit/hyperactivity disorder (ADHD), neuroticism score, birth weight, birth head circumference, height, and insomnia showed statistically significant associations after correction for multiple testing. Parkinson’s disease showed several positive genetic correlations with intracranial and subcortical brain volumes, including those of the nucleus accumbens, brainstem, caudate nucleus, globus pallidus, putamen, thalamus, and ventral diencephalon. We observed negative genetic overlap for ICV with ADHD, insomnia, and neuroticism scores. Conversely, we identified a positive genetic correlation of birth weight, birth head circumference, and height with ICV.

We further investigated the relationship between brain volumes and complex human phenotypes with a statistically significant genetic correlation using the pairwise-GWAS (GWAS-PW) method. With this approach, we identified 338 genomic segments with genetic variants influencing both the volume of a brain structure and a human complex phenotype (**Supplementary Table 23**). Genomic segments with shared genetic variants were identified for all traits that displayed a significant genetic correlation after multiple testing correction, except for the ventral diencephalon and ADHD.

As a sensitivity analysis, we investigated whether adjusting or not adjusting for ICV had an effect on genetic correlations with complex human phenotypes. We used the GWAS for subcortical brain volumes from the UK Biobank with and without adjusting for ICV and estimated genetic correlations with complex human traits. We observed more statistically significant genetic correlations with complex human phenotypes when not adjusting subcortical brain volumes for ICV. However, the direction and magnitude of the genetic correlations remained for the most part consistent regardless of the adjustment for ICV (**Supplementary Tables 24 - 29; Supplementary Figures 126 and 127**).

### Potential causal genetic effects

We estimated the genetic causal proportion (GCP) with the latent causal variable method (LCV) and leveraged the Latent Heritable Confounder Mendelian Randomisation (LHC-MR) method to assess potential causal genetic effects of intracranial and subcortical brain volumes with complex human traits that displayed a statistically significant genetic correlation after Bonferroni multiple testing correction. We observed putative causal genetic effects for a larger putamen volume influencing a higher risk for Parkinson’s disease after multiple testing correction using the LCV [0.05 / 16 [total number of genetic causal proportion tests in the present study] = 3.13×10^−3^] and LHC-MR [0.05 / 32 [total number of LHC-MR tests in the present study] = 1.56×10^−3^] methods. With both methods, we observed that a larger ICV could reduce the likelihood of developing ADHD. Potential causal genetic effects suggesting that a larger ICV could reduce the likelihood of developing insomnia were observed with LCV, but not with LHC-MR. We observed several potential causal genetics effects of nominal significance (*p*-value < 0.05), which are fully described in **Supplementary Table 30** and **Supplementary Table 31**

## Discussion

We performed the largest GWAS meta-analysis of intracranial and subcortical brain volumes to date across international datasets from 19 countries. Here, we complement and extend work from a previous GWAS meta-analysis that identified 48 significantly associated loci with seven subcortical brain volumes^2^. Our results implicated more than 254 independent genetic variants, at the common genome-wide threshold (*p-value* < 5×10^−8^), associated with ICV or the volumes of the brainstem, caudate nucleus, putamen, hippocampus, globus pallidus, thalamus, nucleus accumbens, amygdala and for the first time, the ventral diencephalon, in over 70,000 individuals. Of these 254 independent genetic variants, 161 have not been reported in previous studies^2,14–16,30^. From the independent genome-wide genetic variants reported in previous studies^2,14–16,30^, we replicated 39% (N = 167) in the genome-wide loci in our meta-analysis at the common genome-wide threshold (*p*-value < 5×10^−8^). Our findings provide insights into genes that influence variation in human intracranial and subcortical brain volumetric measures. We show that distinct genetic variants often have a specific effect on the variation of a single brain volume. In addition, we conducted thorough functional annotation and gene prioritisation analyses, including gene-based tests, TWAS, and the integration of single-cell RNA sequencing data with GWAS summary statistics. We investigated the genetic overlap and putative causal genetic effects of intracranial and subcortical brain volumes with other complex human phenotypes. Polygenic scores for intracranial and subcortical volumes showed predictive ability for their corresponding phenotypic measurements, even when examined in a pre-adolescent population with individuals of diverse ancestral backgrounds.

Previous work suggests that heritability estimates for intracranial and subcortical brain volumes range from 33 to 86% in twin and family studies^2,31,32^ and from 9 to 33% using a SNP-based heritability approach^2^. In our study, SNP-based heritability estimates derived from GWAS meta-analysis results ranged from 18 to 38%. These values are consistent with prior findings in the UK Biobank and ENIGMA cohorts^2,33^. Furthermore, a previous GWAS meta-analysis of ICV identified 64 genetic variants explaining 5% of phenotypic variation in a sample of European ancestry^14^. In the present study, we explained 28% (CI = 26 - 30%) of phenotypic variation and identified 83 significant loci associated with ICV at the common genome-wide threshold (*p*-value < 5×10^−8^).

We explored genetic correlations among intracranial and subcortical brain volumes, including the first ever findings for the ventral diencephalon. We identified substantial genetic overlap for these brain volumes, consistent with previous reports, and supporting previously observed phenotypic associations^2^. In contrast with previous findings^2^, we identified several genetic correlations of subcortical brain volumes, including the hippocampus, globus pallidus, thalamus and ventral diencephalon, with the brainstem. The strongest genetic correlation among all brain volumes was observed between the volumes of brainstem and the ventral diencephalon. This finding is consistent with brainstem anatomy and the interconnection with the ventral diencephalon, as the brainstem can be subdivided into the diencephalon (thalamus, hypothalamus), mesencephalon (midbrain), ventral metencephalon (pons), and myelencephalon (medulla)^34^. In addition, with genomic SEM analyses we observed how subcortical brain structures cluster together at a genetic level. The volumes of the nucleus accumbens, caudate nucleus, putamen and globus pallidus clustered together, which is consistent with the structure of the basal ganglia^35–37^ and the striatum^36^. Furthermore, the volume of the globus pallidus also clustered together with those of structures strongly interconnected with the basal ganglia^35^, such as the brainstem, thalamus, and ventral diencephalon, while the volumes of the amygdala and the hippocampus, whose circuitry in the limbic system is well-known to predominantly influence emotion-regulated memories^38^, constituted the third cluster.

Previous studies have aimed to investigate the genetic overlap of intracranial and subcortical brain volumes with neuropsychiatric disorders^1,2,12,39,40^. Here, we identified genetic correlations for eight subcortical brain volumes with Parkinson’s disease and three with ADHD. ICV showed genetic overlap with both Parkinson’s disease and ADHD. ADHD and Parkinson’s disease are predominantly young- and late-onset phenotypes, respectively^41,42^. However, our GWAS summary statistics for intracranial and subcortical brain volumes do not necessarily include people diagnosed with Parkinson’s disease, ADHD, or individuals at high risk for these disorders. Thus, positive genetic correlations with Parkinson’s disease suggest that genetic variants influencing larger volumes during the development of specific structures are also associated with a higher risk for Parkinson’s disease, consistent with previous observations in genetic studies^2^. In contrast, negative genetic correlations with ADHD imply that genetic variants influencing a smaller volume of specific structures are associated with a higher genetic susceptibility for ADHD^43^. We present, for the first time, the further interrogation of the observed genetic correlations via different methods to demonstrate putative causal genetic effects between a range of subcortical brain volumes and various complex human phenotypes.

Identified loci for intracranial and subcortical brain volumes were annotated using gene-based testing, eQTL mapping, TWAS and the integration of single-cell RNA sequencing data with GWAS summary statistics. Most of the genes associated with intracranial or subcortical brain volumes across analyses were uniquely associated with a specific brain volume, shedding light on the independent genetic underpinnings of these structures. While the remaining genes showed effects influencing more than one brain structure, no single gene was associated with all brain measures assessed. We identified gene expression in different neural cell types for genes that have been previously reported to act through pathways related to autophagy (*TUFM*, and *FAIM*)^21–23^, mediation of intracellular signalling (*MAPK3*)^20^, organelle biogenesis and maintenance (*YIPF4*)^44^, and nucleo-cytoplasmic transport of RNA and proteins (*NUP43*)^45^. Some of the identified expressed genes (*CRHR1*^*24–26*^ and *LRRC37A2*^*46,47*^) have been previously associated with neurodegenerative disorders^48^. For instance, it has been suggested that *CRHR1* may have a neuroprotective effect in Parkinson’s disease^24–26^, and may even prevent dementia-related symptoms^49^.

Previous studies suggest that polygenic scores lack predictive ability on ancestral groups that do not match the ancestry of the discovery GWAS^50^. However, in the present study, we observed for the first time that polygenic scores significantly predicted the same intracranial and subcortical brain volumes in a sample of preadolescent children of European and non-European ancestries. Given that PRS prediction was possible in children, it is likely that the genetic variation underlying differences in adult intracranial and subcortical brain volumes is present at an early age. This is consistent with prior work suggesting that prenatal and postnatal development of subcortical brain regions is influenced by genetic variants associated with subcortical brain volumes in adults^51^. Furthermore, our polygenic scores account for a significant fraction of brain variability across ancestries. This suggests that genetic variants responsible for subcortical brain structure could be shared across ancestries, with linkage disequilibrium and minor allele frequency differences underlying differences in accuracy for trans-ancestry predictions^52^. We observed that predictions on participants of African ancestry outperformed those for participants of Asian ancestry. This is inconsistent with previous studies demonstrating that LD patterns in Asians are more similar to those in Europeans when compared with those of African ancestry. We attribute our observations to the difference in sample sizes, which is larger for participants of African ancestry (N = 1,833) than for those of Asian ancestry (N = 152). Overall, our findings point towards polygenic score generalisability across individuals of diverse ancestral backgrounds, and could be leveraged to study brain development in young populations. Well powered polygenic predictors, will potentially enable to boost power of future neuroimaging GWAS performed in samples of underrepresented ancestries^53^, an important endeavour to narrow the ancestry biases in current genetic studies.

When performing GWAS on any brain measurements, the inclusion of ICV as a covariate in the model is frequently used and widely accepted to adjust for differences in head size among participants^2,13,16,30^. However, this practice remains open for discussion as there is potential for collider bias. Correcting for a heritable, correlated covariate, such as ICV, can bias estimates, which could potentially limit the interpretability of gene-identification and other downstream analyses^54^. In the present study, we performed GWAS for subcortical brain volumes in the UK Biobank cohort with and without adjusting for ICV to investigate potential differences. We estimated genetic correlations with complex human phenotypes and performed gene-based tests. For these analyses, we observed more statistically significant associations for the GWAS that were not adjusted for ICV. We suggest that the effect of ICV is driving these associations. For instance, ICV is correlated with head birth circumference and birth weight. When not adjusting for ICV, most if not all of the subcortical brain volumes were genetically correlated with head birth circumference and birth weight after multiple testing correction. Consistently, when adjusting for ICV, a few subcortical brain volumes were barely genetically correlated with these phenotypes. Similar observations were made for gene-based tests. Lastly, when a correction for ICV is not included in volumetric studies using magnetic resonance imaging, sex differences are observed^55^. We consider that for the analyses of brain size-related measurements, the adjustment for ICV is necessary to account for differences in head size and sex, which will directly influence the measurements. We consider this crucial in our study since we leveraged data from different cohorts, such as ABCD and the UK Biobank, which include participants of different age, sex, and total brain size. Future studies should aim to fully investigate the effect of ICV on neurogenomic analyses.

The limitations of this study must be acknowledged. As we mentioned in the methods section, the imaging analysis and visualisation of structural data in all cohorts was performed using the publicly available FreeSurfer package tool, which includes the superior cerebellar peduncle as part of the brainstem. The superior cerebellar peduncle is a structure that connects the cerebellum to the brainstem^56^. However, anatomically, the cerebellar peduncle is not a putative structure of the brainstem^56^. Therefore, we note that the inclusion of the cerebellar peduncle as part of the volume of the brainstem is a limitation of the segmentation performed by the FreeSurfer package tool, which we are unable to address. Furthermore, our GWAS meta-analyses included only participants of European ancestry in the discovery phase. Therefore, the genetic loci associated with intracranial and subcortical brain volumes in the present study are only representative of individuals of European ancestry until confirmed in samples of other ancestral populations.

We provide evidence for the polygenic architecture of intracranial and subcortical brain volumes, presenting findings for the volume of the ventral diencephalon for the first time, and show that polygenic scores could be useful in predicting or imputing brain volume measures in future studies. Multiple genes were associated with the brain volumes investigated in this, the largest and most geographically diverse genetic study to date. Genes identified were expressed in specific neural cell types that influence intracranial and subcortical brain volumes and are involved in autophagy, intracellular signalling and transport, organelle biogenesis and maintenance, or the aetiology of neurodegenerative disorders. Our findings point towards the generalisability of intracranial and subcortical brain volumes’ polygenic scores to non-European ancestry individuals, suggesting a shared genetic basis of these brain volumes across diverse ancestral groups. We observed genetic overlap and putative causal genetic effects of intracranial and subcortical brain volumes with neuropsychiatric conditions, including Parkinson’s disease and ADHD. Overall, our findings advance the understanding of the brain’s complex and polygenic genetic architecture, implicating multiple molecular pathways in human brain structure and suggesting that multiple genetic variants of small effect size are likely to be involved in the development of specific brain volumes. These studies also facilitate our understanding of shared genetic pathways underlying the aetiology of brain disorders and the formation and adaptation of the human brain.

## Data Availability

Detailed information on how to access publicly available GWAS summary data from the ENIGMA and CHARGE consortia is reported on their corresponding publications2,12,15. Researchers can access individual-level data from the UKB and ABCD cohorts following the corresponding data application procedures. Work performed using UKB data was done under application 25331. Full genome-wide summary statistics generated in the present study are available at the ENIGMA website (https://enigma.ini.usc.edu/research/download-enigma-gwas-results).

## Acknowledgments

We thank all of the study participants for contributing to this research. Full acknowledgements and grant support details are provided in the Supplementary Note.

## Author contributions

### Core analysis and writing group

LMGM, AIC, SDT, JAR, ZC, BLM, KLG, JGT, PMT, CLS, SEM, MER.

### Cohort Pis

IA, DA, OAA, AA, DAB, MPMB, DIB, HB, JKB, WC, VC, SC, BC, AMD, GIdZ, CDeC, CDep, SDes, SEh, TE, SEF, MF, BF, HJG, OG, VG, AKH, UKH, AH, SH, PJH, MI, MAI, EGJ, RSK, LJL, SML, HLe, PMe, AM, THM, MMN, NGM, PAN, JO, TP, ZP, BWJHP, BMP, PSS, PGS, AJS, RS, GS, SS, JWS, DJS, JNT, DvE, HvB, NJAv, DJV, MWV, AV, HW, DRW, MWW, TW, AW.

### Imaging data collection

IA, SA, KA, MEB, ASB, VC, BC, FC, EJCdG, CDeC, SEr, TE, GF, IF, DAF, ALG, OGr, OG, VG, AKH, UKH, SH, BH, AJH, NH, MAI, CJJr, EGJ, RSK, SML, HLe, DCML, JM, VSM, KLM, PMe, THM, TWM, SM, PAN, JO, MP, BWJHP, GP, NR, PGS, GS, SS, SMS, HSS, DTT, JNT, MCV, DvE, NJAv, MWV, HW, JMW, MWW, WW, LTW, EW, TW, MPZ.

### Contributed to the editing of the manuscript

IA, DA, JCB, HB, JKB, JGT, VC, CRKC, GD, EJCdG, PL.DJ, SDes, SEh, TE, GF, IF, SEF, AJF, CF, BF, HJG, KLG, NAG, OG, UKH, DPH, SH, JJH, NH, MAI, JCI, NJ, MJK, SML, PHL, HLe, HL, WTL, ML, AFM, KAM, VSM, AM, BLM, THM, WJN, MMN, PAN, TP, BWJHP, BMP, JIR, PSS, CLS, SIT, AJS, GS, ES, SS, LSh, SMS, HSS, DJS, JLS, SIT, PMT, AT, JNT, MWV, JMW, DRW, MWW, LTW, TW, NMS, NGM, JR, LMGM, AIC, MER.

### Genetic data collection

SA, PA, LA, RB, VC, SC, EJCdG, PL.DJ, CDep, SDes, SD, SEr, TE, SEF, AJF, CF, RCG, OG, VG, AKH, BH, GH, MAI, EGJ, SLH, DCML, JM, KAM, PMe, THM, TWM, MMN, PAN, BWJHP, BMP, BP, MDR, JIR, AJS, PRS, MSc, SS, LSh, SMS, HSS, VMS, NJAv, JV, HW.

### Imaging data analysis

SA, KA, RMB, OTC, MC, QC, CRKC, BC, FC, CDeC, SDes, SEh, SEr, GF, IF, TG, ALG, OGr, NAG, AKH, UKH, DPH, SH, DFH, AJH, NJ, RK, DCML, PMa, AFM, KLM, SM, KN, WJN, PAN, JO, SLR, RR, GVR, PGS, CLS, LSc, DTT, MCV, DvE, NJAv, LNV, HW, JMW, WW, LTW, AW, MPZ.

### Genetic data analysis

LA, JCB, JGT, RMB, QC, CRKC, SC, EJCdG, PL.DJ, SDeb, SDes, SD, SEh, MF, TG, KLG, NAG, DPH, EH, MK, MJK, SLH, PHL, SL, DCML, ML, YM, BLM, BM, KN, SLR, GVR, PGS, MSa, CLS, RS, PRS, MSc, LSh, JS, AVS, DvE, DvdM, CW, JY, LMGM, AIC, MER.

## Methods

### Statistics

This study performed several statistical approaches including linear regression, linear mixed effects associations, genome-wide association studies, LD-score regression, bivariate gaussian mixture models, genomic structural equation modelling, and MTAG-based meta analysis of GWAS summary statistics. Each approach is described in detail below.

### Cohorts and GWAS

#### ENIGMA and CHARGE

GWAS summary statistics for the MRI-derived volume of seven subcortical brain structures of interest (nucleus accumbens, amygdala, brainstem, caudate nucleus, globus pallidus, putamen, and thalamus) were obtained from the ENIGMA website following the application and approval of this project. These GWAS summary statistics are detailed elsewhere^2^. This compilation of GWAS summary statistics is the product of a meta-analysis including 48 European ancestry samples from the ENIGMA consortium^57^, the CHARGE consortium^58^ and the first release (N∼8,312) of the UK Biobank neuroimaging traits. Individual cohorts conducted quality control on their genotypic data (including SNP and sample level quality for MAF, missingness and heterozygosity), and phenotypic data (including outlier screening and distribution checks) prior to imputation. GWAS followed standardized ENIGMA/CHARGE analysis plans. Quality control before the meta-analysis of these samples included removing SNPs with poor imputation quality, removal of non-common SNPs (minor allele frequency > 0.01) and SNPs with a low effective minor allele count (< 20) or not represented across the meta-analysis (i.e. present in less than 70% of the total sample size for the discovery GWAS). Furthermore, a sample size (Z-score) weighted meta-analysis was used, as cohorts used different methods for acquisition, processing and adjustment of GWAS. The UK Biobank sample was adjusted for total brain volume, whereas ENIGMA and CHARGE consortium data were adjusted for total ICV^2^. Results from a previous GWAS meta-analysis for ICV^15^ and hippocampal volume^16^ were obtained via a public access repository through application and approval (https://enigma.ini.usc.edu/research/download-enigma-gwas-results/). Strict MRI-scan protocol procedures were followed to ensure high data quality as described thoroughly elsewhere^2^.

#### UK Biobank

We performed GWAS for intracranial and nine subcortical brain volumes with data from the UK Biobank^59^. The UK Biobank genotyping and phenotyping have been described elsewhere^60^. Briefly, our GWAS includes 36,095 participants of European ancestry passing standard quality control procedures as described elsewhere^60^. The subcortical brain structures included the nucleus accumbens, amygdala, brainstem, caudate nucleus, hippocampus, globus pallidus, putamen, thalamus, and ventral diencephalon. We also performed a GWAS on ICV. We excluded outlier measures that were at least four standard deviations from the mean. GWASs were performed using BOLT-LMM (v2.3.2)^61^, which accounts for relatedness via a linear mixed model. This method includes a random effect with a variance-covariance structure specified by a genetic-relatedness matrix (GRM) derived from a subset of SNPs across the genome^61^. The GWAS was adjusted for genotyping array, sex, age, sex*age, age-squared, sex*age-squared, and the first 20 genetic principal components to adjust further for population stratification. We included the neuroimaging data collection site (Data Field 54) as a covariate in the model to account for potential bias due to the use of different scanners across data collection sites. GWASs for subcortical brain volumes were further adjusted for ICV. We excluded variants with a low minor allele frequency (<0.01) or a low-quality imputation score (<0.60) from the analysis. Strict MRI-scan protocol procedures were followed to ensure high data quality as described thoroughly elsewhere^62^.

In the present study, we did not have an independent sample to perform replication analyses. Nonetheless, we leveraged the total sample from the UK Biobank included in our meta-analyses to create two subsamples of N ∼ 18,047. These subsamples were created by randomly splitting the main sample N = 36,095 into two sets of data. We used these subsamples to conduct GWAS for intracranial and subcortical brain volumes as an alternative replication method to compare GWAS findings between these samples and with the meta-analyses. We included the same covariates as described above and performed the same quality control procedure.

Throughout the main set of GWAS analyses and for the meta-analyses we included ICV as a covariate in the GWAS to account for inter-individual variation in subcortical brain volume due to head size differences, which is crucial when using samples including participants from different age groups. However, as previous studies have suggested^54^, adjusting for heritable covariates, such as ICV, could bias effect estimates in GWAS. Therefore, as a sensitivity analysis, we also performed GWAS in the full sample of the UK Biobank cohort (N = 36,095) as described above for nine subcortical brain volumes, but without including ICV as a covariate in the model. This allowed us to understand potential differences in GWAS with and without correcting for ICV.

#### ABCD

The ABCD study is a longitudinal resource that includes children aged nine and ten at recruitment^63^. Conducted in the United States, neuroimaging measures were obtained by the ABCD Data Analysis and Information (DAIC) and the Image Acquisition workgroups. Neuroimaging was performed across 21 sites using three different scanner types. Further information on image acquisition and postprocessing is available elsewhere^64,65^. Brain volumes analysed in this cohort included ICV, hippocampus, ventral diencephalon, brainstem, nucleus accumbens, caudate nucleus, thalamus, globus pallidus, amygdala, and putamen —volumes of the left and right measures (where relevant)— were averaged for each individual. We excluded outlier measures that were at least four standard deviations from the mean. Saliva samples were obtained at a baseline visit, and genotyping was performed using a Smokescreen array following standard DNA extraction protocols. Quality control removed genetic variants with a low call rate (less than 99% of the sample) and samples with a missing rate greater than 20% or conflicting identifiers. This quality-controlled dataset was imputed to the 1000G Phase 3 reference panel using the Michigan Imputation Server ^66^. Imputed genotype probabilities were extracted from the imputed data using QCTOOL v2 (https://www.well.ox.ac.uk/~gav/qctool_v2/). PLINK v2 was used to generate a subset of genetic files as a genetic relatedness matrix (GRM) for GWAS analysis. Briefly, a random list of 500,000 variants passing QC (minor allele frequency >= 0.01; CR >= 0.9 and INFO >= 0.6) was generated and used to create a new set of *PLINK* files (.bed, .bim, .fam) from the imputed genotype probability files. Ancestry was inferred by projecting the ABCD samples onto the principal components of the 1000 Genomes project using PLINK v1.90b6.8 and the flag --pca-clusters (**Supplementary Figure 128**). The Euclidean distance between the centroids for the first three principal components of each 1000G super population and each sample was calculated using Python (v3.5). To assess the validity of this approach, a receiver operating characteristic (ROC) curve was used to investigate whether this distance (multiplied by -1) was able to classify samples according to self-reported white race (which could be considered a proxy for European ancestry). Participants were deemed outliers (i.e., non-Europeans) if they were more than three standard deviations from the super population centroid. Importantly, this cutoff value was close to Youden’s *J*, which could be considered (post-hoc) the optimal cutoff for binary classification (**Supplementary Figure 129**). The final GWAS included 5,267 participants of European ancestry who passed genetic and neuroimaging quality control. The GWAS was performed using BOLT-LMM (v2.3.2) adjusting for age, sex*age, age-squared, sex*age-squared, and the first 20 genetic principal components to adjust further for population stratification. We included the imaging device serial number under the variable name ‘mri_info_deviceserialnumber’ as a covariate in the model, as suggested in previous studies^64^, to account for potential bias due to the use of different scanners across data collection sites. Subcortical volumes GWAS were further adjusted for ICV. We excluded variants with a low minor allele frequency (<0.01) or a low-quality imputation score (<0.60) from the analysis. Strict MRI-scan protocol procedures were followed to ensure high data quality as described thoroughly elsewhere^67^.

### Intracranial and subcortical brain volumes GWAS meta-analyses

We performed a GWAS meta-analysis for each brain volume phenotype across the ENIGMA-CHARGE published summary statistics and the GWAS in the UK Biobank and ABCD performed here, yielding a total sample size of up to 74,898 unique participants of European ancestry across all samples (**Supplementary Table 32**). All participants included in the present study provided written informed consent and the investigators on the participating studies obtained approval from their institutional review board or equivalent organisation. Individual GWAS for subcortical brain volumes were adjusted for ICV, as this reduces inter-individual variation in subcortical brain volume simply due to head size differences^68^. The meta-analyses were performed using MTAG v1.0.8^69^. Meta-analyses were performed, assuming equal heritability and perfect genetic covariance. Independent loci for human intracranial and subcortical brain volumes were determined by combining lead SNPs for all brain volumes under study and performing a conservative clumping procedure in PLINK 1.9^70^ (p^1^ = 1×10^−8^, p^2^ = 1×10^−5^, r^2^ = 1×10^−3^, kb = 1000). Independent genome-wide loci not reported in previous studies are claimed based on a comparison of the independent unique loci identified in the present study across intracranial and subcortical brain volumes with independent genome-wide significant loci for intracranial^14,15^ or subcortical brain^2,16,30^ volumes reported in previous studies. We considered linkage-disequilibrium information in the definition of the independent genome-wide loci not reported in previous studies by performing a clumping procedure using PLINK 1.9^70^ (p^1^ = 1×10^−8^, p^2^ = 1×10^−5^, r^2^ = 1×10^−3^, kb = 1000). We report results at the common genome-wide significance threshold. In addition, we performed multiple testing correction using matSpD to account for the total number of phenotypes as performed in previous studies^12^. We observed that the effective number of independent traits in our analysis was 8. Thus, we set a significance threshold of *p*-value < 5×10^−8^ / 8 = 6.25×10^−9^.

The imaging analysis and visualization of structural data in all cohorts was performed using the publicly available FreeSurfer^71^ package tool (https://surfer.nmr.mgh.harvard.edu/) developed by the Laboratory for Computational Neuroimaging at the Athinoula A. Martinos Center for Biomedical Imaging. Details regarding border definition for specific brain structures is available on the wiki (https://surfer.nmr.mgh.harvard.edu/fswiki/FreeSurferWiki). In particular, the ventral diencephalon contains the following structures: the hypothalamus, basal forebrain, the sublenticular extended amygdala (SLEA), and a portion of the ventral tegmentum, which can also be considered a part of the midbrain^71,72^. These specific substructures do not overlap with the brainstem borders, which is constituted by the medulla oblongata, pons, midbrain and superior cerebellar peduncle^71^.

The phenogram in **Figure 1** was created using the Ritchie Lab Visualization online tool (https://visualization.ritchielab.org/phenograms/plot). Subcortical brain images in **Figure 1** were created using publicly available tutorials (https://surfer.nmr.mgh.harvard.edu/fswiki/CorticalParcellation and https://bookdown.org/u0243256/tbicc/freesurfer.html) in MATLAB (R2023b). The ENIGMA consortia also provides tutorials on the creation of brain-related figures (https://enigma-toolbox.readthedocs.io/en/latest/pages/12.visualization/index.html#subcortical-surface-visualization).

### Functional annotation and gene prioritisation

We performed functional annotation and gene prioritisation analyses using MAGMA, eQTL mapping with TWAS, and by integrating single cell sequencing data with GWAS summary statistics.

First, we performed gene-based tests using MAGMA^73^ v(1.08) as implemented in FUMA (v1.5.2)^74^ (https://fuma.ctglab.nl/). The MAGMA method provides aggregate association *p-*values based on all variants within a gene and its regulatory region^73^. We applied a Bonferroni multiple-testing correction based on the total number of genes and accounted for the effective number of independent traits in our analysis, (0.05 / 17,708 [average number of tests per brain volume] * 8 [estimated number of independent phenotypes] = 2.26×10^−5^).

Second, we conducted an in-depth analysis of genetically regulated gene expression using FUSION (http://gusevlab.org/projects/fusion/), a software tool for TWAS^75^. FUSION leverages SNP-gene expression associations to construct predictive linear models tailored to each gene. The model demonstrating superior predictive performance in cross-validation trials was subsequently employed for predictive applications within the GWAS. Available tissue specimens sourced from five distinct subcortical regions from GTEx v8 (specifically, Accumbens, Amygdala, Hypothalamus, Hippocampus, and Putamen) were included in the analysis. For this, we used a Mendelian randomization framework employing summary-data-based Mendelian Randomisation (SMR) v1.3.1^76^ to assess gene expression in multiple cell lines across the nine subcortical and intracranial volumes (ICV). We also incorporated data from RNA splicing sequencing, based on single-tissue gene expression derived from the brain. We applied Bonferroni multiple testing correction and accounted for the effective number of independent traits in our analysis (0.05 / 1,308 [average number of annotations per brain volume] * 8 [estimated number of independent phenotypes] = 3.06×10^−4^). Moreover, we utilised an eQTL dataset derived from 120 human fetal brains^77^, employing the SMR method to identify genes involved in the development of subcortical brain structures. We applied Bonferroni multiple testing correction and accounted for the effective number of independent traits in our analysis (0.05 / 317 [average number of annotations per brain volume] * 8 [estimated number of independent phenotypes] = 1.26×10^−3^).

Genes prioritised through MAGMA and FUSION analyses from single and multiple brain tissues were further assessed by integrating GWAS summary data with single-cell RNA-sequencing data, which included over 1 million cells at three different stages of the differentiation process. Single-cell RNA-Seq analysis was based on eQTL data of Jerber et al. from 215 human induced pluripotent stem cell lines as they progressed towards a midbrain neural-like fate^28^. This process encompasses the generation of dopaminergic neurons, serotonin transporters, astrocyte-like cells, ependymal cells, and neuron-differentiated clusters. We filtered results for those involving genes associated with intracranial or subcortical brain volumes across MAGMA and TWAS analyses. Then, we applied Bonferroni multiple testing correction technique, considering the effective number of independent traits in our analysis (0.05 / 337 [total number of gene-brain volume associations] * 8 [estimated number of independent phenotypes] = 1.19×10^−3^).

As a sensitivity analysis, we sought to understand potential differences in GWAS for subcortical brain volumes with and without correcting for ICV. Therefore, we performed gene-based tests using MAGMA^73^ v(1.08) as implemented in FUMA^74^ for GWAS in the UK Biobank cohort with and without adjusting for ICV. For each set of GWAS summary statistic (i.e., with and without adjusting for ICV), we applied Bonferroni multiple testing correction technique, considering the effective number of independent traits in our analysis (0.05 / 1097 [total number of gene-brain volume associations] * 8 [estimated number of independent phenotypes] = 3.64×10^−4^).

### SNP-based heritability and genetic correlations

LD score regression (LDSC)^78^ was used to estimate the heritability for each subcortical brain structure. Briefly, this method leverages the expected relationship between the linkage-disequilibrium variant tags and their expected degree of association with a given phenotype to estimate the heritability. It distinguishes between confounding bias and polygenicity^78^. We processed our meta-analysis results using the *munge* function from LDSC v.1.0.1 and performed LD score regression to estimate the percentage of variance explained by the SNPs in the meta-analysis.

The genetic correlation between a pair of phenotypes depicts the relationship of genetic effect sizes at mutual genetic variants across phenotypes^79^. In the present study, we used LD score regression to perform genetic correlation analyses among subcortical brain structures and between complex human phenotypes, including neuropsychiatric disorders and anthropometric measurements, with subcortical brain structures. Details for the GWAS summary statistics for neuropsychiatric and subcortical brain structures are provided in the **Supplementary Table 33** and **Supplementary Methods**. These complex human phenotypes were selected based on criteria applied in previous studies by the ENIGMA consortium, which relies on the public availability of well-powered summary statistics of previously reported brain-related phenotypes and anthropometric measurements^2,15^. These criteria are limited and restricted in the present study by the data transfer agreement with the CHARGE cohort, for which we are not allowed to leverage CHARGE data to investigate any relationships involving substance-related disorders and cognitive or intelligence-related phenotypes. We accounted for multiple testing using Bonferroni correction (0.05 / 320 [total number of genetic correlation tests] = 1.56×10^−4^).

As a sensitivity analysis, we sought to understand potential differences in GWAS for subcortical brain volumes with and without correcting for ICV. Therefore, we estimated the genetic correlation between the GWAS for subcortical brain volumes in the UK Biobank cohort with and without adjusting for ICV. In addition, we estimated genetic correlations for both sets of GWAS summary statistics (i.e., with and without adjusting for ICV) with complex human phenotypes.

### Pairwise GWAS

We leveraged the pairwise-GWAS (v.0.3.6) method^80^ to identify segments of the genome with genomic variants influencing the aetiology of a brain volume and a human complex phenotype. For each pair of genetically correlated phenotypes after multiple testing correction according to our LD score regression results, we conducted GWAS-PW analyses. This method splits the genome into 1703 independent segments and, for each segment, GWAS-PW estimates the posterior probability of association (PPA) for four different models. These models include (i) the genomic segment is uniquely associated to phenotype A, (ii) the region is uniquely associated to phenotype B, (iii) the segment of the genome is influencing the aetiology of both phenotypes through the same genetic variants, and (iv) the genomic segment is involved in the aetiology of both phenotypes via different genetic variants. We provide findings for segments of the genome where model three (the genomic segment is influencing the aetiology of both phenotypes through the same genetic variants) had a PPA > 0.5, given that this threshold has been used in previous studies^81,82^.

### Bivariate MiXeR

We conducted bivariate MiXeR analyses using MiXeR v1.3^83^ to quantify polygenicity among the nine subcortical brain volumes under study. This analysis has been thoroughly described elsewhere^83^. Briefly, MiXeR leverages GWAS summary statistics and a univariate gaussian mixture model to estimate the degree of polygenicity (irrespective of genetic correlation), which is commonly referred to as the number of trait-influencing genetic variants. Then, with a bivariate gaussian mixture model, the additive genetic effect of four components is estimated for every pair of phenotypes: (i) genetic variants that do not influence either phenotype, (ii) genetic variants that only influence phenotype A (iii) genetic variants that only influence phenotype B, and (iv) genetic variants that influence both phenotypes^83^. Thus, MiXeR provides information about the genetic associations between two complex phenotypes as it estimates the total number of shared and phenotype-specific causal variants.

### Genetic factor analyses

To examine genetic clustering of the nine subcortical brain structures we conducted exploratory factor analyses (EFA) based upon the LDSC-derived genetic correlation matrix. The R (v3.5.1) package ‘psych’ was used to conduct the EFAs, with a maximum likelihood extraction method and oblimin rotation method. The factor models identified in the EFA (retaining factor loadings > 0.25) were subsequently carried forward in a confirmatory factor analysis (CFA) in genomic SEM. This was done to assess the fit of the factor model to the data while taking into account uncertainty in covariance estimates. The default diagonally weighted least squares estimator was used.

### Potential causal genetic effects

We leveraged the latent causal variable (LCV)^84^ and Latent Heritable Confounder Mendelian Randomisation (LHC-MR v0.0.0.9000)^85^ methods to investigate potential causal genetic effects between brain volumes under study and those complex human traits that displayed a statistically significant genetic correlation after Bonferroni multiple testing correction.

We employed the LCV method, which has been thoroughly described elsewhere^79,84^, to assess whether the genetic correlations identified in the present study could be explained by putative causal genetic effects and accounted for multiple testing using a Bonferroni correction [0.05 / 16 [total number of genetic causal proportion tests in the present study] = 3.13×10^−3^]. Advantages of the LCV method include that (i) is it less susceptible to confounding by horizontal pleiotropic effects, (ii) it leverages aggregated information across the entire genome (i.e., full genome-wide data) to increase statistical power, and (iii) it is robust to sample overlap^84^.

In the LCV method, the sign of the GCP parameter denotes the direction of potential causal genetic effects^84^. The GCP parameter ranges from -1 to 1, where GCP = 1 suggests full putative causal genetic effects of phenotype A on phenotype B. Conversely, GCP = -1 suggests full putative causal genetic effects of phenotype B on phenotype A. Moreover, a GCP = 0 implies the detection of horizontal pleiotropy, suggesting that an intervention on one phenotype would not affect the other due to the absence of causal genetic effects. Overall, to interpret LCV findings one must consider three important factors: (i) the magnitude of the genetic correlation, (ii) the GCP estimate, and (iii) the direction (positive or negative) of the GCP estimate^84,86–88^.

LHC-MR leverages full GWAS summary statistics (not only genome-wide independent loci like traditional MR methods) to investigate potential causal genetic effects between a pair of genetically correlated phenotypes. LHC-MR has been reported to improve statistical power to estimate bi-directional putative causal genetic effects, direct heritabilities, and confounder effects while accounting for sample overlap. LHC-MR has been suggested to outperform a number of traditional MR methods^85^. Full details for the LHC-MR method are described elsewhere^85^. We accounted for multiple testing using a Bonferroni correction [0.05 / 32 [total number of LHC-MR tests in the present study] = 1.56×10^−3^]. We performed LHC-MR analyses with R (v3.5.1).

### Polygenic scores estimation and association analyses

We performed the meta-analysis again but without the ABCD cohort to ensure sample independence and test polygenic prediction in European (N = 5,267), non-European (N = 5,173), African-only (N = 1,833), Asian-only (N = 152), and all samples (N = 10,440). Non-European ancestry individuals include, but are not limited to african-only and asian-only ancestries as individuals with admixed ancestry were also considered. To avoid bias due to the correlation between SNPs arising from linkage-disequilibrium (LD), a Bayesian analysis was used to approximate the results of a conditional GWAS (i.e. one estimating the effect for all SNPs simultaneously). This was performed using *SBayesR*^*89*^ implemented within the *Genome-wide Complex Trait Bayesian analysis* (GCTB v2.0) software tool^90^. Polygenic scores for intracranial and subcortical brain volumes were estimated by multiplying the multivariate effect size (obtained from *SBayesR*) times the allelic dosage of the effect allele and summing across all loci for each participant. Only SNPs passing quality control (minor allele frequency > 0.01, call rate > 0.9 and imputation score > 0.6) were included in the derived polygenic scores. To test for the association between intracranial and subcortical brain volumes polygenic scores with their corresponding phenotype and estimate the percentage of phenotypic variance explained, we performed a linear mixed effects model, in GCTA version 1.91.7 beta1, with a random effect and with a variance-covariance specified by a genetic relatedness matrix to account for cryptic relatedness among participants of the ABCD cohort. The results were plotted in Python (v3.5) using *seaborn, matplotlib* and in-house scripts. Sensitivity analyses assessed whether differential variance explained within the ABCD cohort was due to differential ancestry, sample size differences, or cryptic relatedness. These analyses consisted of (i) using a clumping and thresholding approach to derive polygenic scores with a linear mixed effects model implemented in GCTA to perform the prediction; (ii) performing the association analyses using a multivariate linear regression in Python (v3.5), and the library *statsmodels*. Additional covariates included in the model were sex, age, and the first twenty genetic ancestry components to adjust for population stratification. The percentage of variance explained was estimated as the difference in R^2^ between the full model (i.e., including the polygenic scores) and a reduced model including only covariates; and (iii) Using *SBayesR* derived polygenic scores to perform associations analyses with multivariate linear regressions among participants of European ancestry including ICV as one of the covariates.

## Ethics statement

Our study is based on meta-analysis of previously published, publicly available data for which appropriate site-specific Institutional Review Boards and ethical review at local institutions have previously approved the use of these data. For full-details on the institutions that have approved the use of these data please refer to the ‘Acknowledgments’ section.

## Data availability

Detailed information on how to access publicly available GWAS summary data from the ENIGMA and CHARGE consortia is reported on their corresponding publications^2,12,15^. Researchers can access individual-level data from the UKB and ABCD cohorts following the corresponding data application procedures. Work performed using UKB data was done under application 25331. Full genome-wide summary statistics generated in the present study are available at the ENIGMA website (http://enigma.ini.usc.edu/research/download-enigma-gwas-results).

## Code availability

No custom code was used in this study. Publicly available software tools were used to perform genetic analyses and are referenced throughout the manuscript.

