## SupplementaryNote for "Genomic analysis of intracranial and subcortical brain volumes yields polygenic scores accounting for variation across ancestries"

### Supplementary methods

#### GWAS for complex human phenotypes

We leveraged GWAS summary statistics, including individuals of European ancestry, for various complex human phenotypes, including neuropsychiatric disorders, (**Supplementary Table 33**) to perform genetic correlation analyses. A detailed description of each of the GWAS summary statistics for each phenotype is available in their corresponding publication. Below, we briefly describe the main characteristics of each dataset. GWAS summary statistics from the Psychiatric Genetics Consortium (PGC) are publicly available at https://pgc.unc.edu/for-researchers/download-results/. For depression and Parkinson's disease, the full set of GWAS summary statistics, including samples for 23andMe, were accessed via a Data Access Request Form https://research.23andme.com/dataset-access/. Other phenotypes can be accessed following instructions in the *Data Availability* section in their corresponding publication.

Publicly available GWAS summary statistics for ADHD included 38,691 cases and 186,843 controls [1]. An inverse variance-weighted GWAS meta-analysis of childhood ADHD classified under DSM-IV was performed on samples from the PGC, deCODE and the Lundbeck Foundation Initiative for Integrative Psychiatric Research (iPSYCH) [1]. Strict quality control and imputation procedures were performed [1]. Principal components were included as covariates in the model to control for putative population stratification [1]. This dataset is publicly available on the PGC's website.

Publicly available GWAS summary statistics for Alzheimer's disease were retrieved from the GWAS catalogue (https://www.ebi.ac.uk/gwas/) under accession number GCST90027158 [2]. These summary statistics included 111,326 clinically diagnosed/proxy cases and 677,663 controls from several cohorts, such as the Rotterdam study, FinnGen and the UK Biobank. Briefly, a two-stage GWAS was performed [2]. Stage I included several GWAS analyses of different samples for clinically diagnosed and proxy cases. These GWAS were conducted using logistic regression and an additive genetic model as implemented in SNPTEST 2.5.4-beta3 [3], PLINK v1.90 [4], or SAIGE [5]. In stage II, using GWAS from stage I, a fixed-effect meta-analysis with an inverse variance weighted approach was performed as implemented in METAL [6].

Publicly available GWAS summary statistics for bipolar disorder [7] were obtained from the PGC's public repository. These summary statistics include 41,917 cases and 371,549 controls from 57 cohorts collected in Europe, North America and Australia [7]. Briefly, for each cohort, a GWAS was conducted using an additive logistic regression model in PLINK v1.90 [4]. Then, a GWAS meta-analysis was performed using an inverse-variance-weighted fixed-effects model in METAL (version 2011-03-25) [6].

GWAS summary statistics for depression were obtained from the PGC and through a Data Access Request Form via 23andMe [8]. An inverse-variance weighted meta-analysis was performed on samples from the UK Biobank, the PGC, and 23andMe, including 246,363 cases and 561,190 controls [8]. This dataset, excluding the samples from 23andMe, is publicly available on the PGC's website.

Publicly available GWAS summary statistics for neuroticism score, excluding samples from 23andMe, were retrieved from the Department of Complex Trait Genetics public repository (https://ctg.cncr.nl/software/summary_statistics) [9]. These summary statistics include 449,484 individuals from the UK Biobank and the Genetics of Personality Consortium [10]. This GWAS meta-analysis was performed using a two-sided sample-size-weighted fixed-effects analysis as implemented in METAL [6].

GWAS summary statistics for Parkinson's disease included 56,306 cases and 1,417,791 controls, yielding a total sample size of 1,474,097 [11]. These summary statistics represent a meta-analysis from several different cohorts, including the International Parkinson's Disease Genomics Consortium (IPDG), 23andMe, and the UK Biobank. The meta-analysis was performed using a fixed-effects model as implemented in METAL [6].

Publicly available GWAS summary statistics for schizophrenia were obtained from the PGC's public repository [12]. These summary statistics include 53,386 cases and 77,258 controls from around 90 cohorts. For each cohort, a GWAS was performed using an additive logistic regression model as implemented in PLINK [4]. Covariates included a subset of the first 20 principal components derived within each cohort. In addition, the meta-analysis of the GWAS for the individual cohorts was performed using a standard error inverse-weighted fixed-effects model [12].

Publicly available GWAS summary statistics for multisite chronic pain were retrieved from its corresponding publication [13]. This GWAS was performed on 387,649 participants of European ancestry from the UK Biobank cohort via the item *pain types experienced in the last month* (field ID 6159). Possible answers for this item included 'None of the above'; 'Prefer not to answer'; pain at seven different body sites (head, face, neck/shoulder, back, stomach/abdomen, hip, knee); or 'all over the body'. Those who reported pain, were asked if this pain had lasted for three months or longer.

GWAS summary statistics for anorexia nervosa were obtained from the PGC's public repository [14] and included 33 datasets with 16,992 cases and 55,525 controls. The Ricopili (Rapid Imputation Consortium Pipeline) was used to perform quality control procedures and imputation.

Autism spectrum disorder GWAS summary statistics were downloaded from the PGC's public repository [15]. The Ricopili pipeline was used to perform quality control, imputation, PCA, and primary association analyses. GWAS summary statistics included 8,381 individuals with autism spectrum disorder and 27,969 controls and the meta-analysis was performed using MTAG [16].

GWAS summary statistics for birth weight included 321,223 participants of European ancestry and were obtained [17] from the EGG website (https://egg-consortium.org/). The meta-analysis was performed using a fixed-effects meta-analysis in GWAMA [18].

Birth head circumference GWAS summary statistics were retrieved from the EGG website (https://egg-consortium.org/) and included 29,192 participants of European ancestry [19]. This GWAS meta-analysis was conducted with a fixed-effects inverse-variance weighted meta-analysis as implemented in METAL [6].

GWAS summary statistics for body mass index were obtained from the GIANT consortium website (https://portals.broadinstitute.org/collaboration/giant/index.php/GIANT_consortium_data_files) and included 456,426 participants of European ancestry from the UK Biobank cohort [20]. This GWAS was performed using a linear mixed model association testing implemented in BOLT-LMM v2.3.

Height GWAS summary statistics, excluding samples from 23andMe, are available at the GIANT consortium website (https://portals.broadinstitute.org/collaboration/giant/index.php/GIANT_consortium_data_files), and include 2,450,000 participants of European ancestry [21,22]. These summary statistics result from a GWAS meta-analysis conducted using a modified version of RAREMETAL [21].

Anxiety GWAS summary statistics included 31,977 cases and 82,144 controls of European ancestry in an inverse-variance weighted, fixed-effect meta-analysis in METAL [23].

Obsessive-compulsive disorder GWAS summary statistics were downloaded from the PGC's public repository [24] and included 2,688 individuals of European ancestry as cases and 7,037 genomically matched controls. The meta-analysis was performed using the inverse variance method as implemented in METAL.

GWAS summary statistics for Tourette Syndrome were obtained from the PGC's public repository and correspond to a GWAS meta-analysis in 4,819 case subjects and 9,488 controls [25]. The GWAS meta-analysis was conducted using the inverse-variance method as implemented in METAL.

Publicly available GWAS summary statistics for insomnia, excluding samples from 23andMe, were retrieved from its corresponding publication [26]. This GWAS included 109,402 cases and 277,131 controls from the UK Biobank cohort.

Publicly available GWAS summary statistics for snoring included 152,000 cases and 256,000 controls from the UK Biobank cohort. Participants were selected with the item (Field-ID: 1210) "Does your partner or a close relative or friend complain about your snoring?" which could be answered with "Yes", "No", "Don't know", or "Prefer not to answer". The GWAS was performed using BOLT-LMM to account for cryptic relatedness and population stratification. These summary statistics were retrieved from its corresponding publication [27].

GWAS summary statistics for obstructive sleep apnoea included 175,522 cases and 350,475 controls of European ancestry. The meta-analysis was performed using 28. For this study, we performed MTAG and combined sleep apnoea and snoring GWAS summary statistics [28].

GWAS summary statistics for suicide attempt included 35,786 cases and 779,392 ancestry-matched controls across 22 cohorts of European ancestry. These summary statistics were obtained from The International Suicide Genetics Consortium (ISGC) Cohort. The meta-analysis was performed using an inverse variance-weighted fixed effects model (standard error) in METAL [29].

GWAS summary statistics for genetic generalized epilepsy were retrieved from the Epilepsy Genetic Association Database (epiGAD) (epigad.org). These included 6,952 cases and 17,719 controls of European ancestry. Briefly, a P-value-based fixed-effects meta-analysis was performed using METAL [30].

### Variance in phenotype explained by SNPs (PVE)

We estimated the proportion of phenotypic variance (PVE) for intracranial and subcortical brain volumes explained by the SNPs based on the equation used to evaluate that the genetic variants selected for the exposure were strongly associated with the exposure:

$$PVE = \frac{2\beta^2 MAF\,(1 - MAF)}{2\beta^2 MAF(1-MAF) + (SE(\beta))^2 2N\, MAF\,(1-MAF)}$$

Where β is the effect of the variant, *MAF* is the minor allele frequency, *SE* the standard error and *N* is the sample size.

### Supplementary figures

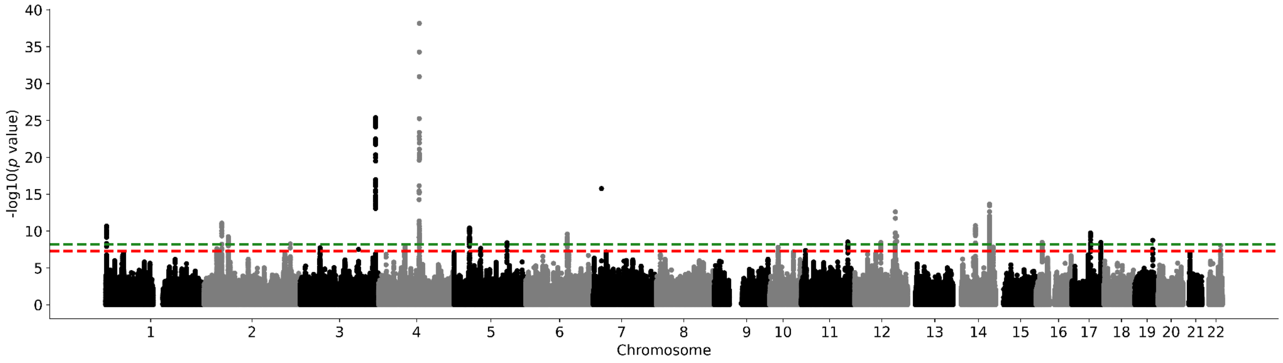

**Supplementary Figure 1. Nucleus accumbens meta-analysis Manhattan plot.** Results for nucleus accumbens GWAS. Genome-wide significance is shown for the common threshold of *p*-value < $5x10^{-8}$ (red dashed line), and also for the multiple comparisons-corrected threshold of *p*-value < $6.25x10^{-9}$ (green dashed line). The p-values referenced here correspond to a two-tailed Z-test test as implemented in MTAG.

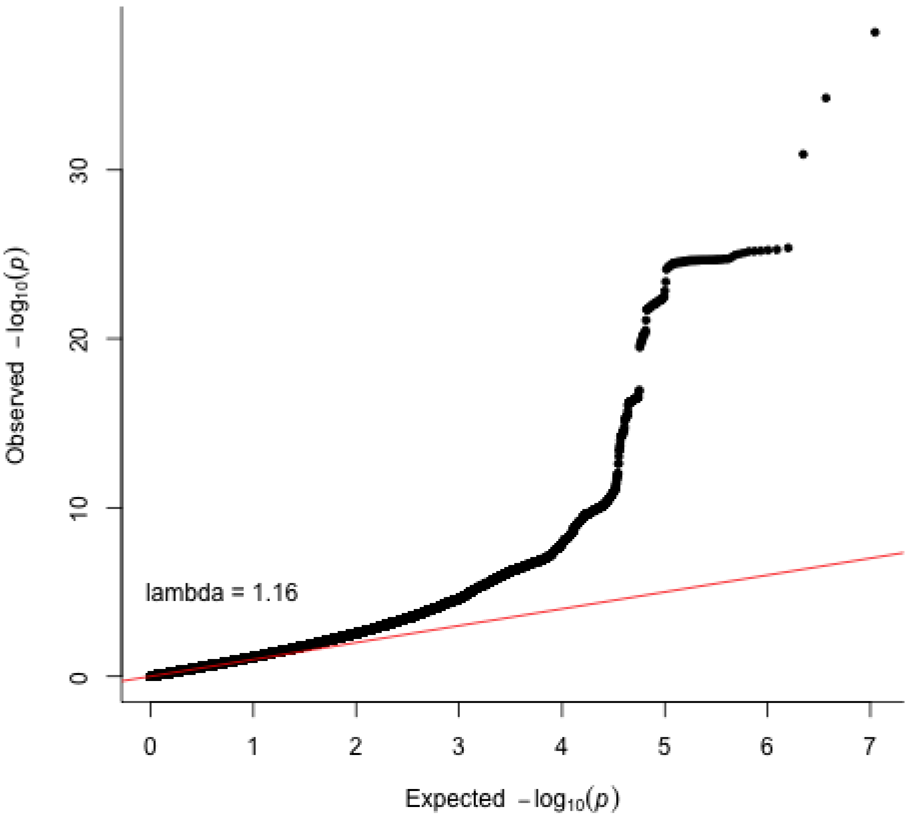

**Supplementary Figure 2. Nucleus accumbens meta-analysis QQ plot.** The p-values referenced here correspond to a two-tailed Z-test test as implemented in MTAG.

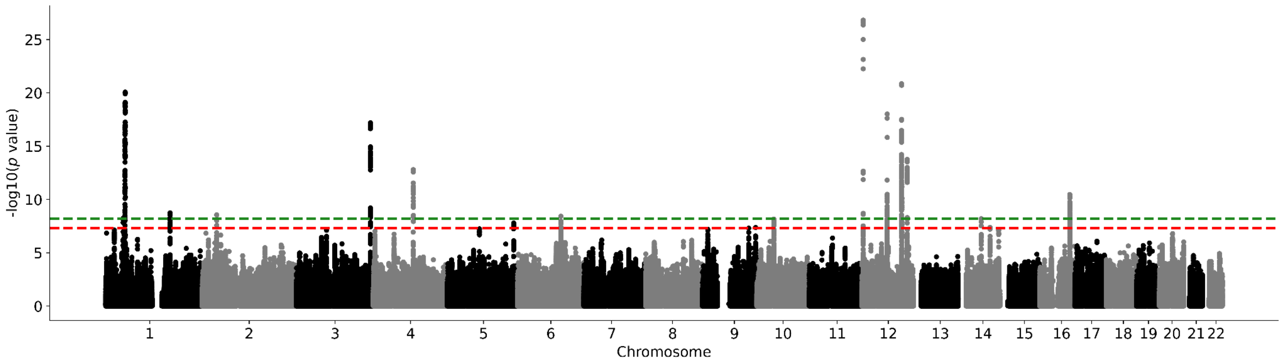

**Supplementary Figure 3. Amygdala meta-analysis Manhattan plot.** Results for amygdala GWAS. Genome-wide significance is shown for the common threshold of *p*-value < $5x10^{-8}$ (red dashed line), and also for the multiple comparisons-corrected threshold of *p*-value < $6.25x10^{-9}$ (green dashed line). The p-values referenced here correspond to a two-tailed Z-test test as implemented in MTAG.

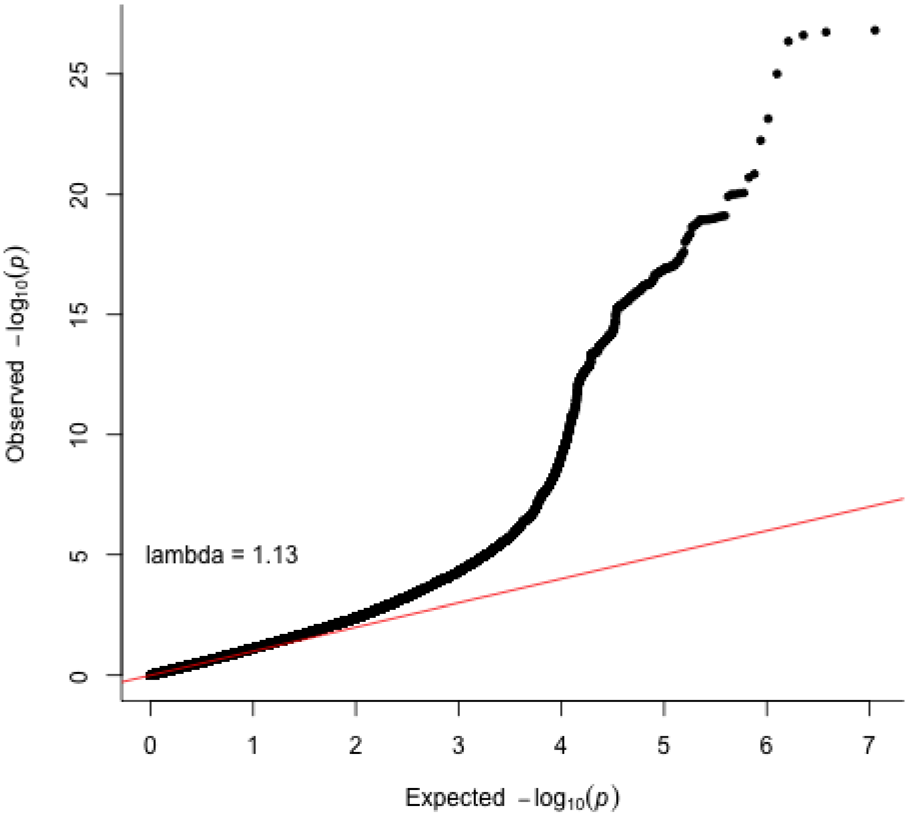

**Supplementary Figure 4. Amygdala meta-analysis QQ plot.** The p-values referenced here correspond to a two-tailed Z-test test as implemented in MTAG.

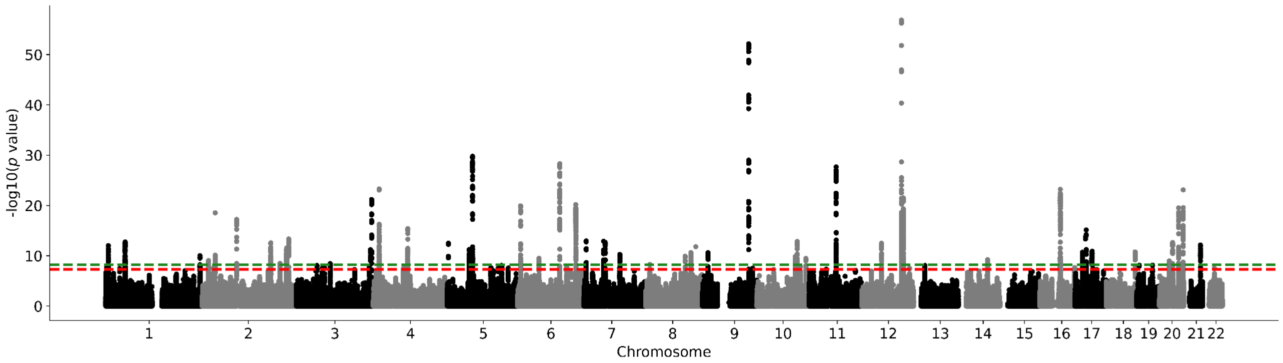

**Supplementary Figure 5. Brainstem meta-analysis Manhattan plot.** Results for brainstem GWAS. Genome-wide significance is shown for the common threshold of *p*-value < $5\text{x}10^{-8}$ (red dashed line), and also for the multiple comparisons-corrected threshold of *p*-value < $6.25\text{x}10^{-9}$ (green dashed line). The p-values referenced here correspond to a two-tailed Z-test test as implemented in MTAG.

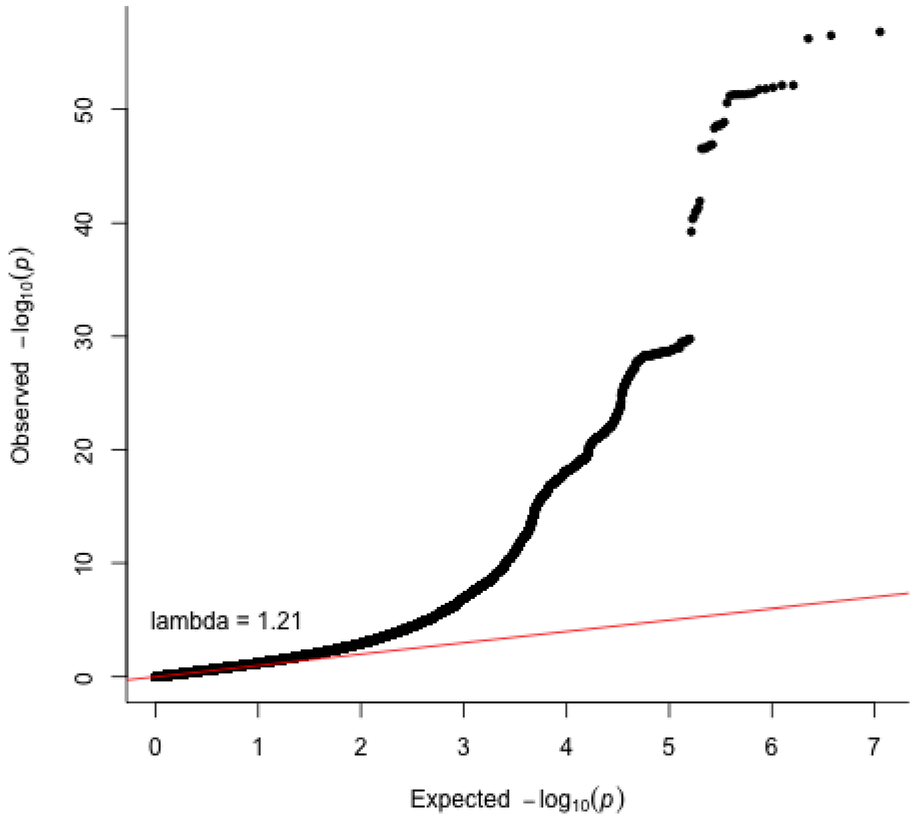

**Supplementary Figure 6. Brainstem meta-analysis QQ plot.** The p-values referenced here correspond to a two-tailed Z-test test as implemented in MTAG.

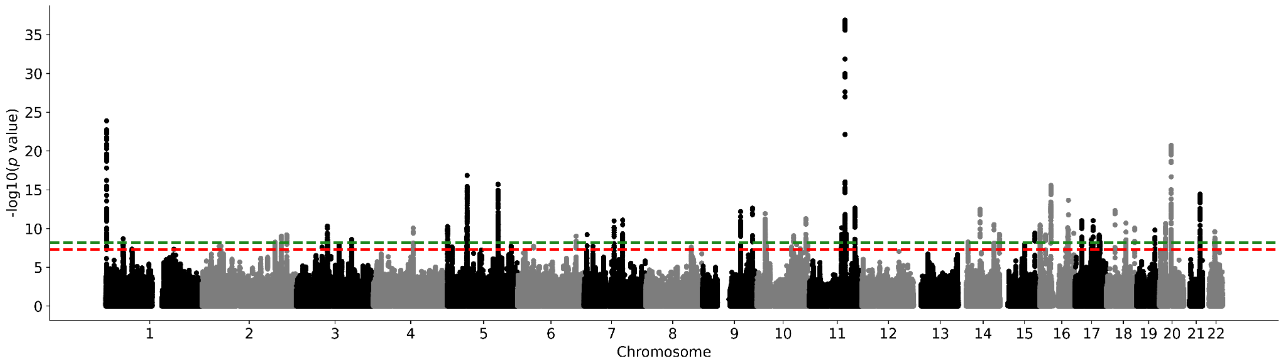

**Supplementary Figure 7. Caudate nucleus meta-analysis Manhattan plot.** Results for caudate nucleus GWAS. Genome-wide significance is shown for the common threshold of *p*-value < $5x10^{-8}$ (red dashed line), and also for the multiple comparisons-corrected threshold of *p*-value < $6.25x10^{-9}$ (green dashed line). The p-values referenced here correspond to a two-tailed Z-test test as implemented in MTAG.

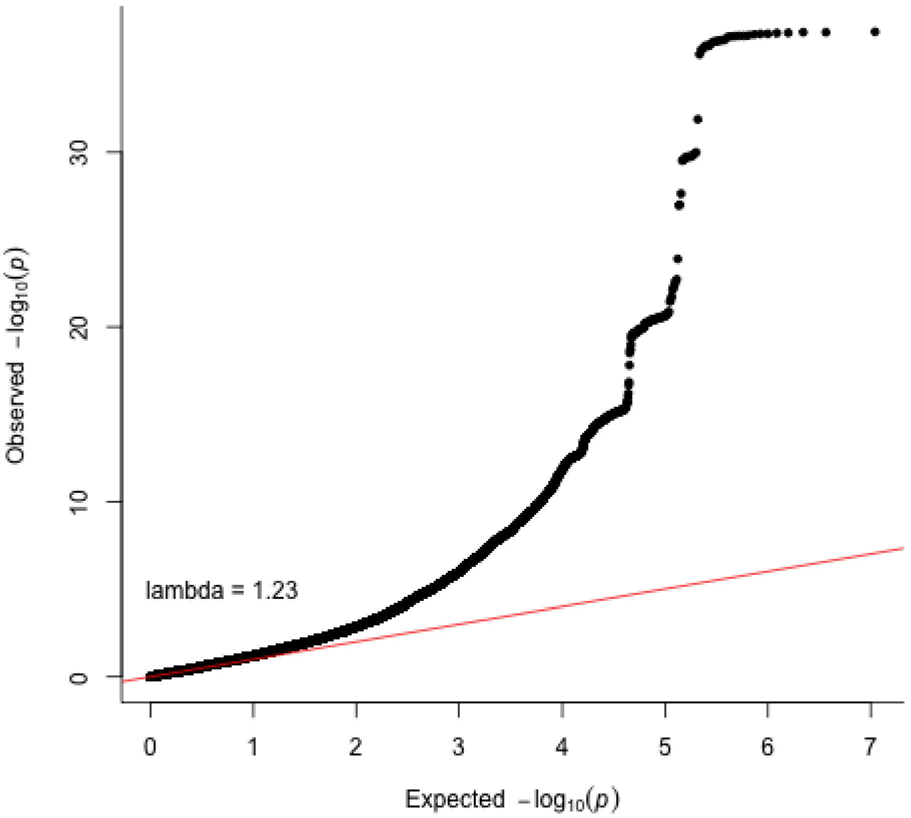

**Supplementary Figure 8. Caudate nucleus meta-analysis QQ plot.** The p-values referenced here correspond to a two-tailed Z-test test as implemented in MTAG.

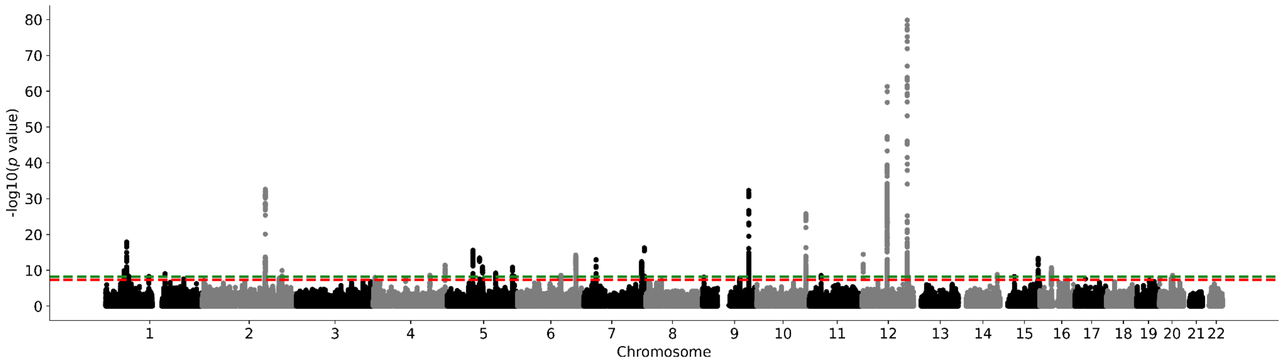

**Supplementary Figure 9. Hippocampus meta-analysis Manhattan plot.** Results for hippocampus GWAS. Genome-wide significance is shown for the common threshold of *p*-value < $5x10^{-8}$ (red dashed line), and also for the multiple comparisons-corrected threshold of *p*-value < $6.25x10^{-9}$ (green dashed line). The p-values referenced here correspond to a two-tailed Z-test test as implemented in MTAG.

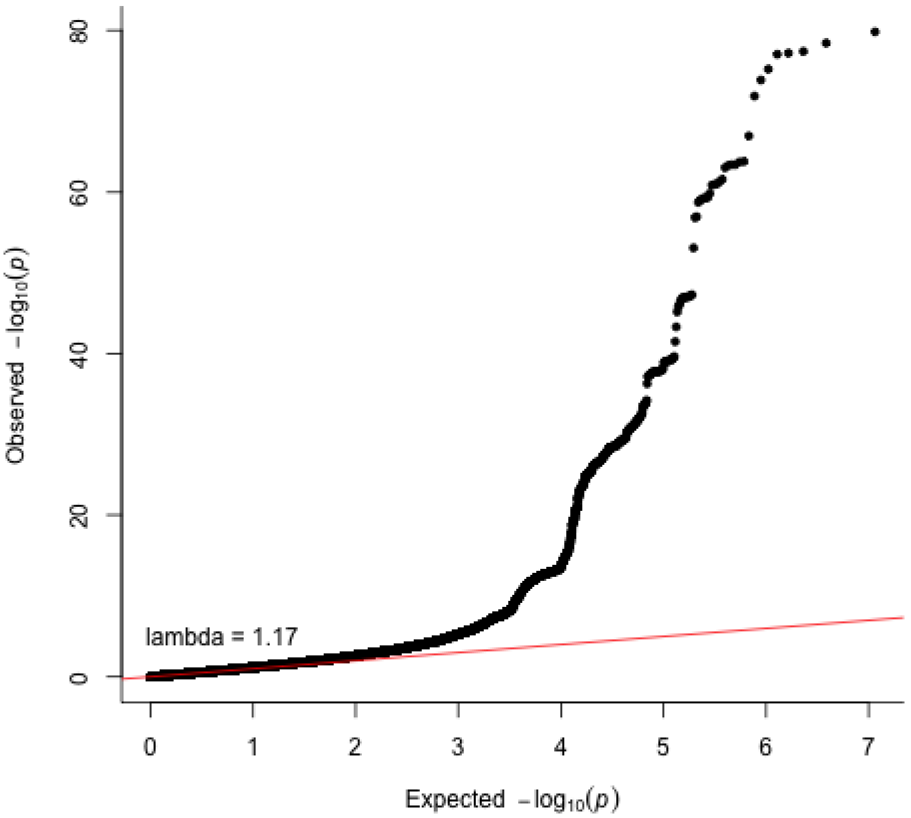

**Supplementary Figure 10. Hippocampus meta-analysis QQ plot.** The p-values referenced here correspond to a two-tailed Z-test test as implemented in MTAG.

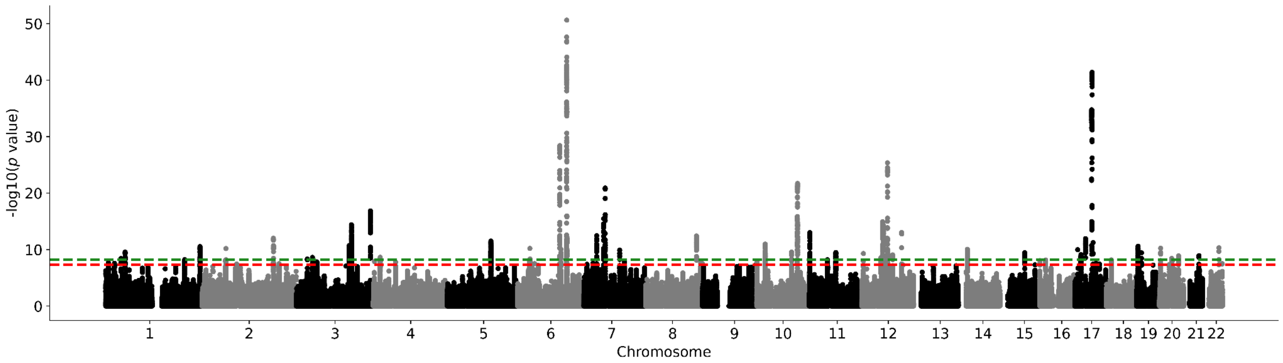

**Supplementary Figure 11. Intracranial volume meta-analysis Manhattan plot.** Results for intracranial volume GWAS. Genome-wide significance is shown for the common threshold of *p*-value < $5x10^{-8}$ (red dashed line), and also for the multiple comparisons-corrected threshold of *p*-value < $6.25x10^{-9}$ (green dashed line). The p-values referenced here correspond to a two-tailed Z-test test as implemented in MTAG.

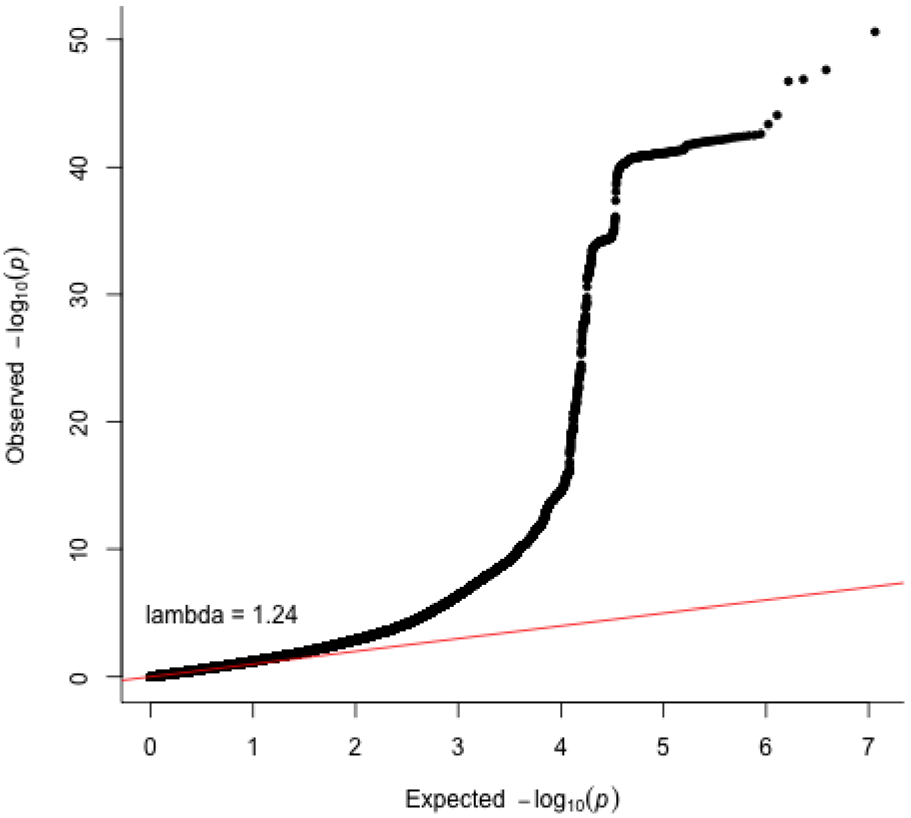

**Supplementary Figure 12. Intracranial volume meta-analysis QQ plot.** The p-values referenced here correspond to a two-tailed Z-test test as implemented in MTAG.

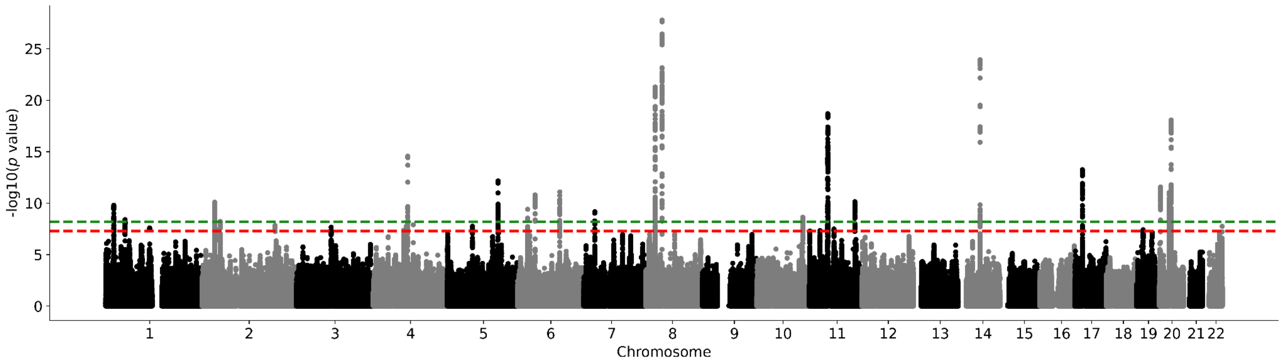

**Supplementary Figure 13. Globus pallidus meta-analysis Manhattan plot.** Results for globus pallidus GWAS. The p-values referenced here correspond to a two-tailed Z-test test as implemented in MTAG.

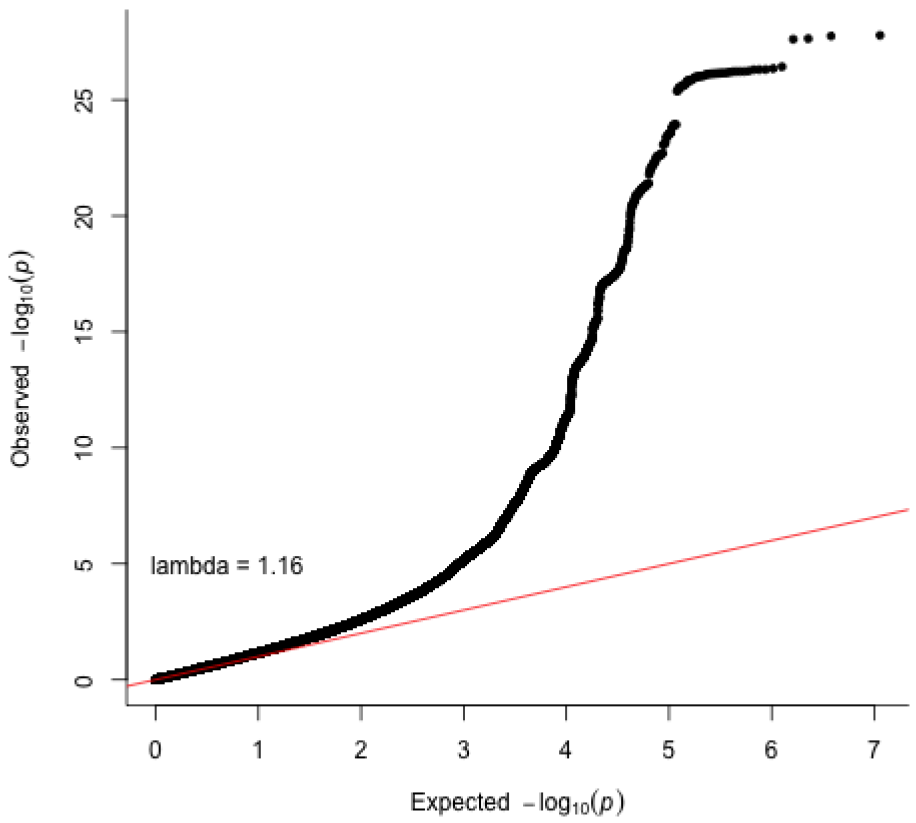

**Supplementary Figure 14. Globus pallidum meta-analysis QQ plot.** The p-values referenced here correspond to a two-tailed Z-test test as implemented in MTAG.

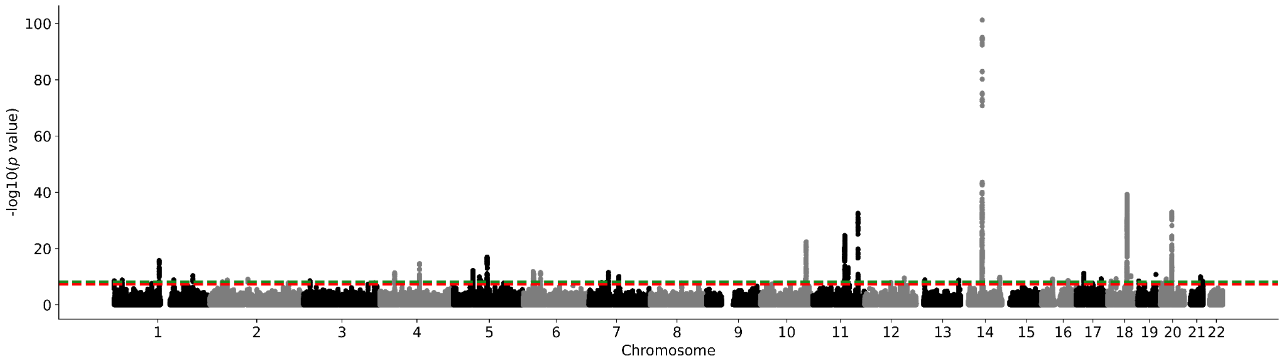

**Supplementary Figure 15. Putamen meta-analysis Manhattan plot.** Results for putamen GWAS. Genome-wide significance is shown for the common threshold of *p*-value < $5x10^{-8}$ (red dashed line), and also for the multiple comparisons-corrected threshold of *p*-value < $6.25x10^{-9}$ (green dashed line). The p-values referenced here correspond to a two-tailed Z-test test as implemented in MTAG.

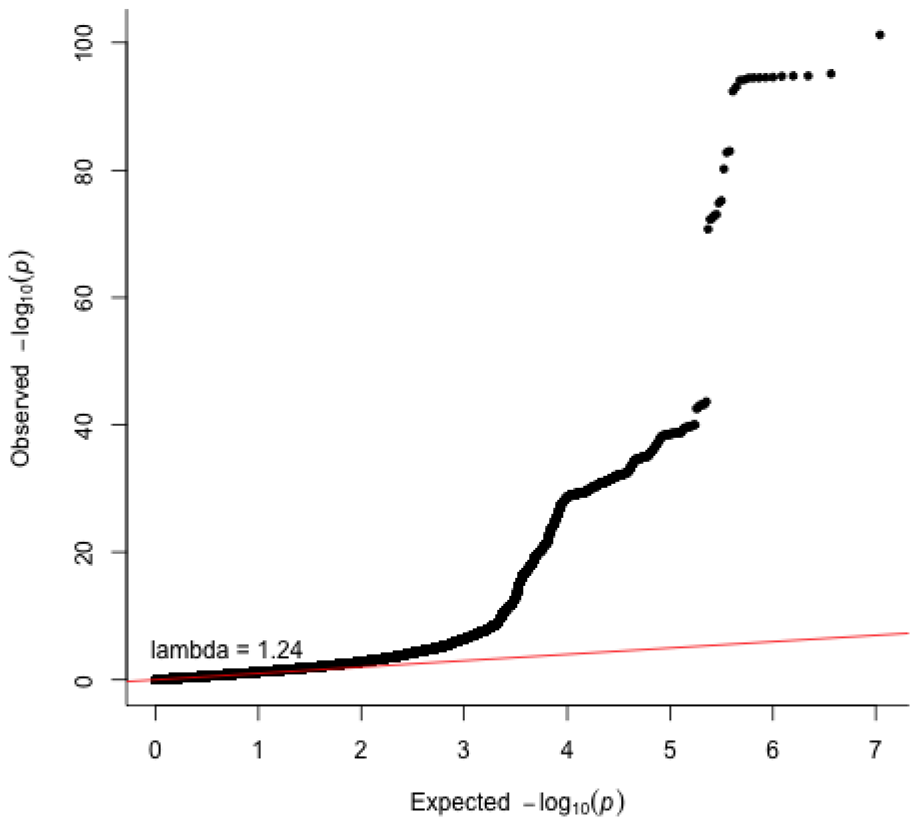

**Supplementary Figure 16. Putamen meta-analysis QQ plot.** The p-values referenced here correspond to a two-tailed Z-test test as implemented in MTAG.

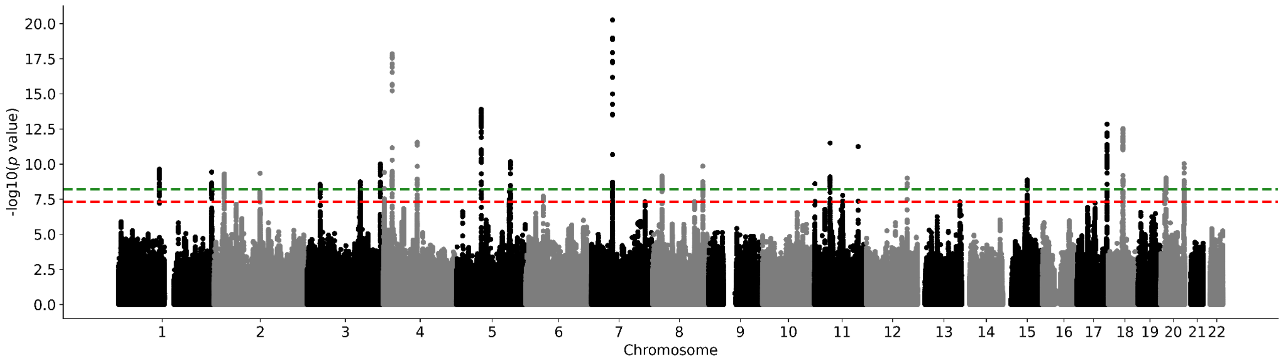

**Supplementary Figure 17. Thalamus meta-analysis Manhattan plot.** Results for thalamus GWAS. Genome-wide significance is shown for the common threshold of *p*-value < $5x10^{-8}$ (red dashed line), and also for the multiple comparisons-corrected threshold of *p*-value < $6.25x10^{-9}$ (green dashed line). The p-values referenced here correspond to a two-tailed Z-test test as implemented in MTAG.

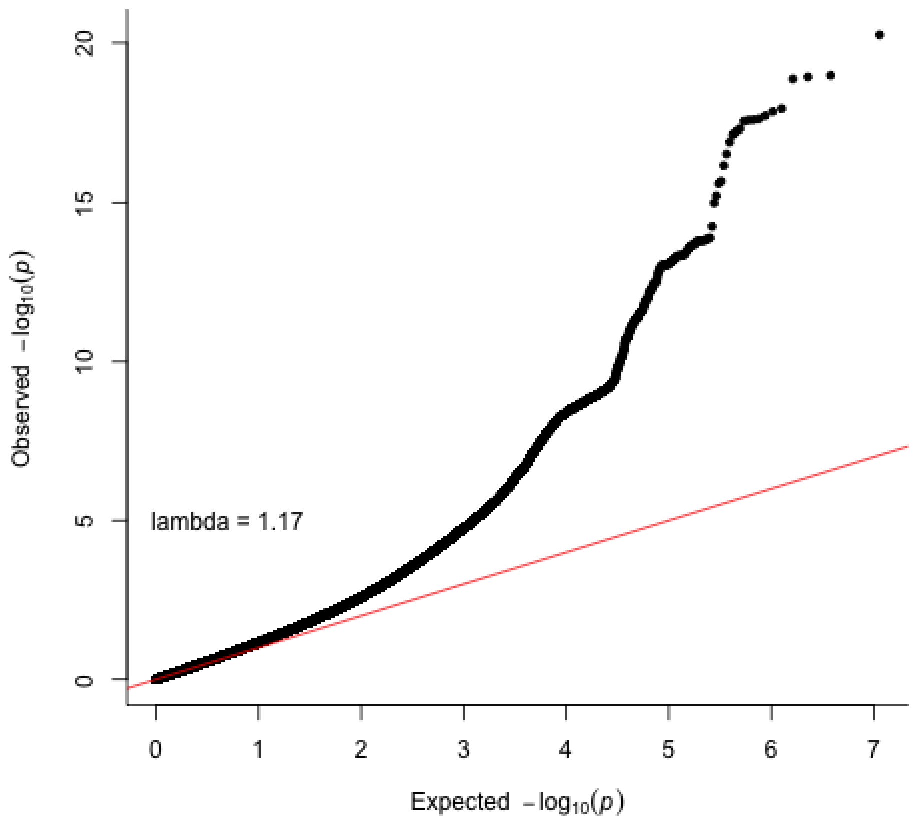

**Supplementary Figure 18. Thalamus meta-analysis QQ plot.** The p-values referenced here correspond to a two-tailed Z-test test as implemented in MTAG.

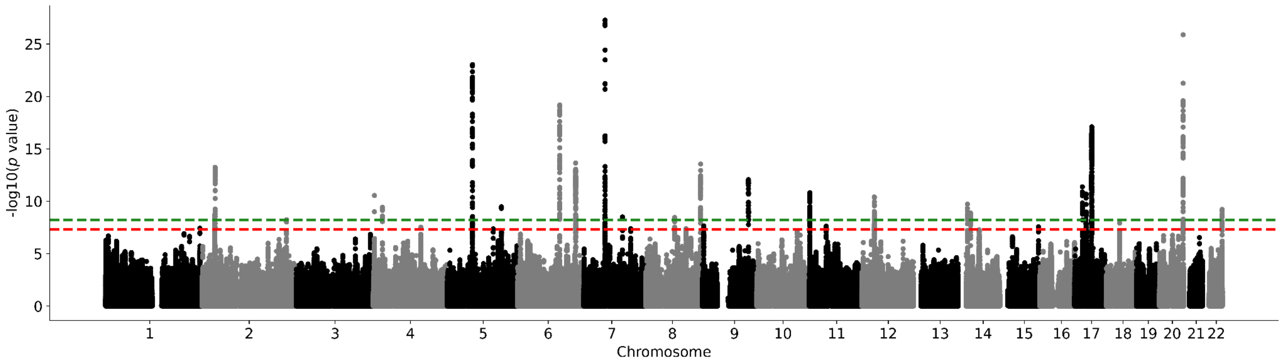

**Supplementary Figure 19. Ventral diencephalon meta-analysis Manhattan plot.** Results for ventral diencephalon GWAS. Genome-wide significance is shown for the common threshold of *p*-value < $5x10^{-8}$ (red dashed line), and also for the multiple comparisons-corrected threshold of *p*-value < $6.25x10^{-9}$ (green dashed line). The p-values referenced here correspond to a two-tailed Z-test test as implemented in MTAG.

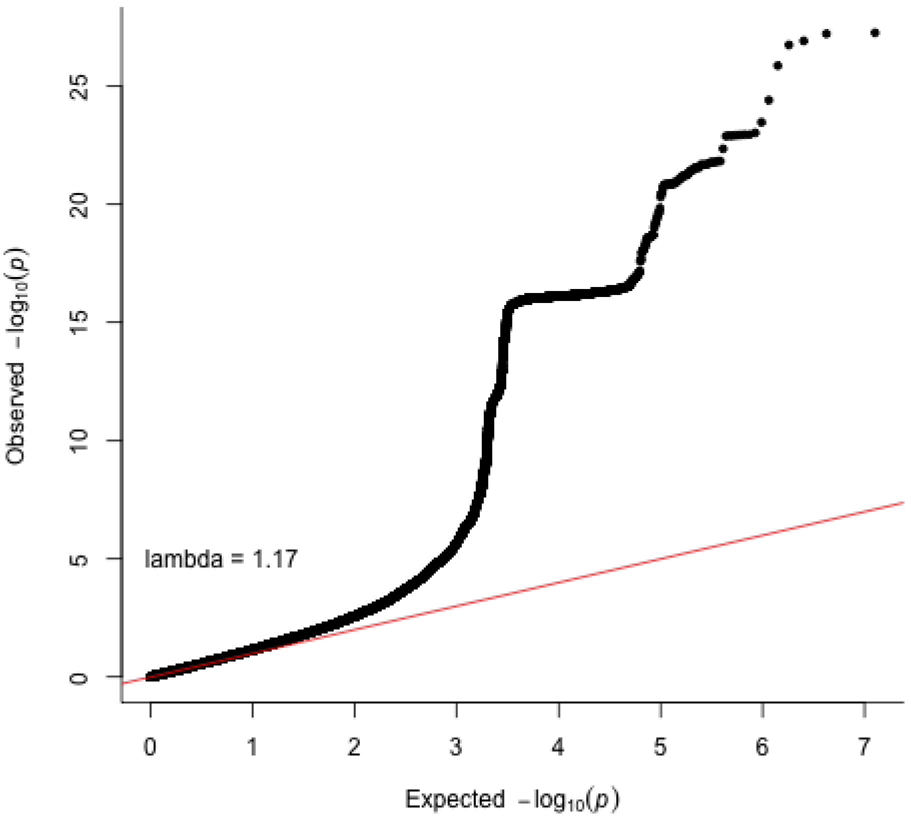

**Supplementary Figure 20. Ventral diencephalon meta-analysis QQ plot.** The p-values referenced here correspond to a two-tailed Z-test test as implemented in MTAG.

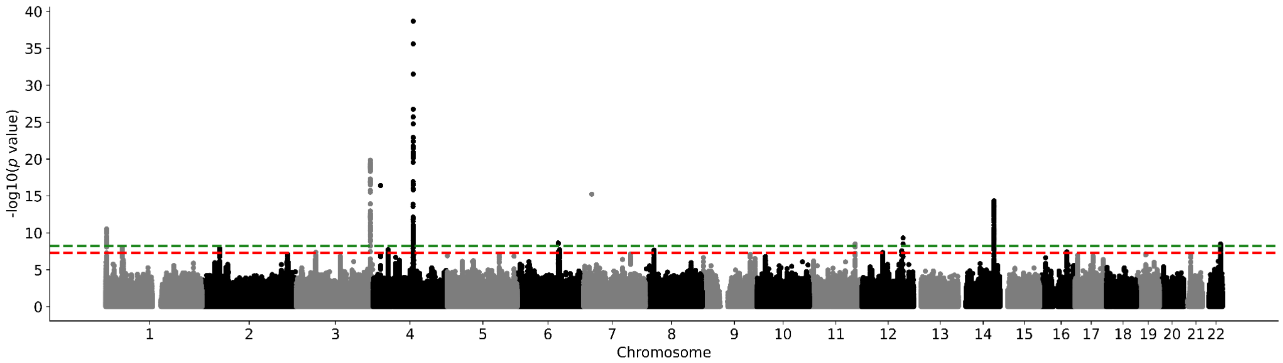

**Supplementary Figure 21. UK Biobank nucleus accumbens Manhattan plot.** Results for nucleus accumbens GWAS in the UK Biobank. Genome-wide significance is shown for the common threshold of *p*-value < $5x10^{-8}$ (red dashed line), and also for the multiple comparisons-corrected threshold of *p*-value < $6.25x10^{-9}$ (green dashed line). Two-sided P-values shown were derived from the non-infinitesimal mixed model association test p-value as implemented in BOLT-LMM.

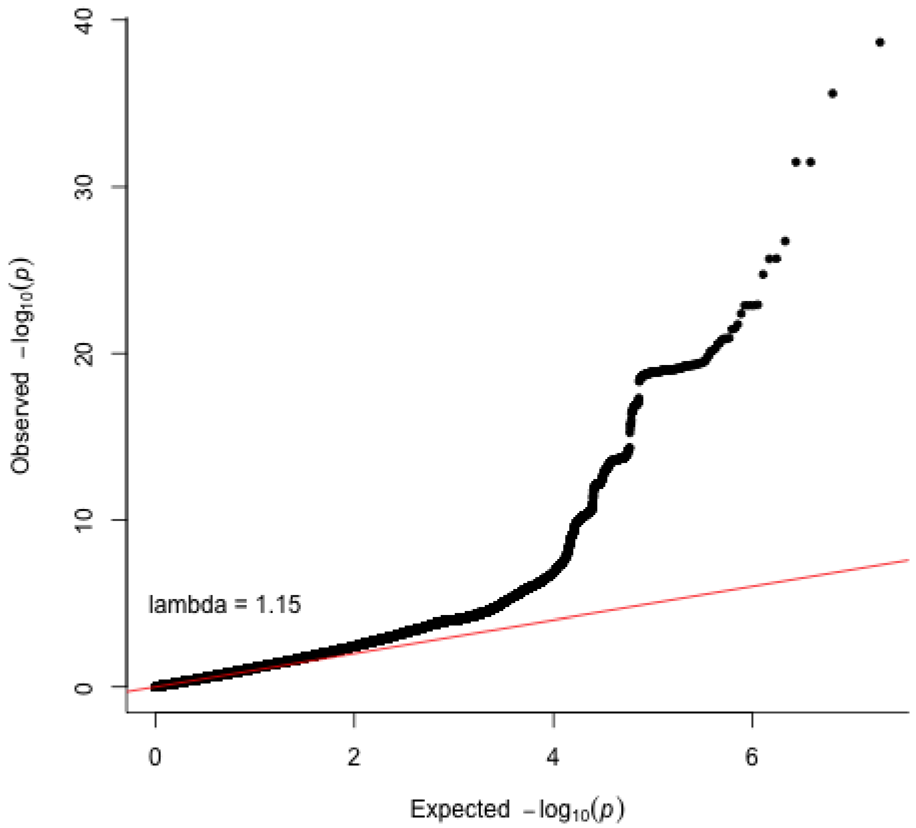

**Supplementary Figure 22. Nucleus accumbens QQ plot in the UK Biobank.** Two-sided P-values shown were derived from the non-infinitesimal mixed model association test p-value as implemented in BOLT-LMM.

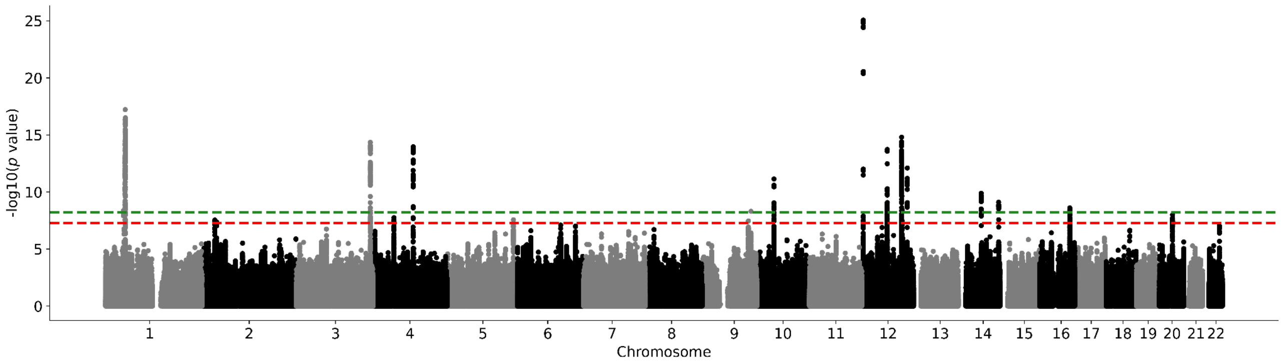

**Supplementary Figure 23. UK Biobank amygdala Manhattan plot.** Results for amygdala GWAS in the UK Biobank. Genome-wide significance is shown for the common threshold of *p*-value < $5 \times 10^{-8}$ (red dashed line), and also for the multiple comparisons-corrected threshold of *p*-value < $6.25 \times 10^{-9}$ (green dashed line). Two-sided P-values shown were derived from the non-infinitesimal mixed model association test p-value as implemented in BOLT-LMM.

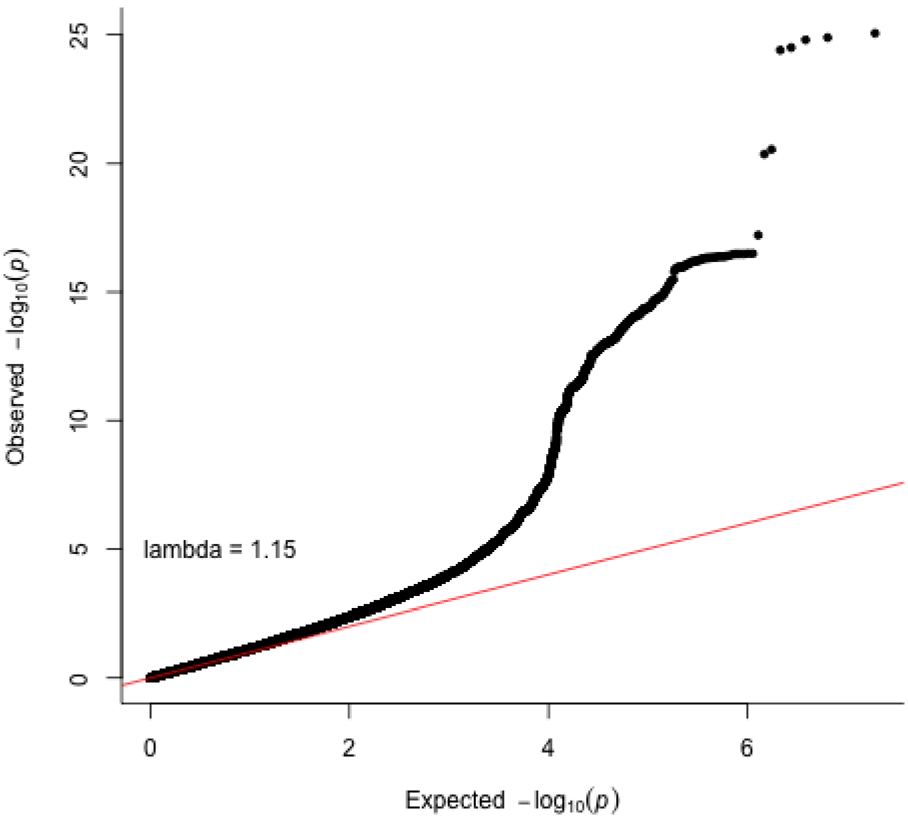

**Supplementary Figure 24. Amygdala QQ plot in the UK Biobank.** Two-sided P-values shown were derived from the non-infinitesimal mixed model association test p-value as implemented in BOLT-LMM.

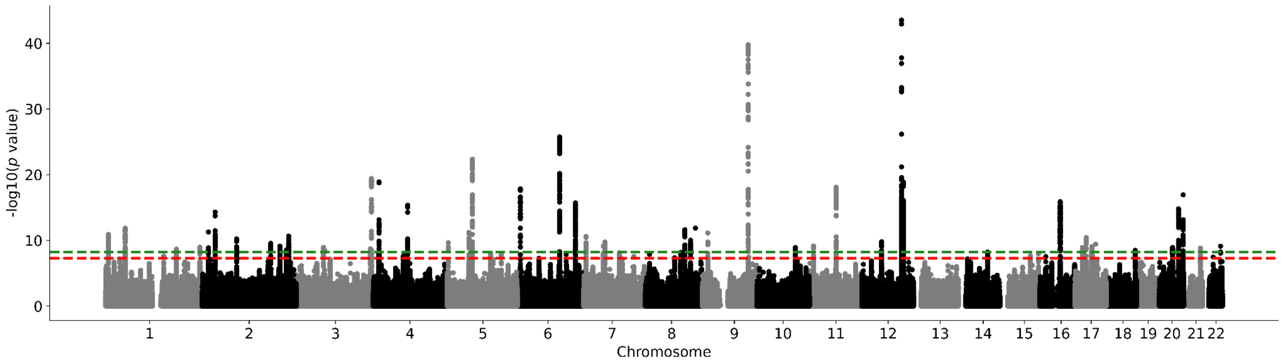

**Supplementary Figure 25. UK Biobank brainstem Manhattan plot.** Results for brainstem GWAS in the UK Biobank. Genome-wide significance is shown for the common threshold of $p$-value < 5x10$^{-8}$ (red dashed line), and also for the multiple comparisons-corrected threshold of $p$-value < 6.25x10$^{-9}$ (green dashed line). Two-sided P-values shown were derived from the non-infinitesimal mixed model association test p-value as implemented in BOLT-LMM.

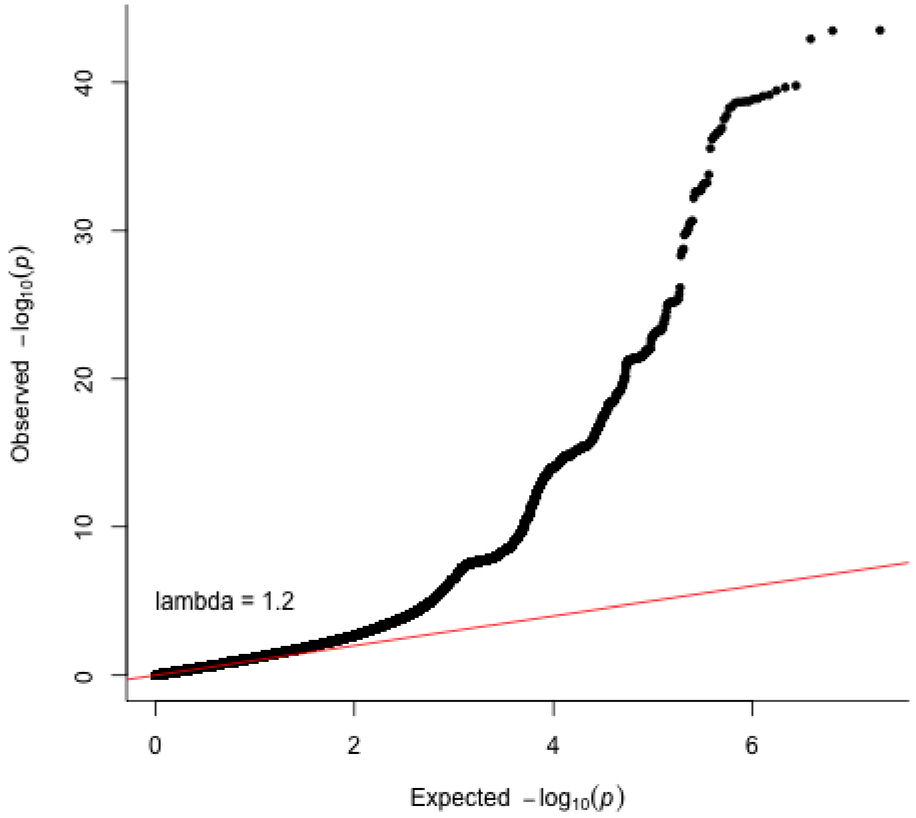

**Supplementary Figure 26. Brainstem QQ plot in the UK Biobank .** Two-sided P-values shown were derived from the non-infinitesimal mixed model association test p-value as implemented in BOLT-LMM.

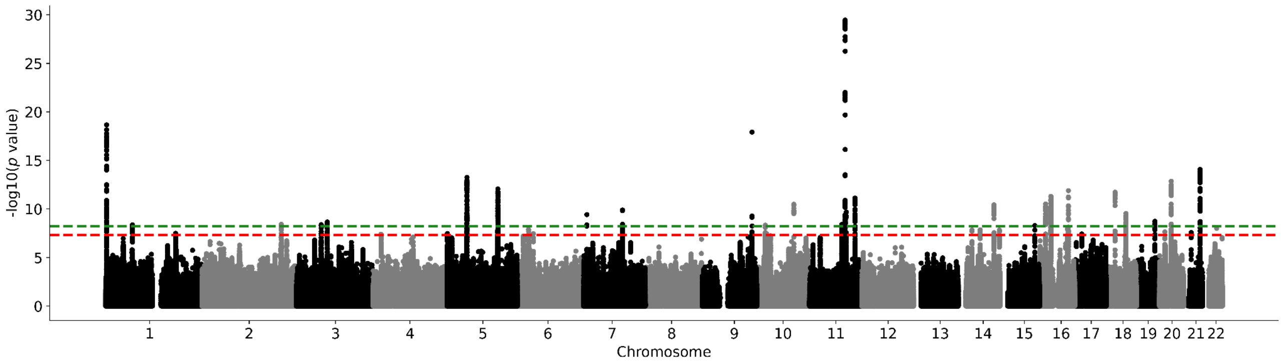

**Supplementary Figure 27. UK Biobank caudate nucleus Manhattan plot.** Results for caudate nucleus GWAS in the UK Biobank. Genome-wide significance is shown for the common threshold of *p*-value < $5x10^{-8}$ (red dashed line), and also for the multiple comparisons-corrected threshold of *p*-value < $6.25x10^{-9}$ (green dashed line). Two-sided P-values shown were derived from the non-infinitesimal mixed model association test p-value as implemented in BOLT-LMM.

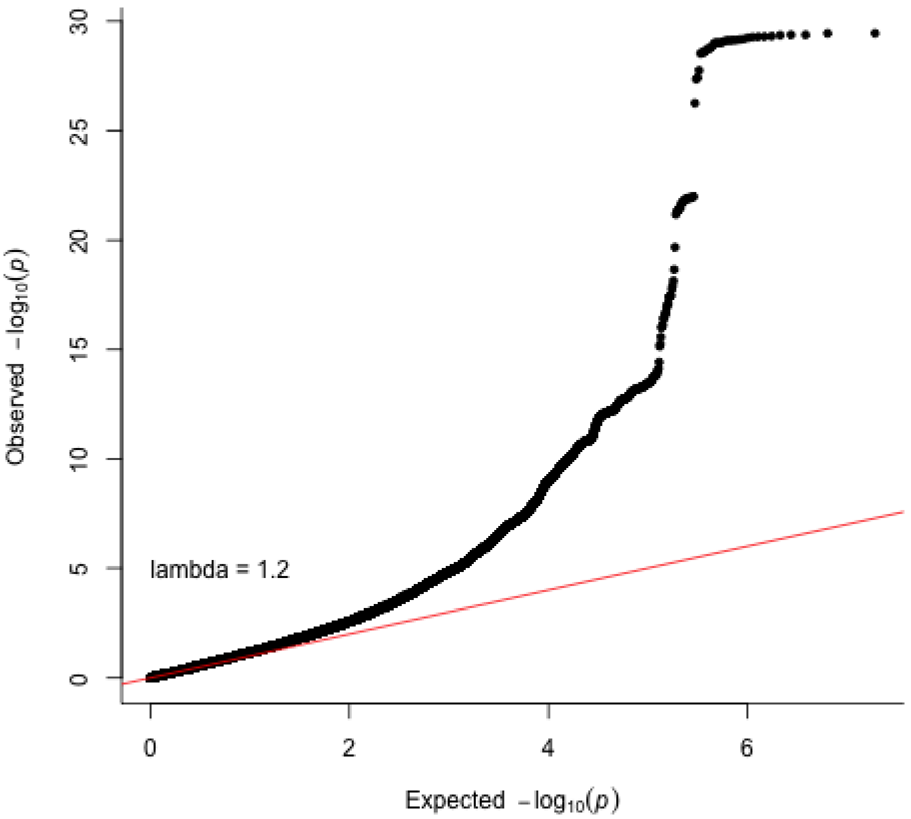

**Supplementary Figure 28. Caudate nucleus QQ plot in the UK Biobank.** Two-sided P-values shown were derived from the non-infinitesimal mixed model association test p-value as implemented in BOLT-LMM.

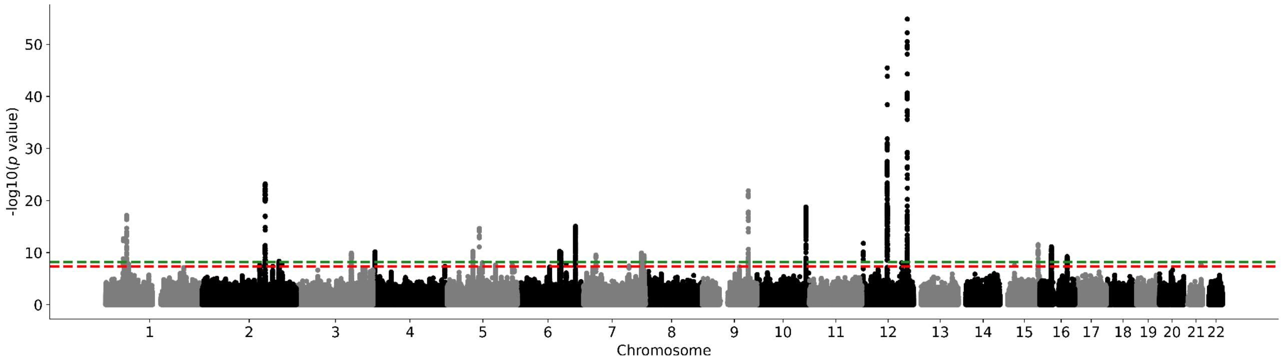

**Supplementary Figure 29. UK Biobank hippocampus Manhattan plot.** Results for hippocampus GWAS in the UK Biobank. Genome-wide significance is shown for the common threshold of *p*-value < $5 \times 10^{-8}$ (red dashed line), and also for the multiple comparisons-corrected threshold of *p*-value < $6.25 \times 10^{-9}$ (green dashed line). Two-sided P-values shown were derived from the non-infinitesimal mixed model association test p-value as implemented in BOLT-LMM.

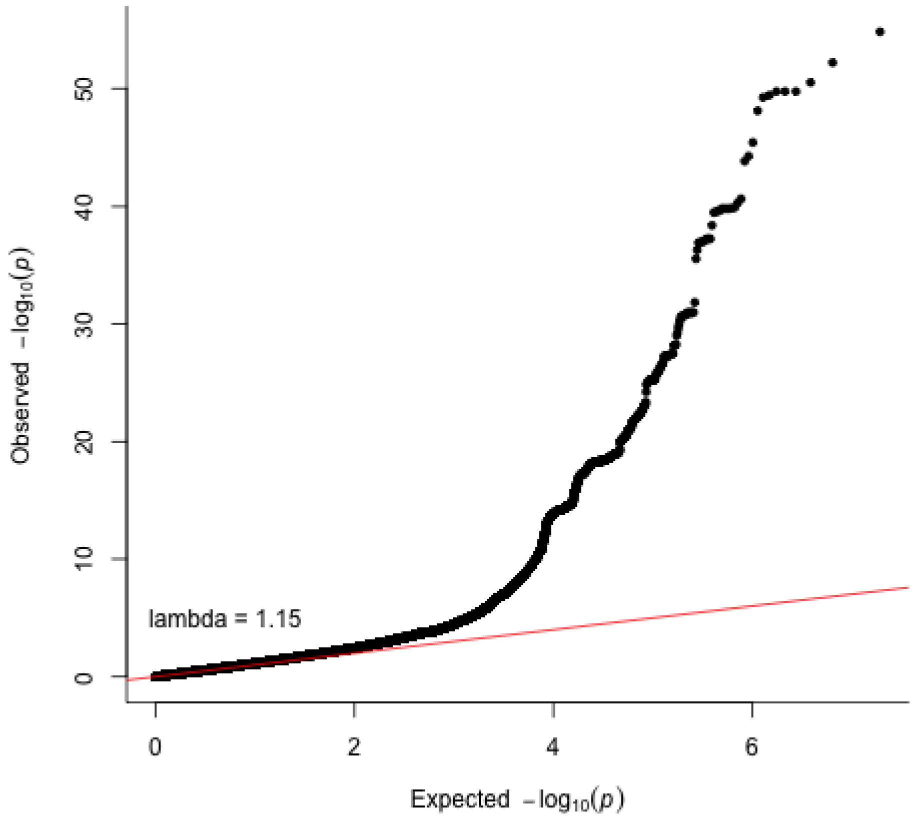

**Supplementary Figure 30. Hippocampus QQ plot in the UK Biobank.** Two-sided P-values shown were derived from the non-infinitesimal mixed model association test p-value as implemented in BOLT-LMM.

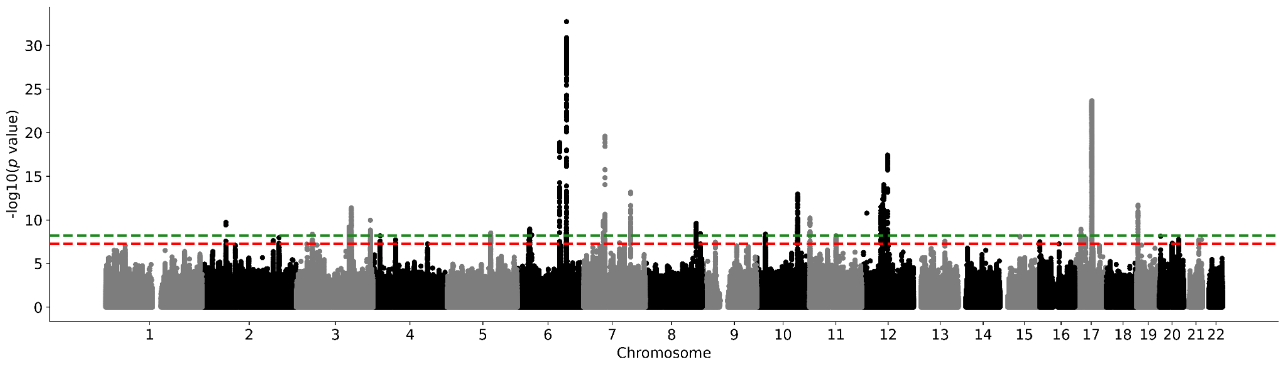

**Supplementary Figure 31. UK Biobank intracranial volume Manhattan plot.** Results for intracranial volume GWAS in the UK Biobank. Genome-wide significance is shown for the common threshold of *p*-value < $5x10^{-8}$ (red dashed line), and also for the multiple comparisons-corrected threshold of *p*-value < $6.25x10^{-9}$ (green dashed line). Two-sided P-values shown were derived from the non-infinitesimal mixed model association test p-value as implemented in BOLT-LMM.

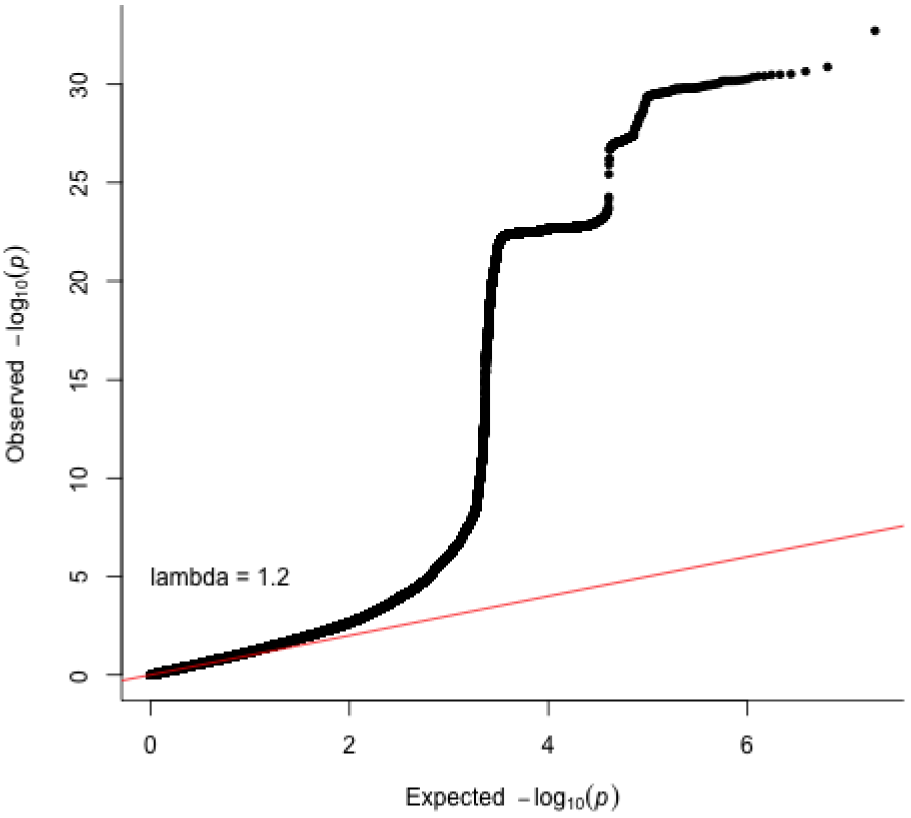

**Supplementary Figure 32. Intracranial volume QQ plot in the UK Biobank.** Two-sided P-values shown were derived from the non-infinitesimal mixed model association test p-value as implemented in BOLT-LMM.

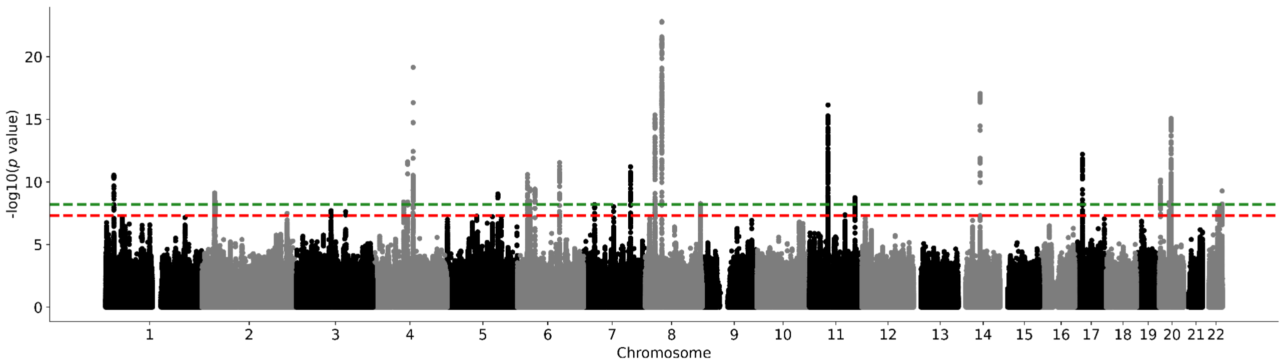

**Supplementary Figure 33. UK Biobank globus pallidus Manhattan plot.** Results for globus pallidus GWAS in the UK Biobank. Genome-wide significance is shown for the common threshold of *p*-value < $5x10^{-8}$ (red dashed line), and also for the multiple comparisons-corrected threshold of *p*-value < $6.25x10^{-9}$ (green dashed line). Two-sided P-values shown were derived from the non-infinitesimal mixed model association test p-value as implemented in BOLT-LMM.

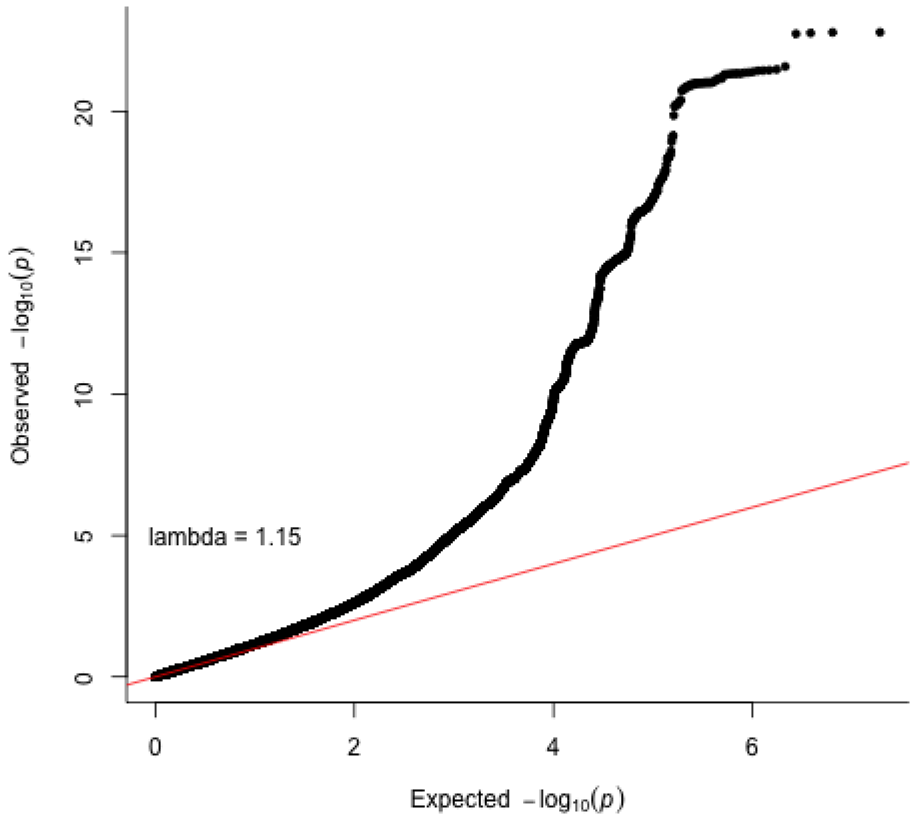

**Supplementary Figure 34. Globus pallidum QQ plot in the UK Biobank.** Two-sided P-values shown were derived from the non-infinitesimal mixed model association test p-value as implemented in BOLT-LMM.

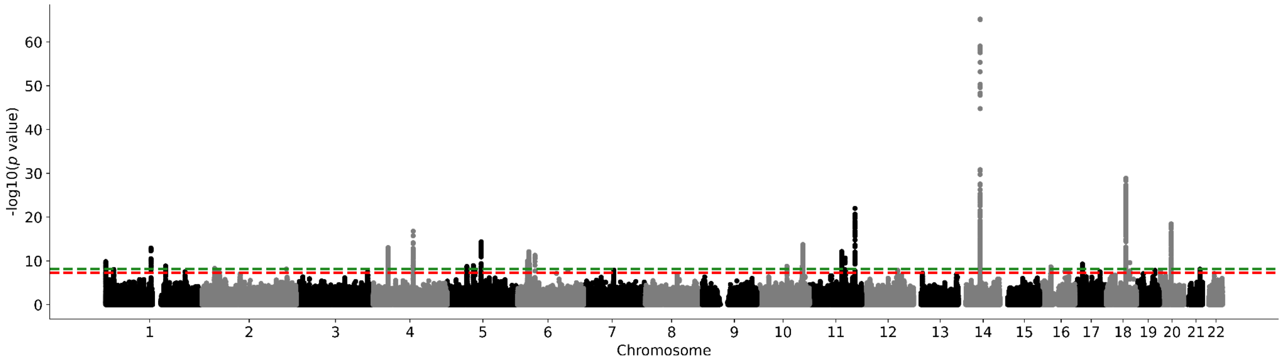

**Supplementary Figure 35. UK Biobank putamen Manhattan plot.** Results for putamen GWAS in the UK Biobank. Genome-wide significance is shown for the common threshold of *p*-value < $5x10^{-8}$ (red dashed line), and also for the multiple comparisons-corrected threshold of *p*-value < $6.25x10^{-9}$ (green dashed line). Two-sided P-values shown were derived from the non-infinitesimal mixed model association test p-value as implemented in BOLT-LMM.

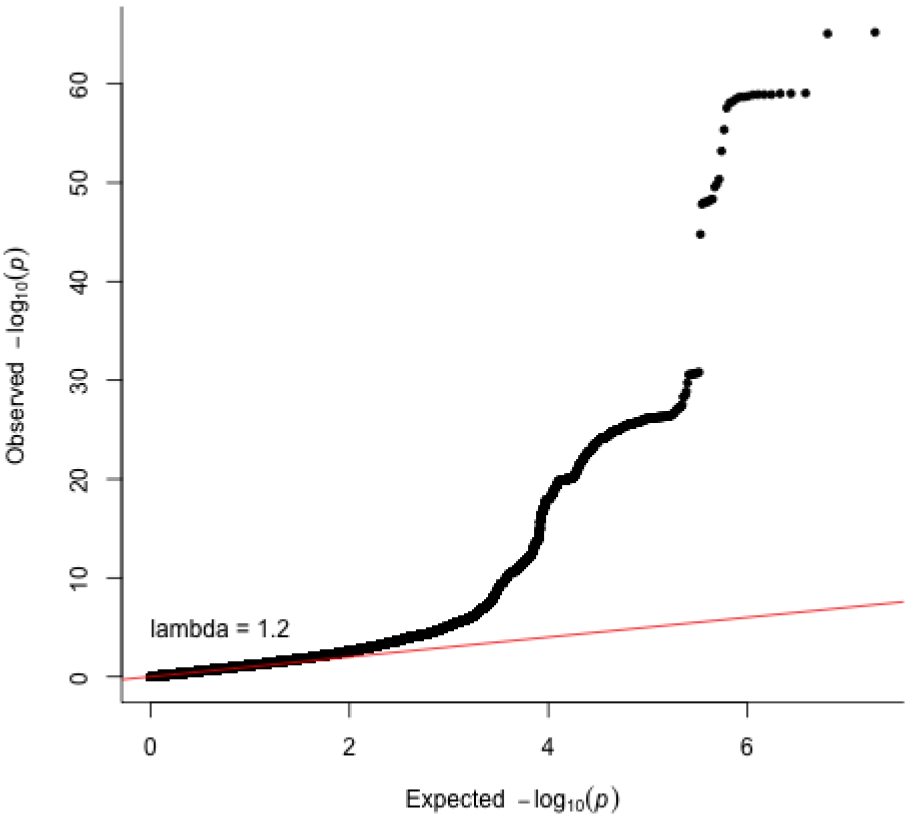

**Supplementary Figure 36. Putamen QQ plot in the UK Biobank.** Two-sided P-values shown were derived from the non-infinitesimal mixed model association test p-value as implemented in BOLT-LMM.

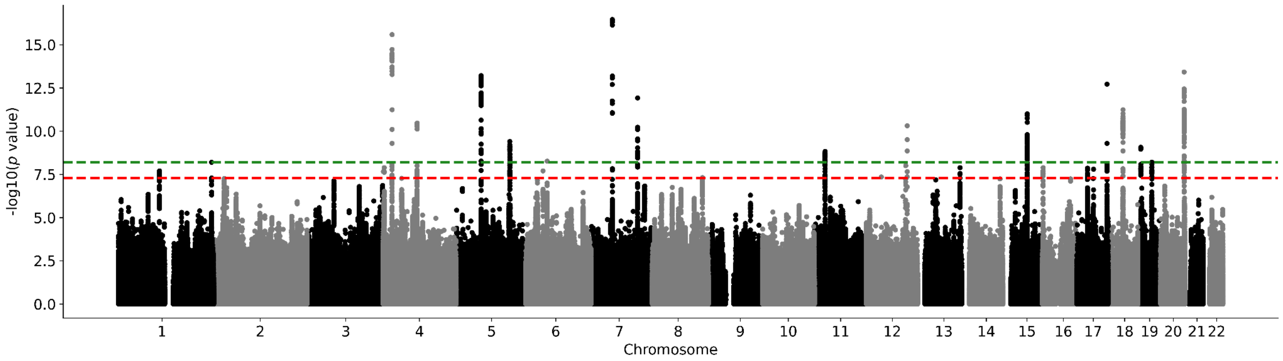

**Supplementary Figure 37. UK Biobank thalamus Manhattan plot.** Results for thalamus GWAS in the UK Biobank. Genome-wide significance is shown for the common threshold of *p*-value < $5 \times 10^{-8}$ (red dashed line), and also for the multiple comparisons-corrected threshold of *p*-value < $6.25 \times 10^{-9}$ (green dashed line). Two-sided P-values shown were derived from the non-infinitesimal mixed model association test p-value as implemented in BOLT-LMM.

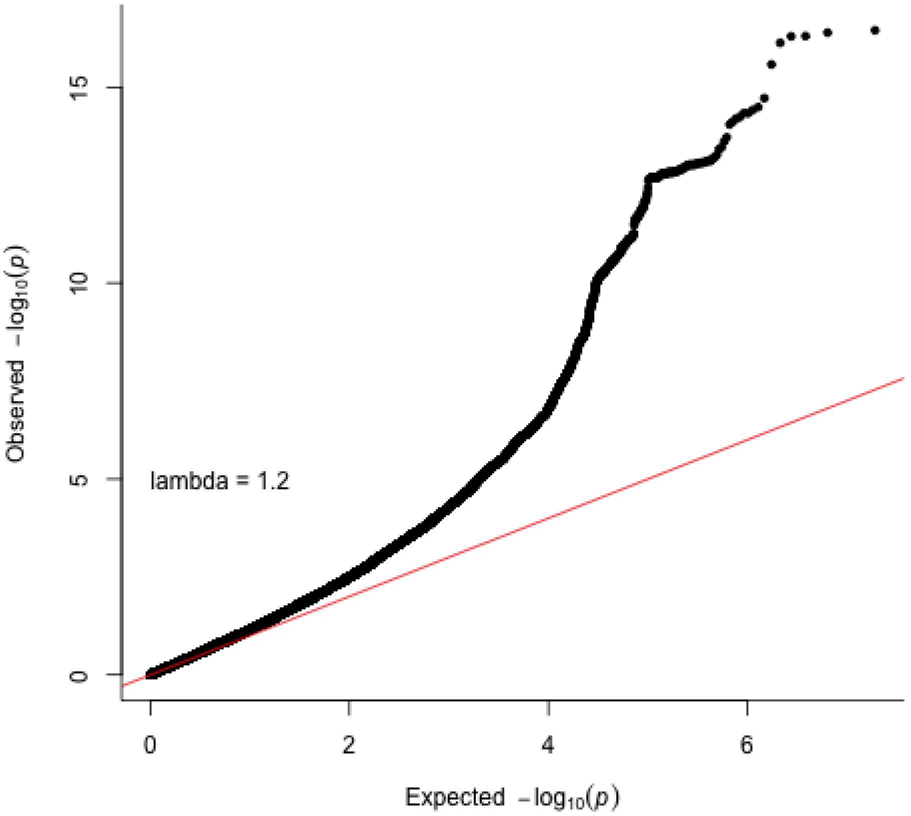

**Supplementary Figure 38. Thalamus QQ plot in the UK Biobank.** Two-sided P-values shown were derived from the non-infinitesimal mixed model association test p-value as implemented in BOLT-LMM.

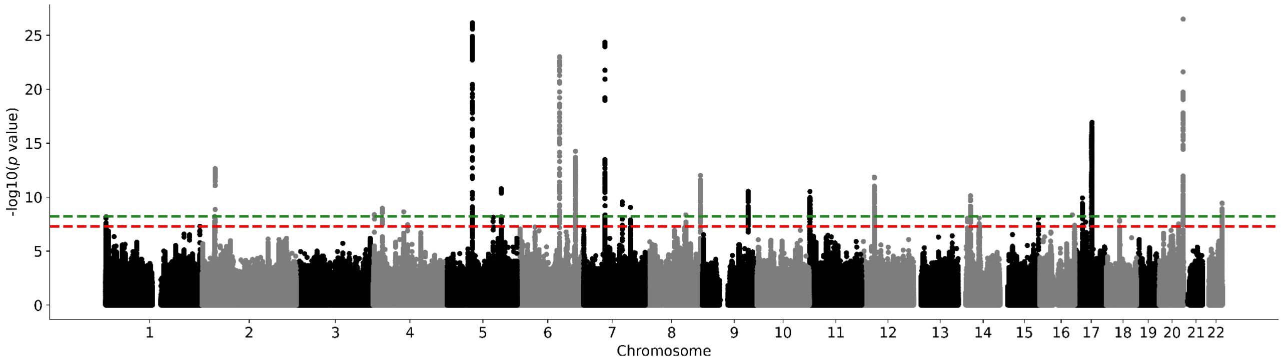

**Supplementary Figure 39. UK Biobank ventral diencephalon Manhattan plot.** Results for ventral diencephalon GWAS in the UK Biobank. Genome-wide significance is shown for the common threshold of *p*-value < $5x10^{-8}$ (red dashed line), and also for the multiple comparisons-corrected threshold of *p*-value < $6.25x10^{-9}$ (green dashed line). Two-sided P-values shown were derived from the non-infinitesimal mixed model association test p-value as implemented in BOLT-LMM.

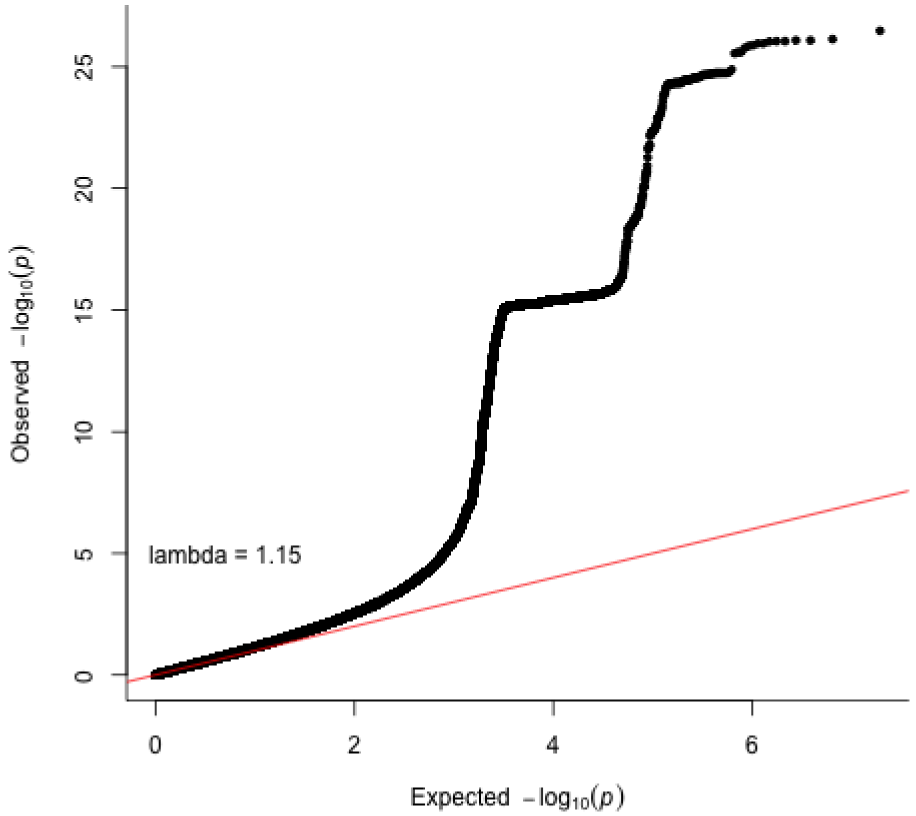

**Supplementary Figure 40. Ventral diencephalon QQ plot in the UK Biobank.** Two-sided P-values shown were derived from the non-infinitesimal mixed model association test p-value as implemented in BOLT-LMM.

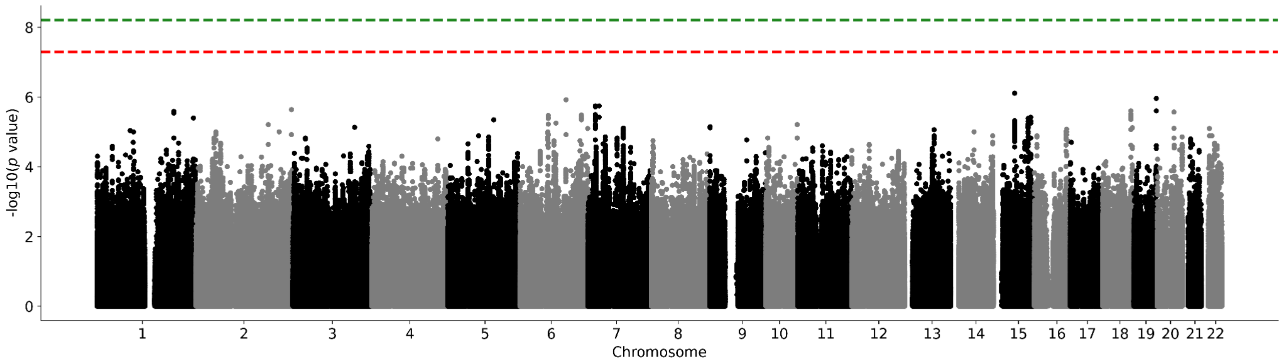

**Supplementary Figure 41. ABCD cohort nucleus accumbens Manhattan plot.** Results for nucleus accumbens GWAS in the ABCD cohort. Genome-wide significance is shown for the common threshold of *p*-value < 5x10$^{-8}$ (red dashed line), and also for the multiple comparisons-corrected threshold of *p*-value < 6.25x10$^{-9}$ (green dashed line). Two-sided P-values shown were derived from the non-infinitesimal mixed model association test p-value as implemented in BOLT-LMM.

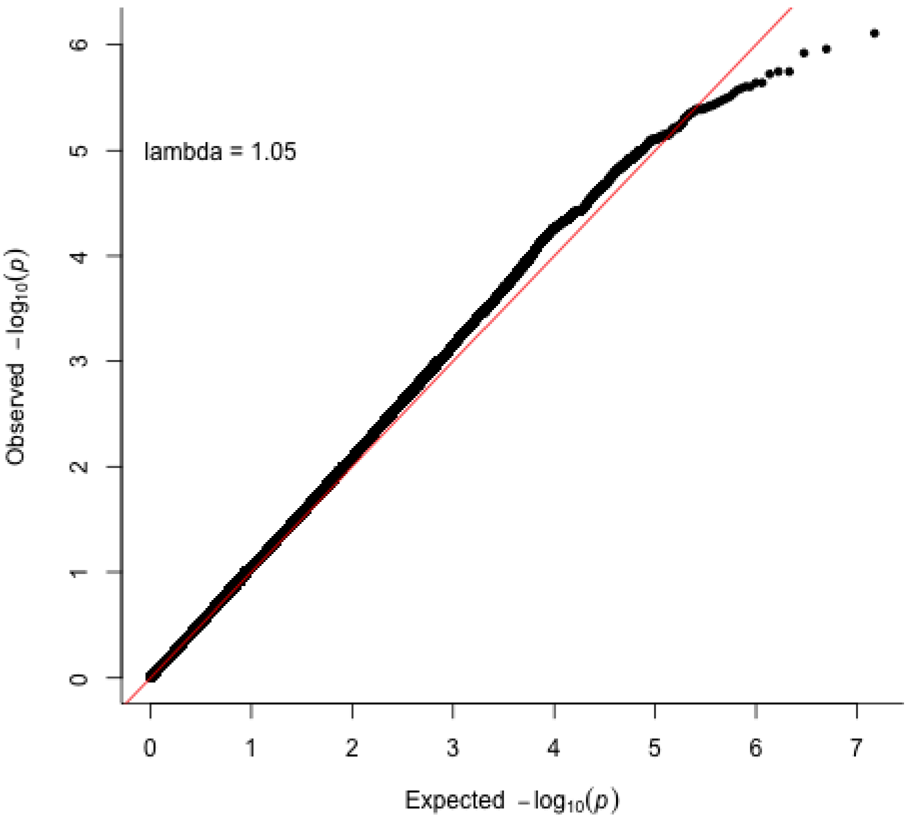

**Supplementary Figure 42. Nucleus accumbens QQ plot in the ABCD cohort.** Two-sided P-values shown were derived from the non-infinitesimal mixed model association test p-value as implemented in BOLT-LMM.

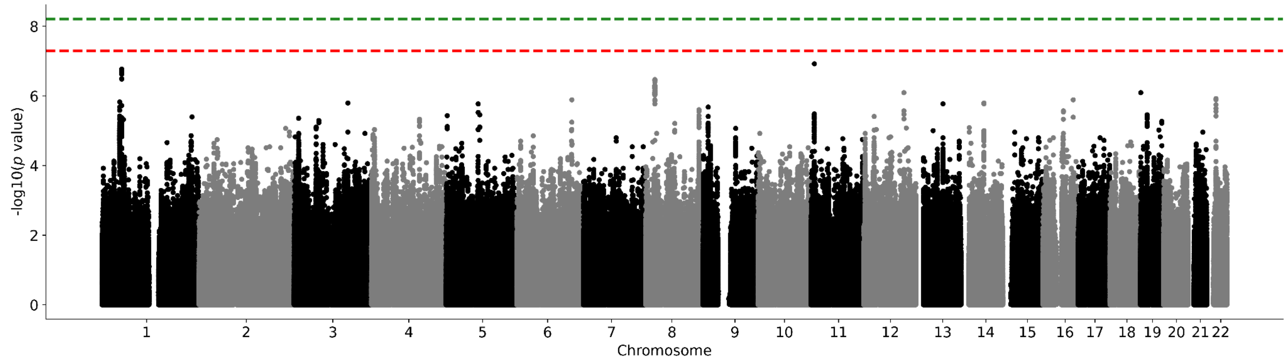

**Supplementary Figure 43. ABCD cohort amygdala Manhattan plot.** Results for amygdala GWAS in the ABCD cohort. Genome-wide significance is shown for the common threshold of *p*-value < $5x10^{-8}$ (red dashed line), and also for the multiple comparisons-corrected threshold of *p*-value < $6.25x10^{-9}$ (green dashed line). Two-sided P-values shown were derived from the non-infinitesimal mixed model association test p-value as implemented in BOLT-LMM.

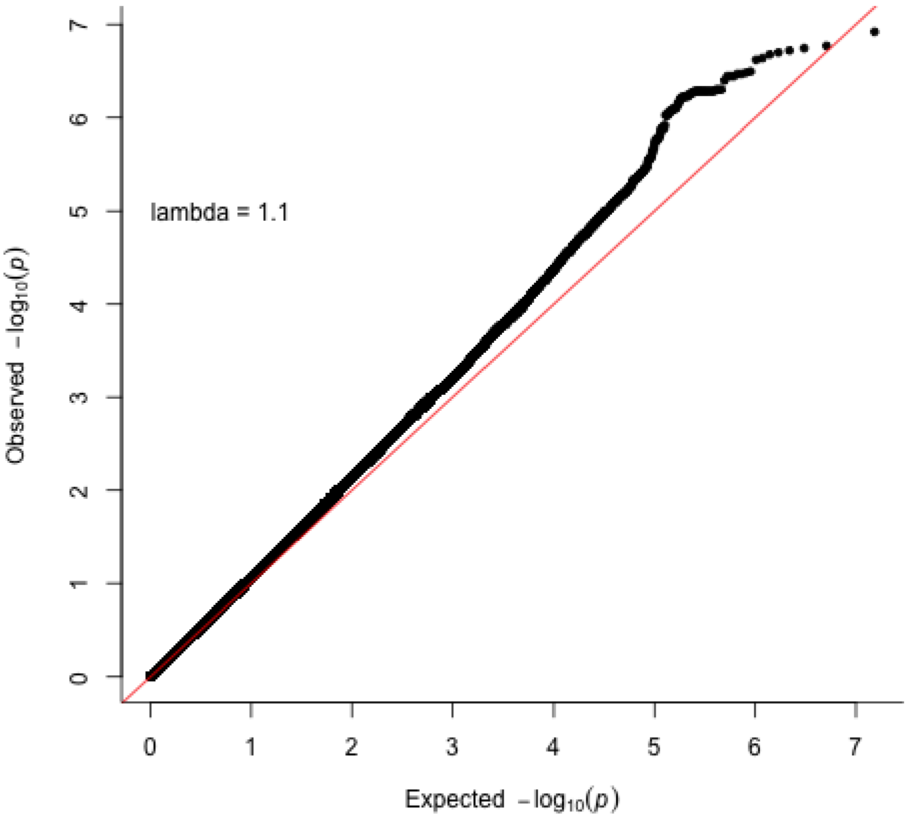

**Supplementary Figure 44. Amygdala QQ plot in the ABCD cohort.** Two-sided P-values shown were derived from the non-infinitesimal mixed model association test p-value as implemented in BOLT-LMM.

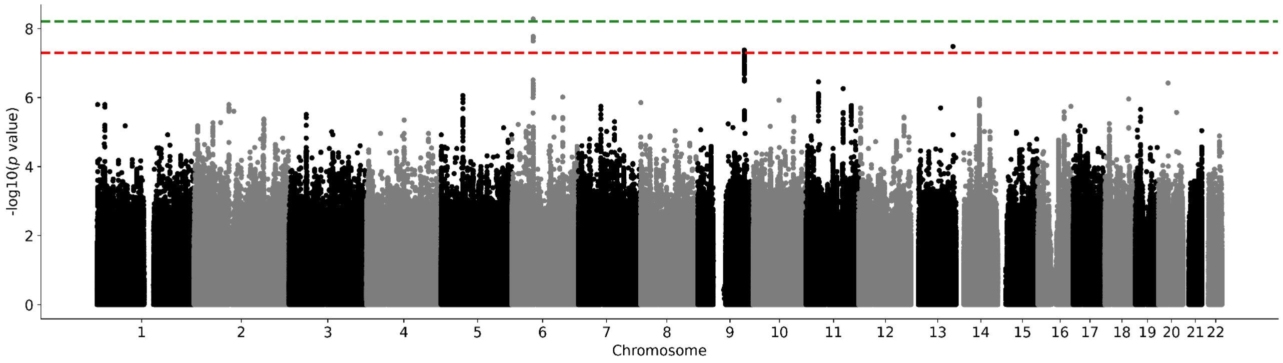

**Supplementary Figure 45. ABCD cohort brainstem Manhattan plot.** Results for brainstem GWAS in the ABCD cohort. Genome-wide significance is shown for the common threshold of $p$-value < $5x10^{-8}$ (red dashed line), and also for the multiple comparisons-corrected threshold of $p$-value < $6.25x10^{-9}$ (green dashed line). Two-sided P-values shown were derived from the non-infinitesimal mixed model association test p-value as implemented in BOLT-LMM.

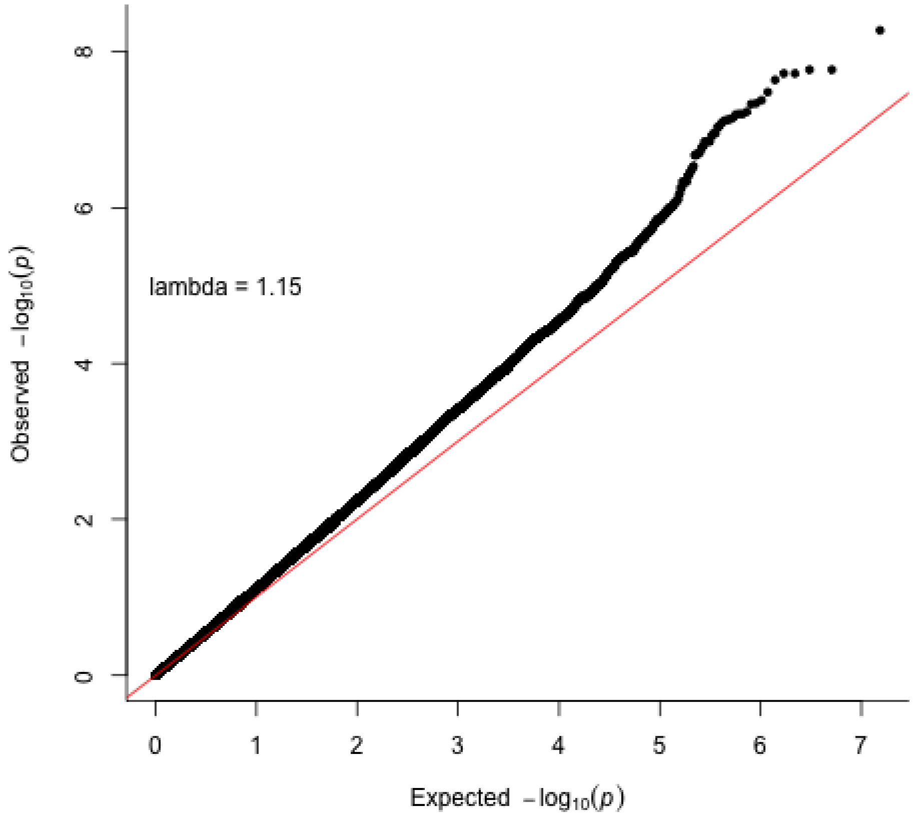

**Supplementary Figure 46. Brainstem QQ plot in the ABCD cohort.** Two-sided P-values shown were derived from the non-infinitesimal mixed model association test p-value as implemented in BOLT-LMM.

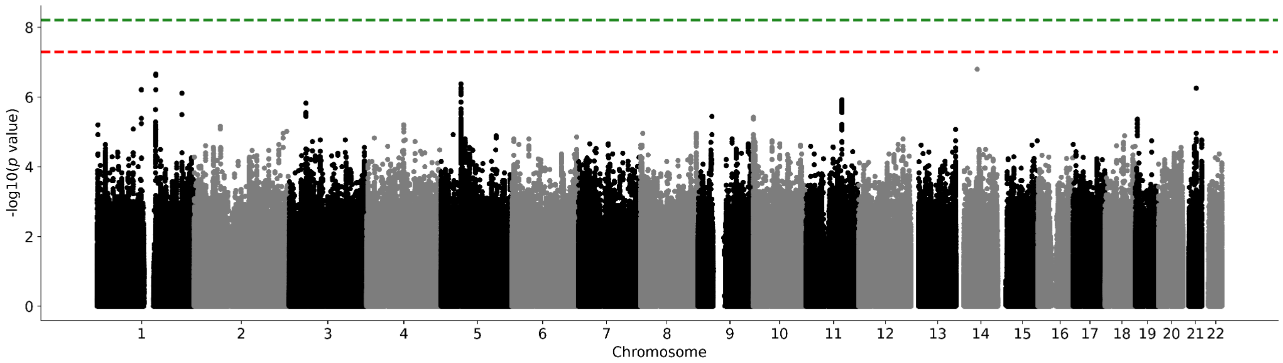

**Supplementary Figure 47. ABCD cohort caudate nucleus Manhattan plot.** Results for caudate nucleus GWAS in the ABCD cohort. Genome-wide significance is shown for the common threshold of *p*-value < $5x10^{-8}$ (red dashed line), and also for the multiple comparisons-corrected threshold of *p*-value < $6.25x10^{-9}$ (green dashed line). Two-sided P-values shown were derived from the non-infinitesimal mixed model association test p-value as implemented in BOLT-LMM.

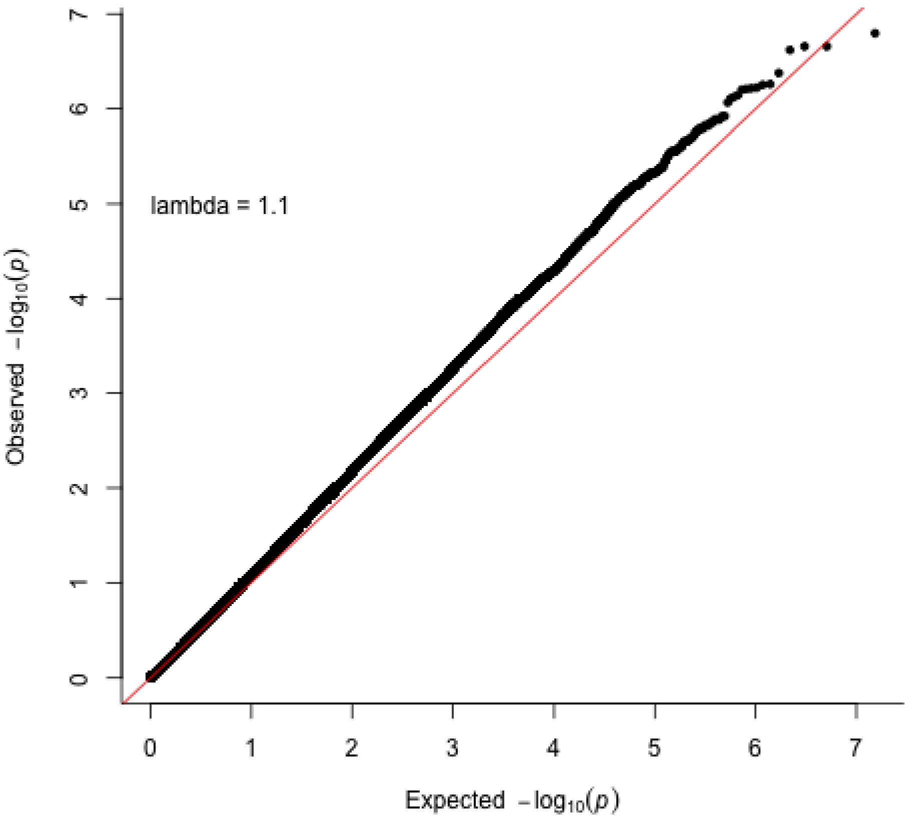

**Supplementary Figure 48. Caudate nucleus QQ plot in the ABCD cohort.** Two-sided P-values shown were derived from the non-infinitesimal mixed model association test p-value as implemented in BOLT-LMM.

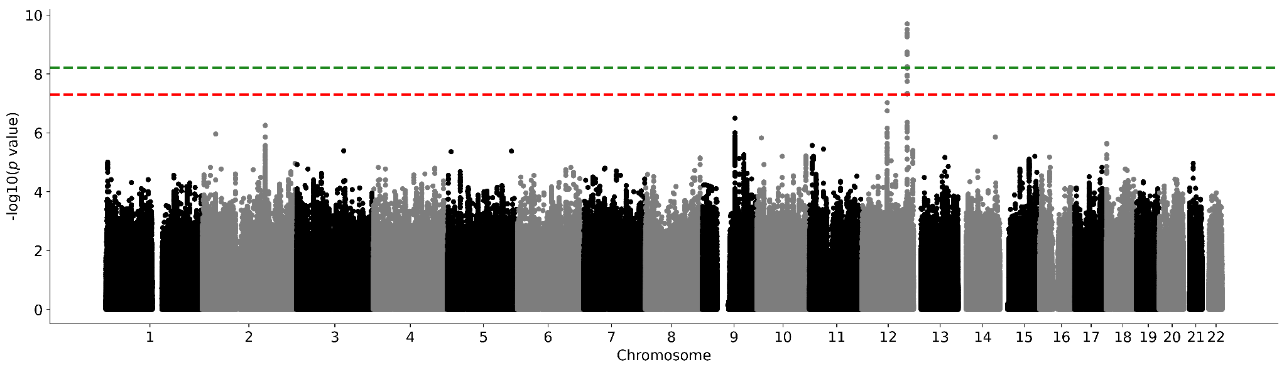

**Supplementary Figure 49. ABCD cohort hippocampus Manhattan plot.** Results for hippocampus GWAS in the ABCD cohort. Genome-wide significance is shown for the common threshold of *p*-value < $5x10^{-8}$ (red dashed line), and also for the multiple comparisons-corrected threshold of *p*-value < $6.25x10^{-9}$ (green dashed line). Two-sided P-values shown were derived from the non-infinitesimal mixed model association test p-value as implemented in BOLT-LMM.

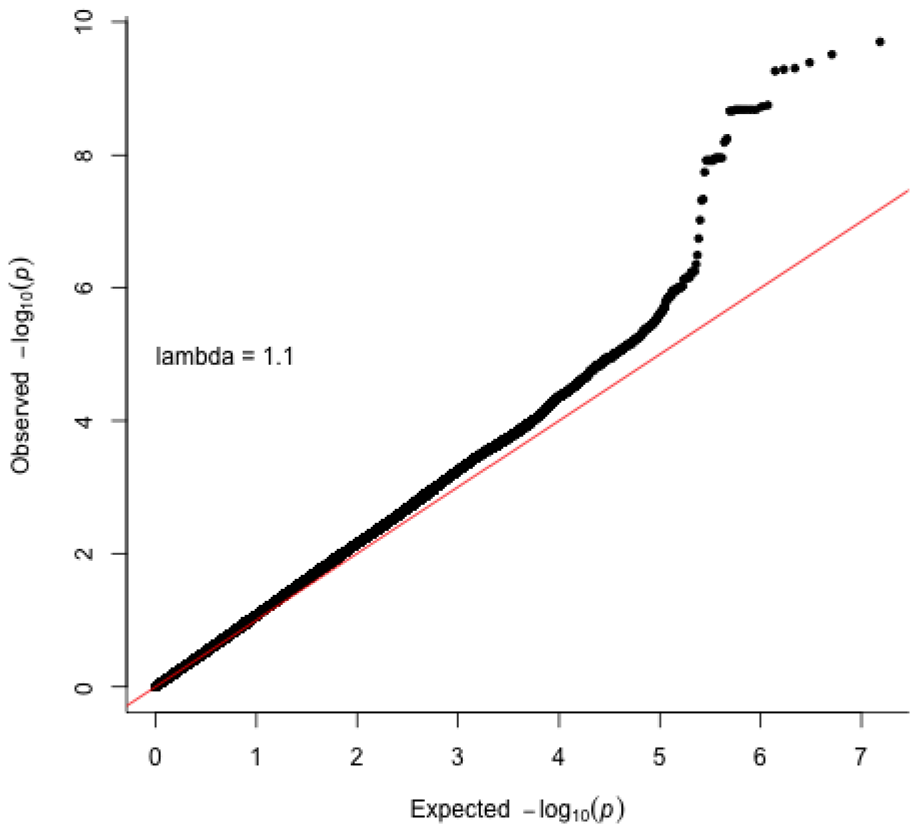

**Supplementary Figure 50. Hippocampus QQ plot in the ABCD cohort.** Two-sided P-values shown were derived from the non-infinitesimal mixed model association test p-value as implemented in BOLT-LMM.

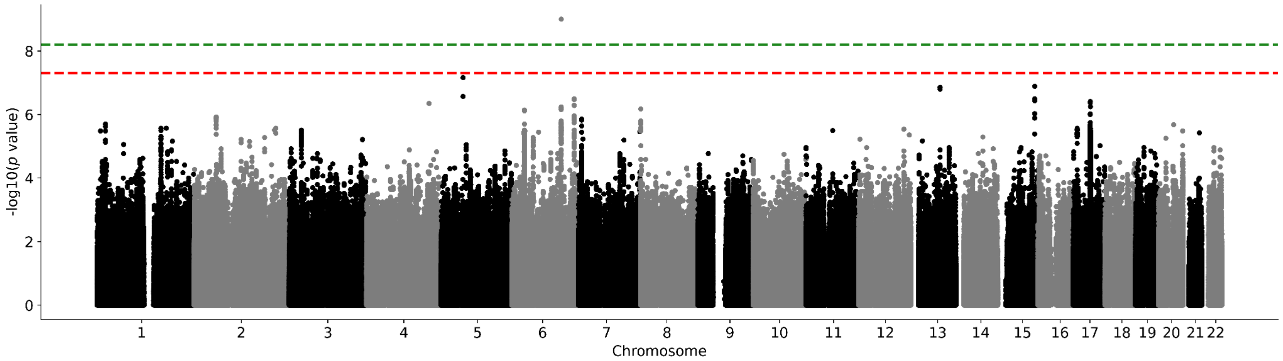

**Supplementary Figure 51. ABCD cohort intracranial volume Manhattan plot.** Results for intracranial volume GWAS in the ABCD cohort. Genome-wide significance is shown for the common threshold of *p*-value < $5x10^{-8}$ (red dashed line), and also for the multiple comparisons-corrected threshold of *p*-value < $6.25x10^{-9}$ (green dashed line). Two-sided P-values shown were derived from the non-infinitesimal mixed model association test p-value as implemented in BOLT-LMM.

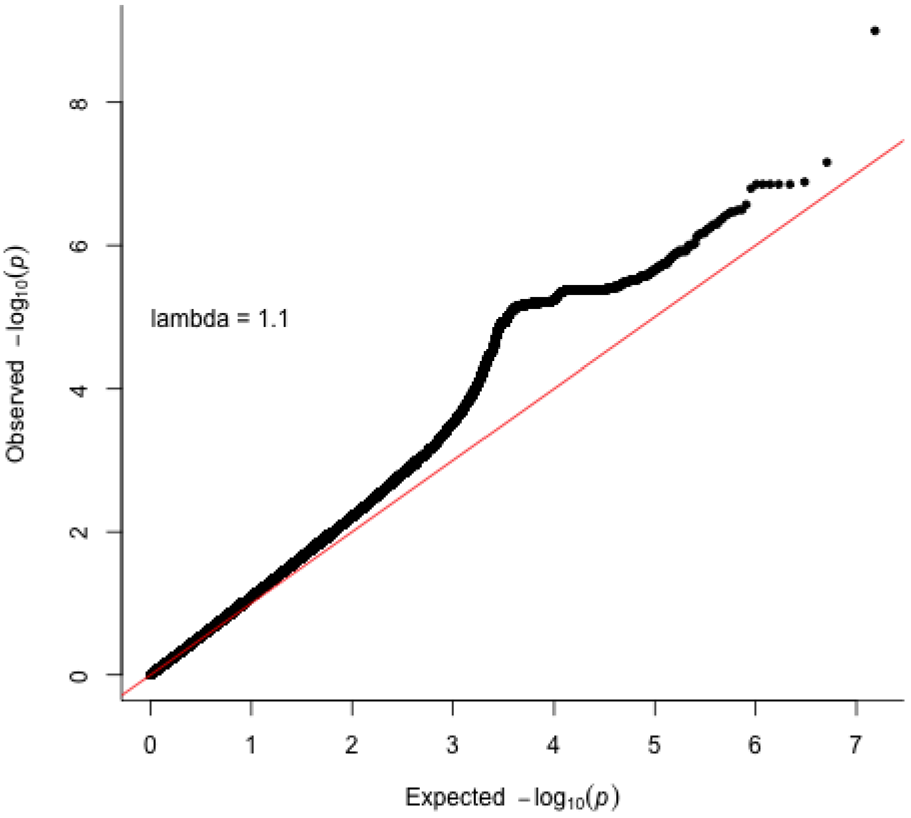

**Supplementary Figure 52. Intracranial volume QQ plot in the ABCD cohort.** Two-sided P-values shown were derived from the non-infinitesimal mixed model association test p-value as implemented in BOLT-LMM.

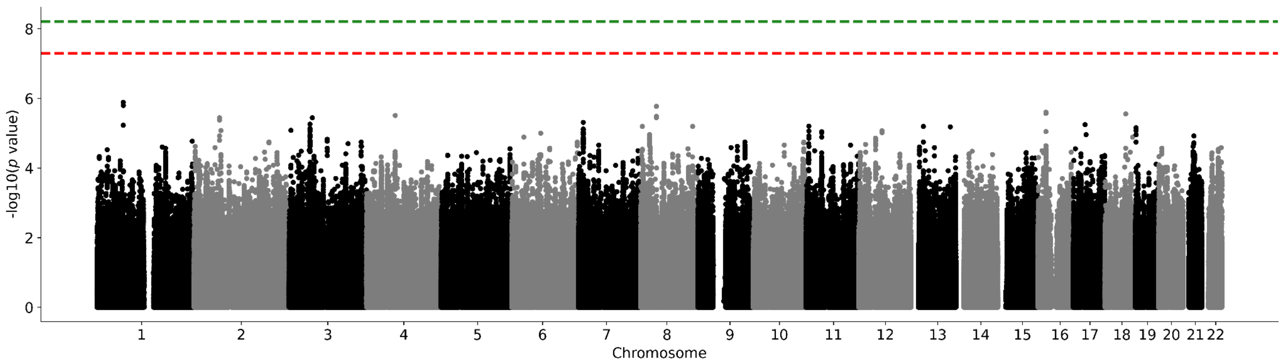

**Supplementary Figure 53. ABCD cohort globus pallidus Manhattan plot.** Results for globus pallidus GWAS in the ABCD cohort. Genome-wide significance is shown for the common threshold of *p*-value < $5x10^{-8}$ (red dashed line), and also for the multiple comparisons-corrected threshold of *p*-value < $6.25x10^{-9}$ (green dashed line). Two-sided P-values shown were derived from the non-infinitesimal mixed model association test p-value as implemented in BOLT-LMM.

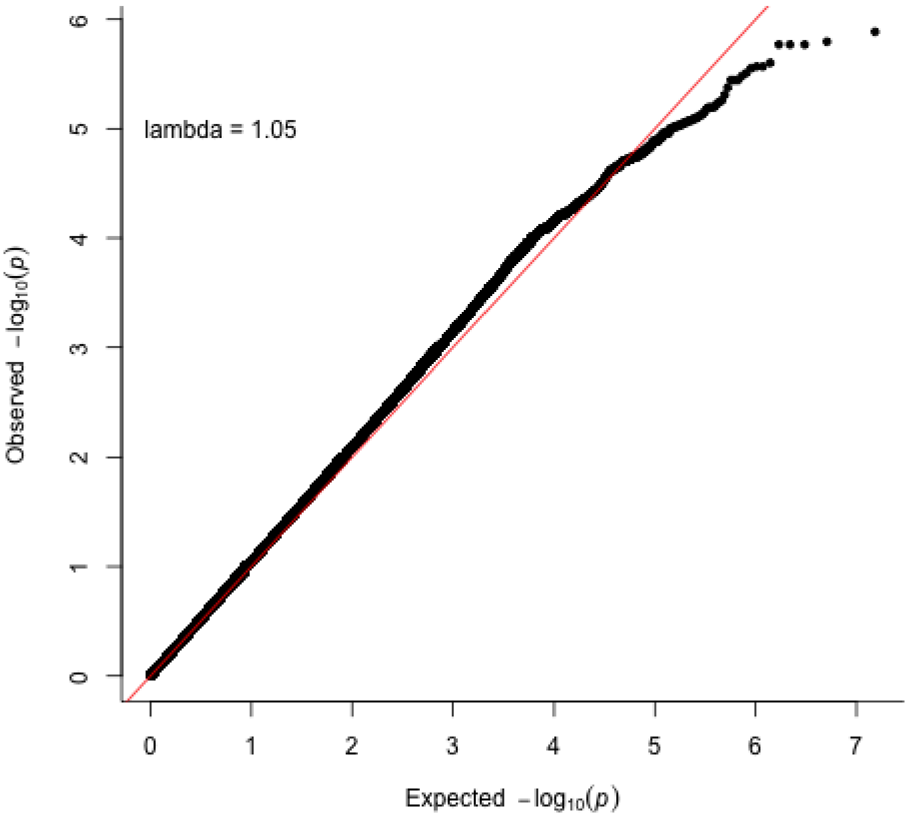

**Supplementary Figure 54. Globus pallidum QQ plot in the ABCD cohort.** Two-sided P-values shown were derived from the non-infinitesimal mixed model association test p-value as implemented in BOLT-LMM.

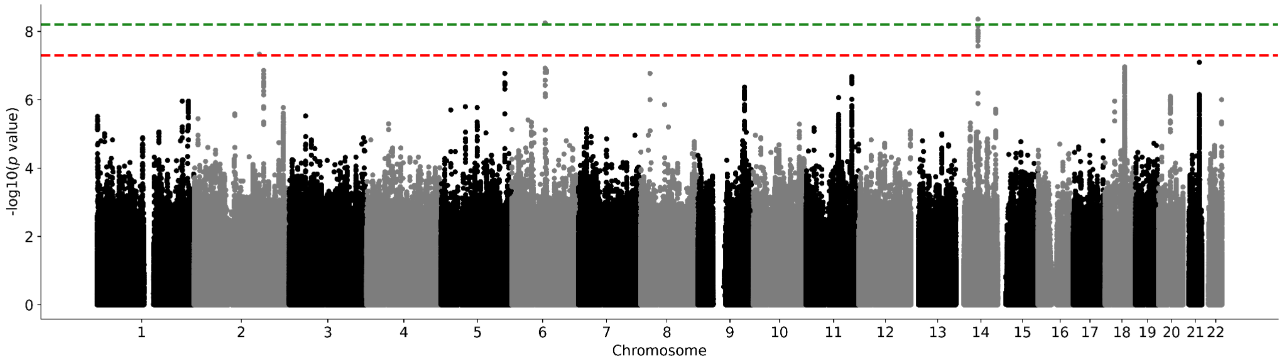

**Supplementary Figure 55. ABCD cohort putamen Manhattan plot.** Results for putamen GWAS in the ABCD cohort. Genome-wide significance is shown for the common threshold of *p*-value < $5x10^{-8}$ (red dashed line), and also for the multiple comparisons-corrected threshold of *p*-value < $6.25x10^{-9}$ (green dashed line). Two-sided P-values shown were derived from the non-infinitesimal mixed model association test p-value as implemented in BOLT-LMM.

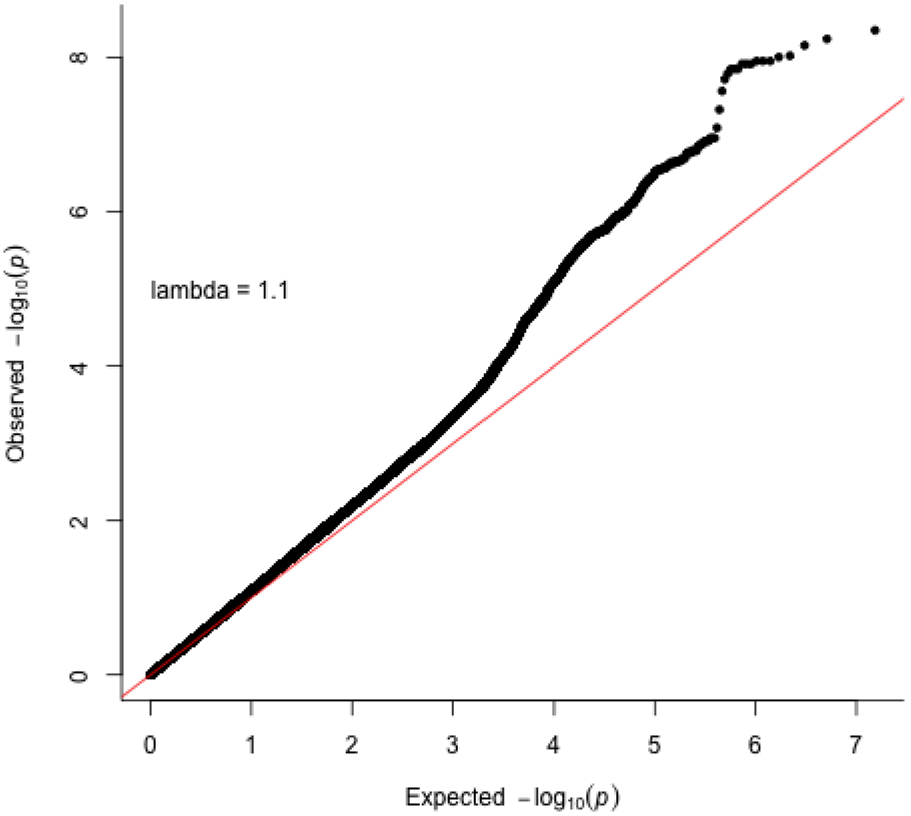

**Supplementary Figure 56. Putamen QQ plot in the ABCD cohort.** Two-sided P-values shown were derived from the non-infinitesimal mixed model association test p-value as implemented in BOLT-LMM.

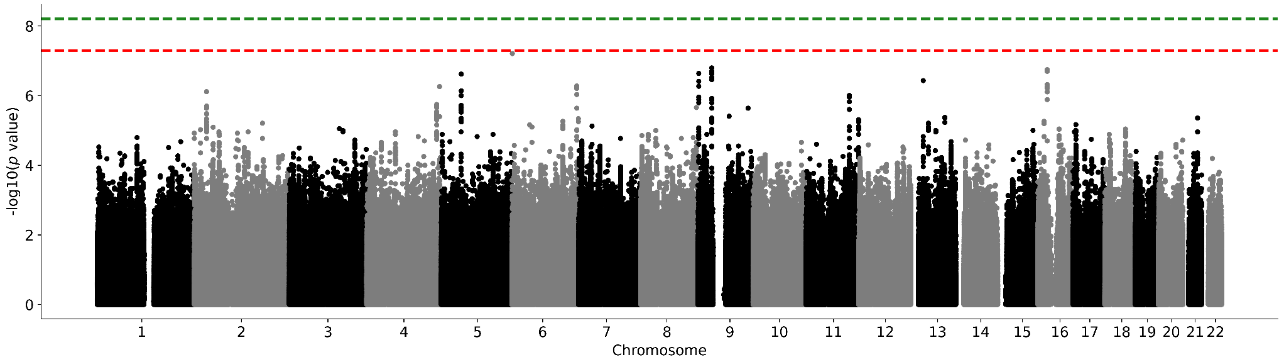

**Supplementary Figure 57. ABCD cohort thalamus Manhattan plot.** Results for thalamus GWAS in the ABCD cohort. Genome-wide significance is shown for the common threshold of *p*-value < $5x10^{-8}$ (red dashed line), and also for the multiple comparisons-corrected threshold of *p*-value < $6.25x10^{-9}$ (green dashed line). Two-sided P-values shown were derived from the non-infinitesimal mixed model association test p-value as implemented in BOLT-LMM.

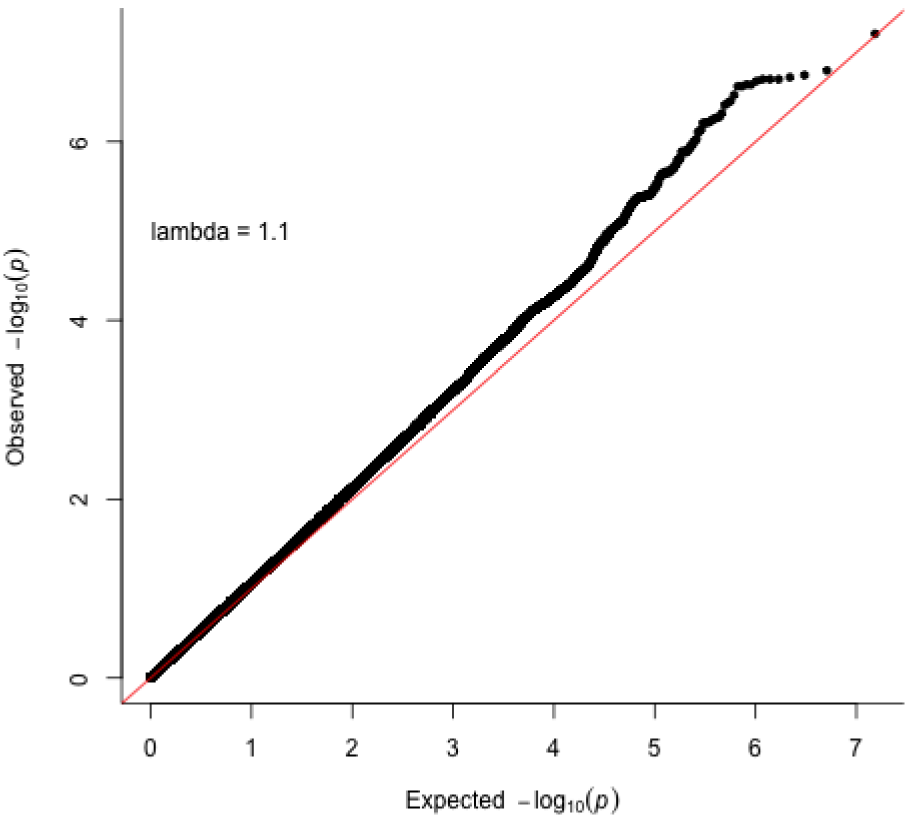

**Supplementary Figure 58. Thalamus QQ plot in the ABCD cohort.** Two-sided P-values shown were derived from the non-infinitesimal mixed model association test p-value as implemented in BOLT-LMM.

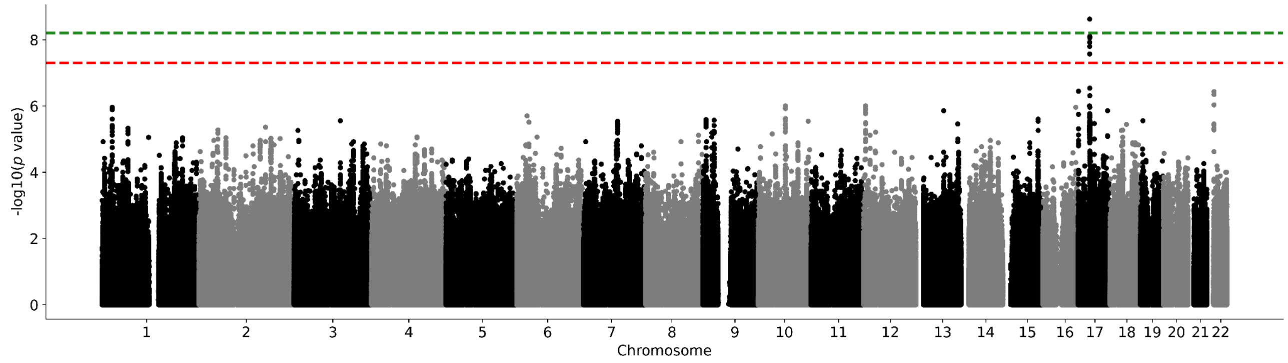

**Supplementary Figure 59. ABCD cohort ventral diencephalon Manhattan plot.** Results for ventral diencephalon GWAS in the ABCD cohort. Genome-wide significance is shown for the common threshold of *p*-value < $5x10^{-8}$ (red dashed line), and also for the multiple comparisons-corrected threshold of *p*-value < $6.25x10^{-9}$ (green dashed line). Two-sided P-values shown were derived from the non-infinitesimal mixed model association test p-value as implemented in BOLT-LMM.

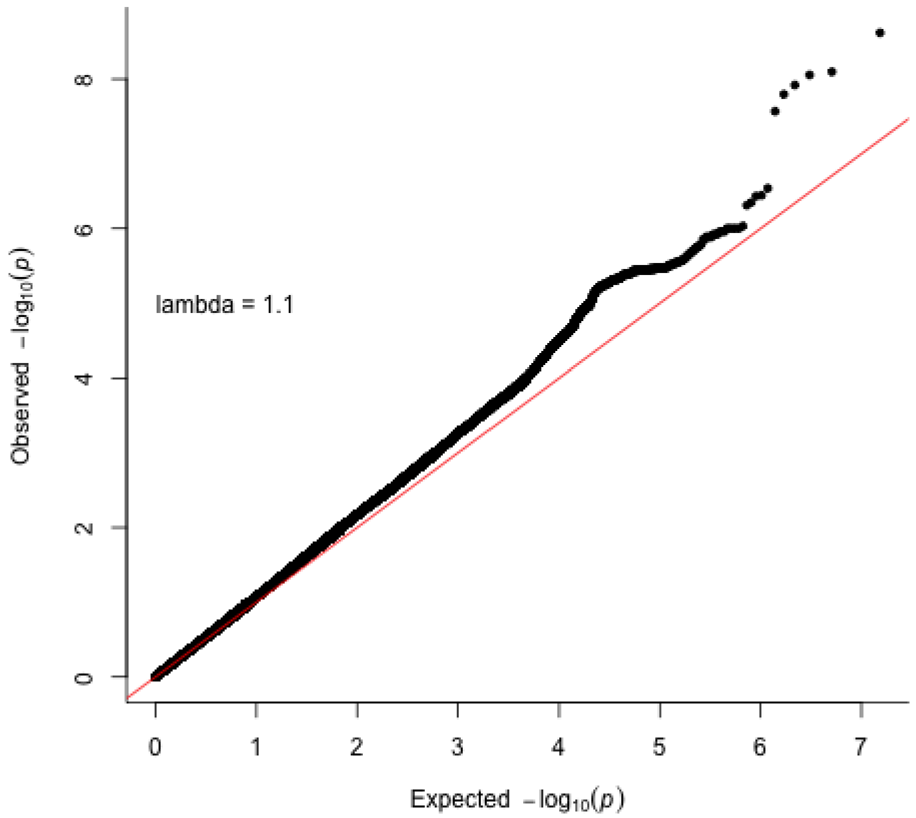

**Supplementary Figure 60. Ventral diencephalon QQ plot in the ABCD cohort.** Two-sided P-values shown were derived from the non-infinitesimal mixed model association test p-value as implemented in BOLT-LMM.

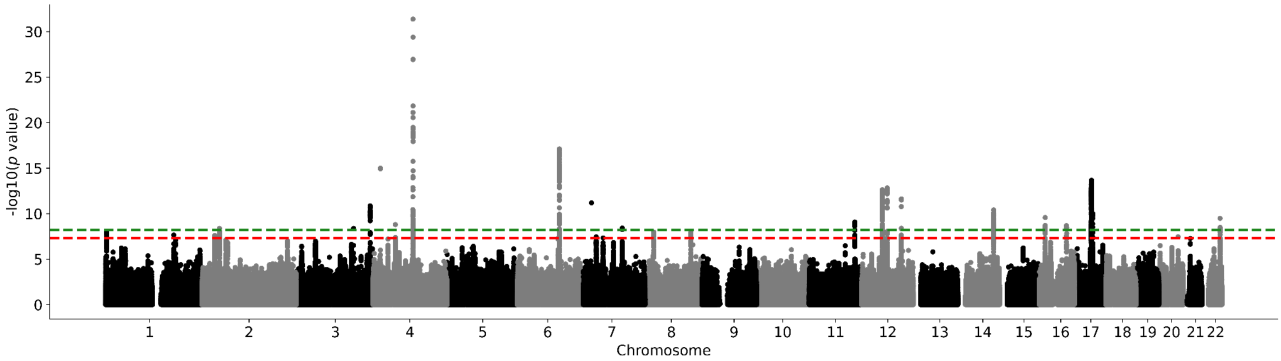

**Supplementary Figure 61. UK Biobank nucleus accumbens Manhattan plot without adjusting for ICV.** Results for nucleus accumbens GWAS in the UK Biobank. Genome-wide significance is shown for the common threshold of *p*-value < $5x10^{-8}$ (red dashed line), and also for the multiple comparisons-corrected threshold of *p*-value < $6.25x10^{-9}$ (green dashed line). Two-sided P-values shown were derived from the non-infinitesimal mixed model association test p-value as implemented in BOLT-LMM.

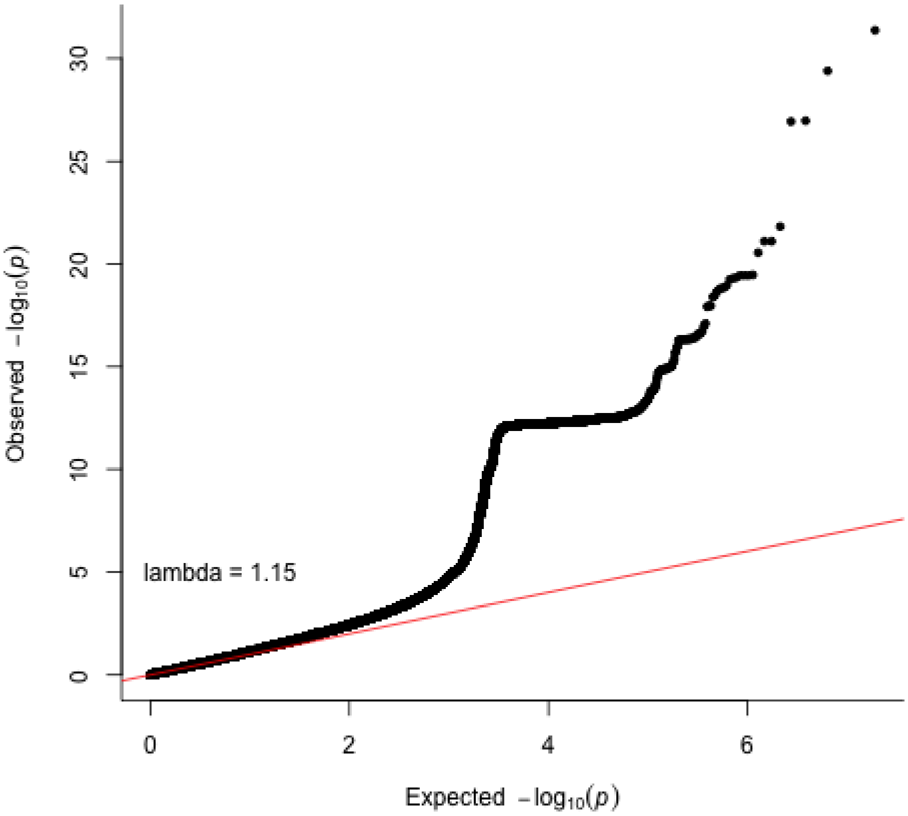

**Supplementary Figure 62. Nucleus accumbens QQ plot in the UK Biobank without adjusting for ICV.** Two-sided P-values shown were derived from the non-infinitesimal mixed model association test p-value as implemented in BOLT-LMM.

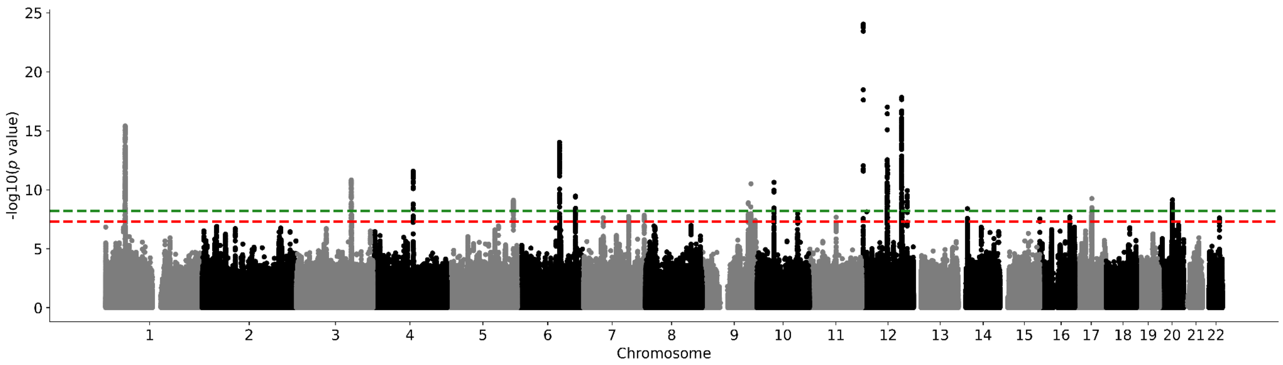

**Supplementary Figure 63. UK Biobank amygdala Manhattan plot without adjusting for ICV.** Results for amygdala GWAS in the UK Biobank. Genome-wide significance is shown for the common threshold of *p*-value < $5x10^{-8}$ (red dashed line), and also for the multiple comparisons-corrected threshold of *p*-value < $6.25x10^{-9}$ (green dashed line). Two-sided P-values shown were derived from the non-infinitesimal mixed model association test p-value as implemented in BOLT-LMM.

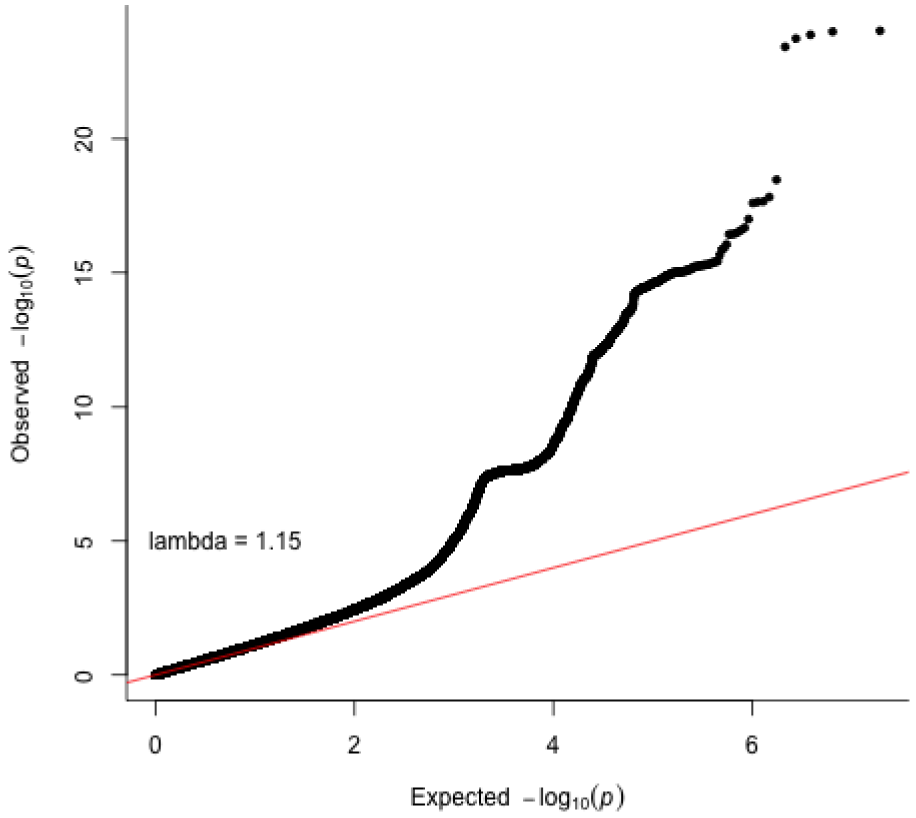

**Supplementary Figure 64. Amygdala QQ plot in the UK Biobank without adjusting for ICV.** Two-sided P-values shown were derived from the non-infinitesimal mixed model association test p-value as implemented in BOLT-LMM.

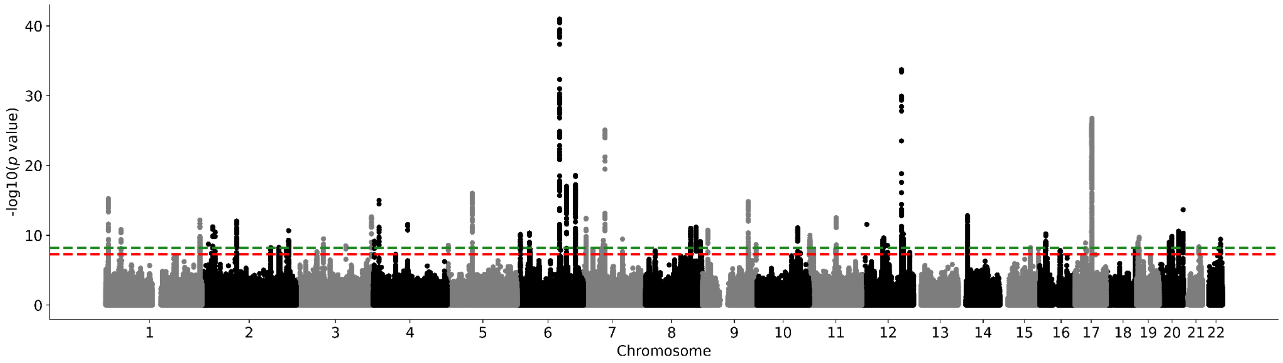

**Supplementary Figure 65. UK Biobank brainstem Manhattan plot without adjusting for ICV.** Results for brainstem GWAS in the UK Biobank. Genome-wide significance is shown for the common threshold of *p*-value < $5x10^{-8}$ (red dashed line), and also for the multiple comparisons-corrected threshold of *p*-value < $6.25x10^{-9}$ (green dashed line). Two-sided P-values shown were derived from the non-infinitesimal mixed model association test p-value as implemented in BOLT-LMM.

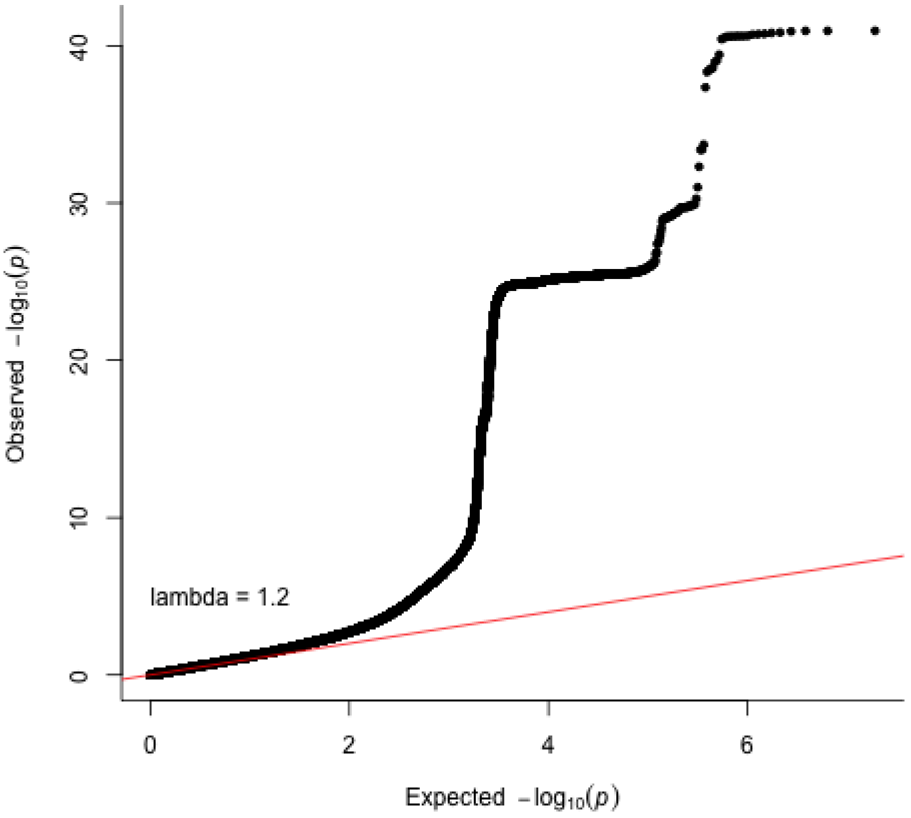

**Supplementary Figure 66. Brainstem QQ plot in the UK Biobank without adjusting for ICV.** Two-sided P-values shown were derived from the non-infinitesimal mixed model association test p-value as implemented in BOLT-LMM.

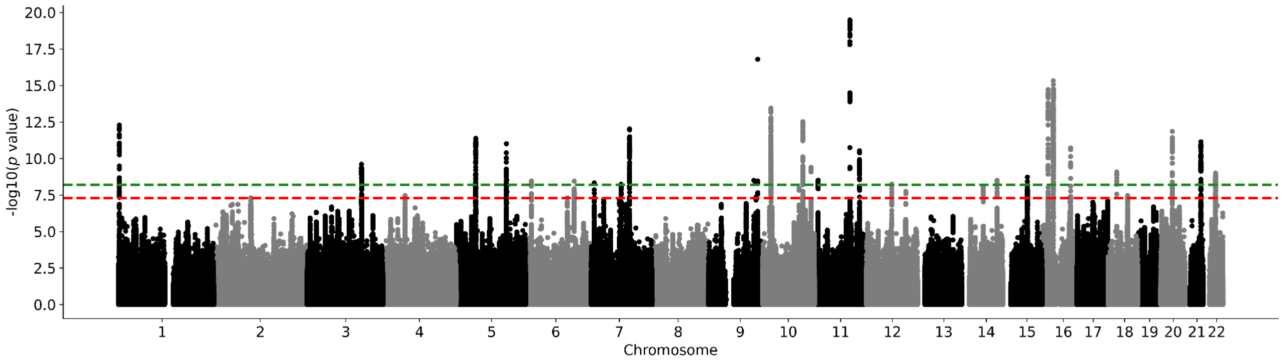

**Supplementary Figure 67. UK Biobank caudate nucleus Manhattan plot without adjusting for ICV.** Results for caudate nucleus GWAS in the UK Biobank. Genome-wide significance is shown for the common threshold of *p*-value < $5x10^{-8}$ (red dashed line), and also for the multiple comparisons-corrected threshold of *p*-value < $6.25x10^{-9}$ (green dashed line). Two-sided P-values shown were derived from the non-infinitesimal mixed model association test p-value as implemented in BOLT-LMM.

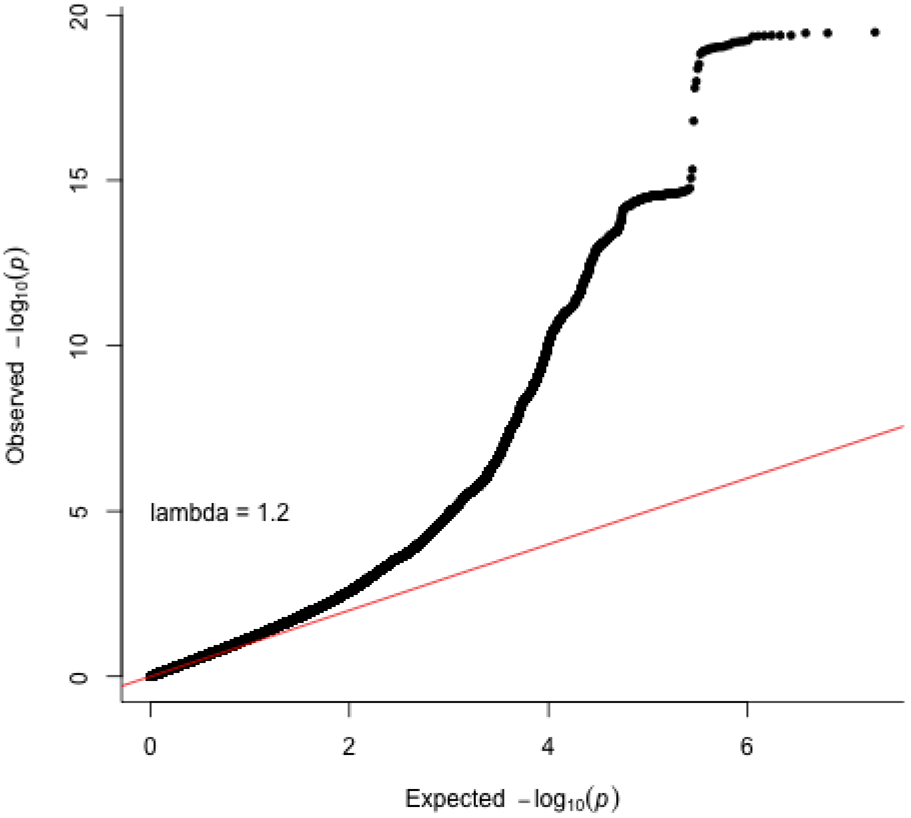

**Supplementary Figure 68. Caudate nucleus QQ plot in the UK Biobank without adjusting for ICV.** Two-sided P-values shown were derived from the non-infinitesimal mixed model association test p-value as implemented in BOLT-LMM.

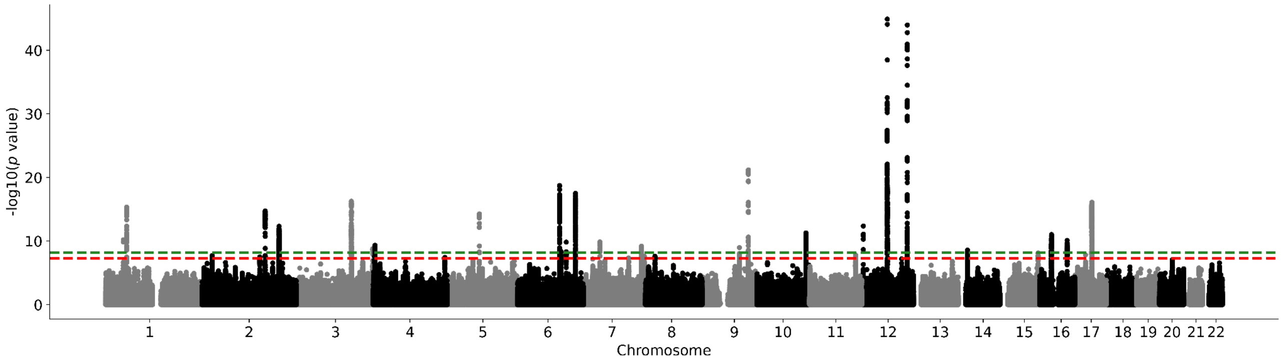

**Supplementary Figure 69. UK Biobank hippocampus Manhattan plot without adjusting for ICV.** Results for hippocampus GWAS in the UK Biobank. Genome-wide significance is shown for the common threshold of $p$-value < $5x10^{-8}$ (red dashed line), and also for the multiple comparisons-corrected threshold of $p$-value < $6.25x10^{-9}$ (green dashed line). Two-sided P-values shown were derived from the non-infinitesimal mixed model association test p-value as implemented in BOLT-LMM.

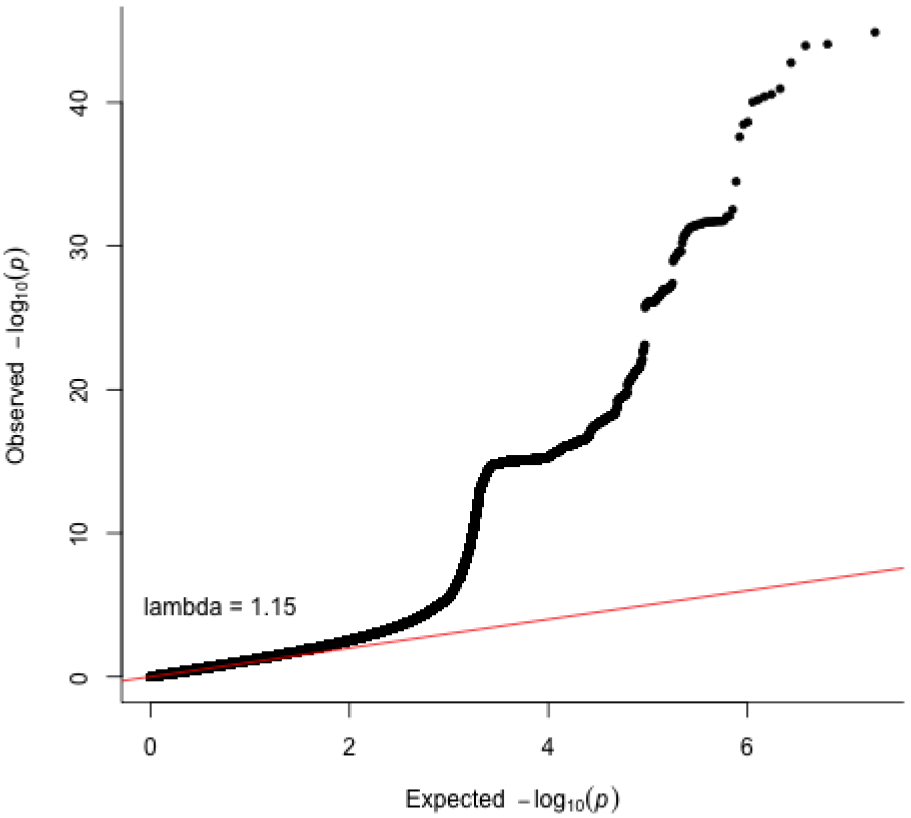

**Supplementary Figure 70. Hippocampus QQ plot in the UK Biobank without adjusting for ICV.** Two-sided P-values shown were derived from the non-infinitesimal mixed model association test p-value as implemented in BOLT-LMM.

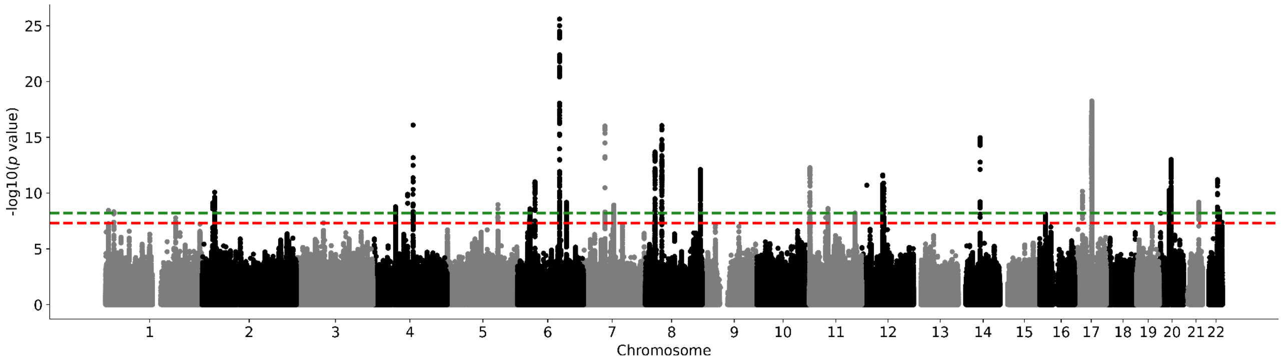

**Supplementary Figure 71. UK Biobank globus pallidus Manhattan plot without adjusting for ICV.** Results for globus pallidus GWAS in the UK Biobank. Genome-wide significance is shown for the common threshold of *p*-value < $5 \times 10^{-8}$ (red dashed line), and also for the multiple comparisons-corrected threshold of *p*-value < $6.25 \times 10^{-9}$ (green dashed line). Two-sided P-values shown were derived from the non-infinitesimal mixed model association test p-value as implemented in BOLT-LMM.

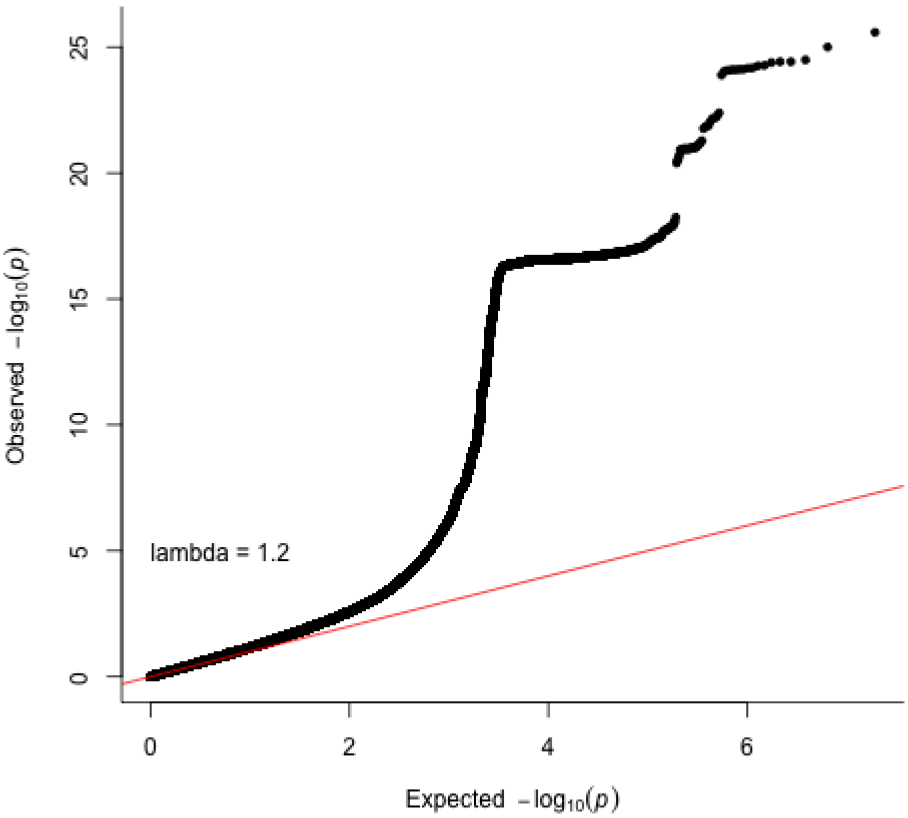

**Supplementary Figure 72. Globus pallidum QQ plot in the UK Biobank without adjusting for ICV.** Two-sided P-values shown were derived from the non-infinitesimal mixed model association test p-value as implemented in BOLT-LMM.

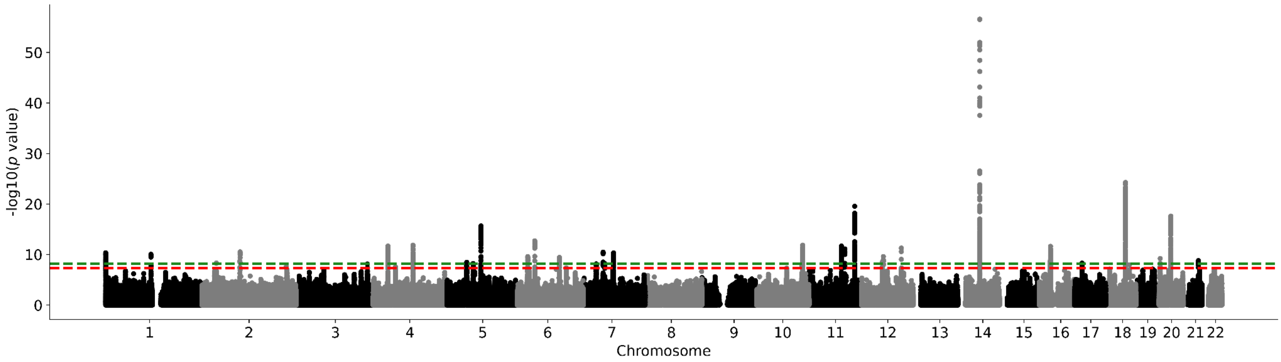

**Supplementary Figure 73. UK Biobank putamen Manhattan plot without adjusting for ICV.** Results for putamen GWAS in the UK Biobank. Genome-wide significance is shown for the common threshold of *p*-value < $5x10^{-8}$ (red dashed line), and also for the multiple comparisons-corrected threshold of *p*-value < $6.25x10^{-9}$ (green dashed line). Two-sided P-values shown were derived from the non-infinitesimal mixed model association test p-value as implemented in BOLT-LMM.

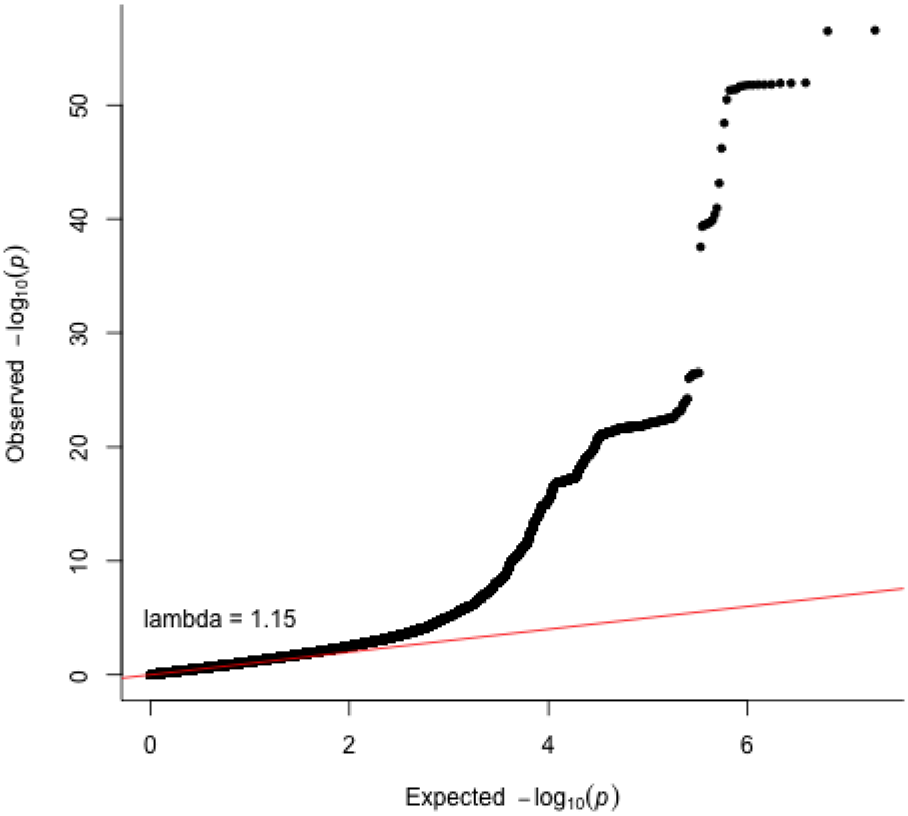

**Supplementary Figure 74. Putamen QQ plot in the UK Biobank without adjusting for ICV.** Two-sided P-values shown were derived from the non-infinitesimal mixed model association test p-value as implemented in BOLT-LMM.

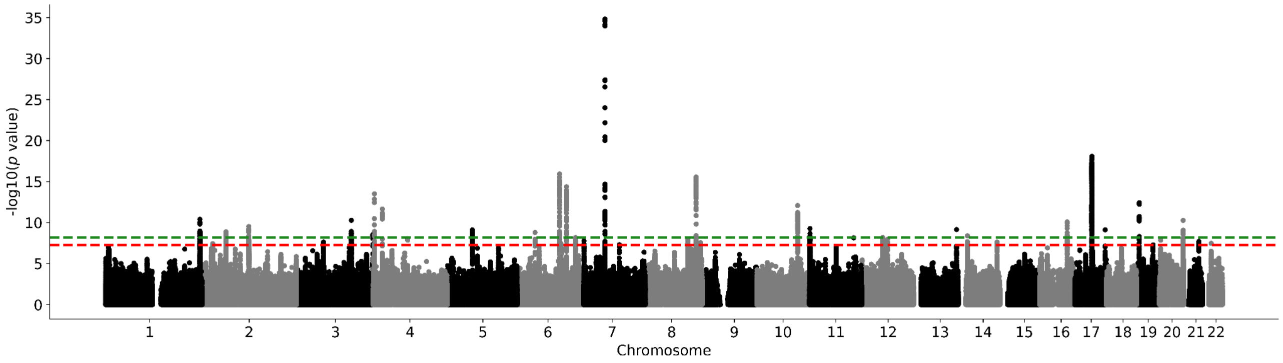

**Supplementary Figure 75. UK Biobank thalamus Manhattan plot without adjusting for ICV.** Results for thalamus GWAS in the UK Biobank. Genome-wide significance is shown for the common threshold of *p*-value < $5x10^{-8}$ (red dashed line), and also for the multiple comparisons-corrected threshold of *p*-value < $6.25x10^{-9}$ (green dashed line). Two-sided P-values shown were derived from the non-infinitesimal mixed model association test p-value as implemented in BOLT-LMM.

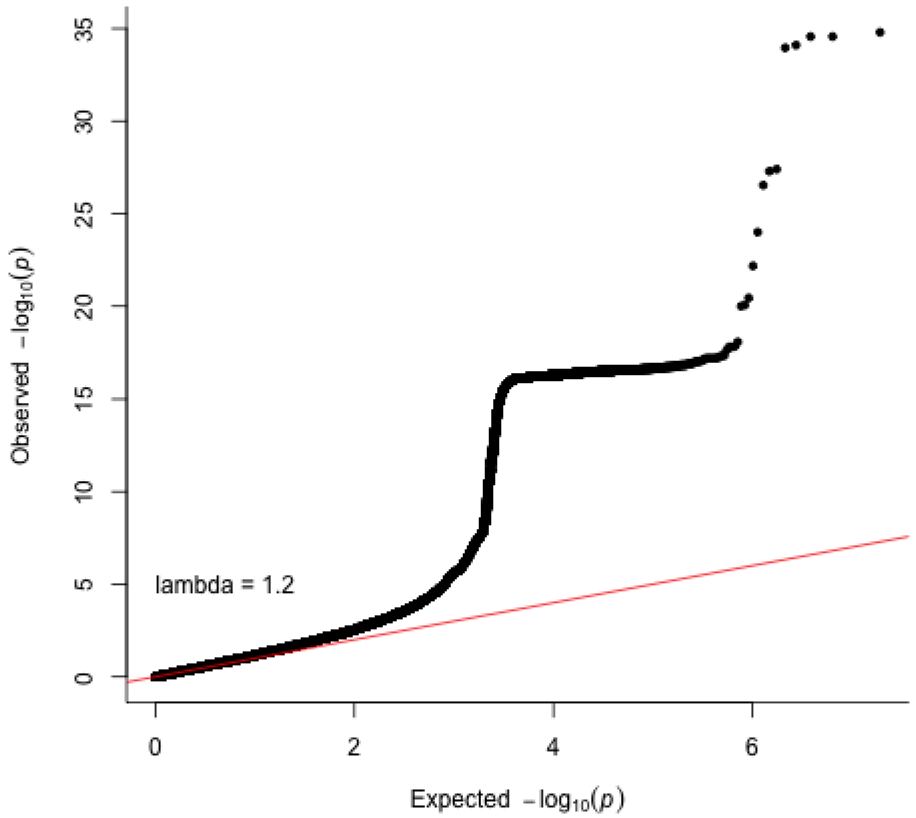

**Supplementary Figure 76. Thalamus QQ plot in the UK Biobank without adjusting for ICV.** Two-sided P-values shown were derived from the non-infinitesimal mixed model association test p-value as implemented in BOLT-LMM.

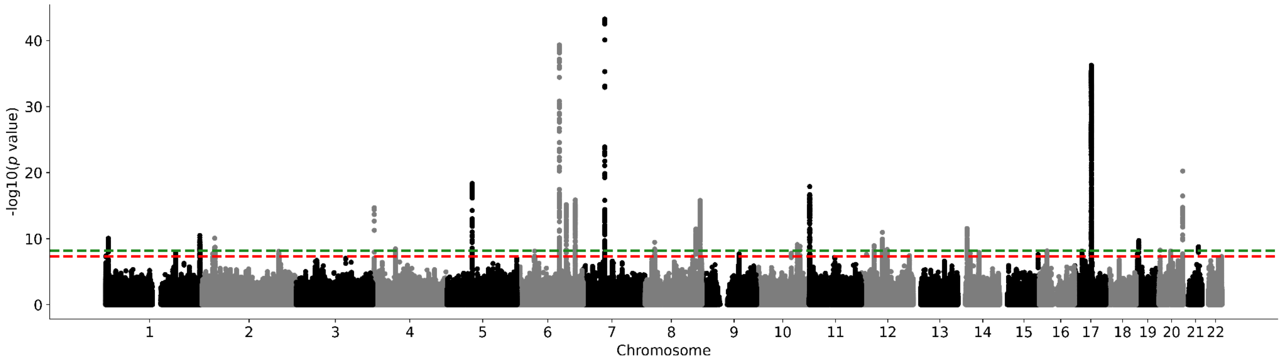

**Supplementary Figure 77. UK Biobank ventral diencephalon Manhattan plot without adjusting for ICV.** Results for ventral diencephalon GWAS in the UK Biobank. Genome-wide significance is shown for the common threshold of $p$-value < $5x10^{-8}$ (red dashed line), and also for the multiple comparisons-corrected threshold of $p$-value < $6.25x10^{-9}$ (green dashed line). Two-sided P-values shown were derived from the non-infinitesimal mixed model association test p-value as implemented in BOLT-LMM.

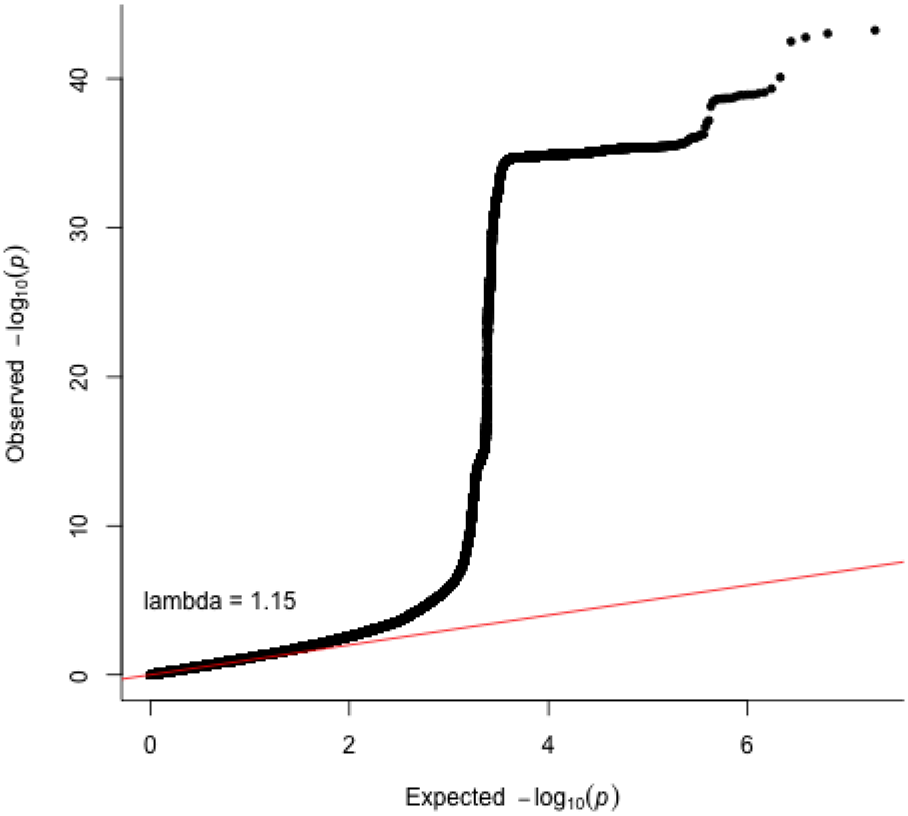

**Supplementary Figure 78. Ventral diencephalon QQ plot in the UK Biobank without adjusting for ICV.** Two-sided P-values shown were derived from the non-infinitesimal mixed model association test p-value as implemented in BOLT-LMM.

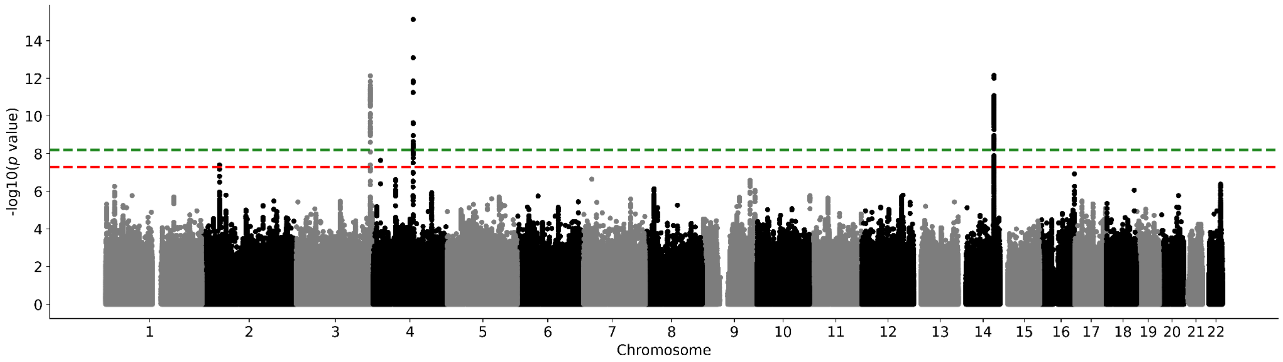

**Supplementary Figure 79. UK Biobank subsample 1 nucleus accumbens Manhattan plot.** Results for nucleus accumbens GWAS in the UK Biobank. Genome-wide significance is shown for the common threshold of *p*-value < $5x10^{-8}$ (red dashed line), and also for the multiple comparisons-corrected threshold of *p*-value < $6.25x10^{-9}$ (green dashed line). Two-sided P-values shown were derived from the non-infinitesimal mixed model association test p-value as implemented in BOLT-LMM.

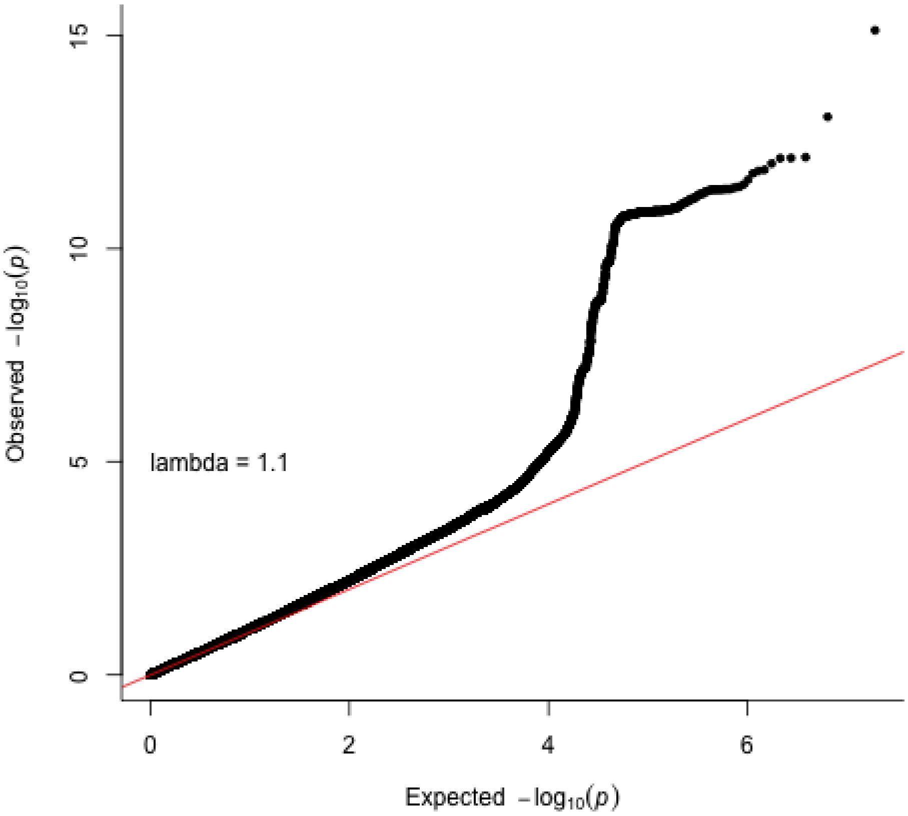

**Supplementary Figure 80. Nucleus accumbens QQ plot in the UK Biobank subsample 1.** Two-sided P-values shown were derived from the non-infinitesimal mixed model association test p-value as implemented in BOLT-LMM.

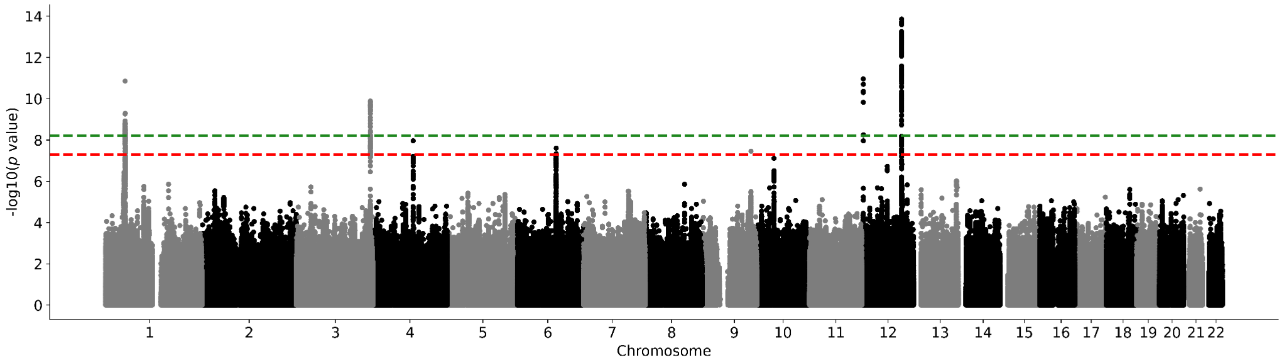

**Supplementary Figure 81. UK Biobank subsample 1 amygdala Manhattan plot.** Results for amygdala GWAS in the UK Biobank. Genome-wide significance is shown for the common threshold of *p*-value < $5x10^{-8}$ (red dashed line), and also for the multiple comparisons-corrected threshold of *p*-value < $6.25x10^{-9}$ (green dashed line). Two-sided P-values shown were derived from the non-infinitesimal mixed model association test p-value as implemented in BOLT-LMM.

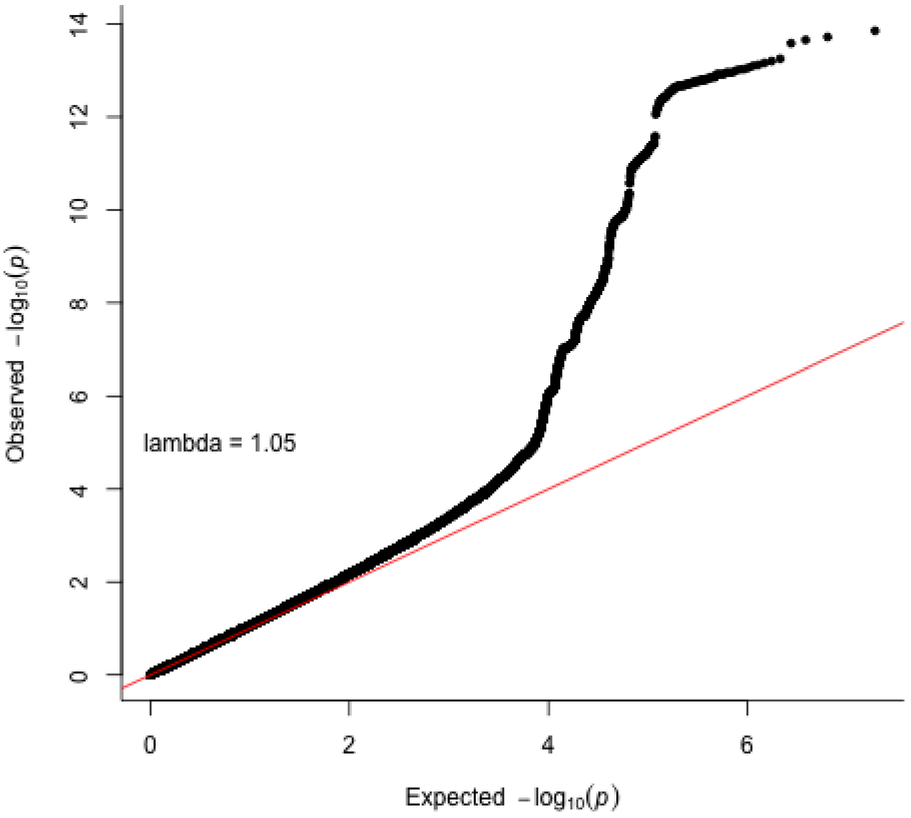

**Supplementary Figure 82. Amygdala QQ plot in the UK Biobank subsample 1.** Two-sided P-values shown were derived from the non-infinitesimal mixed model association test p-value as implemented in BOLT-LMM.

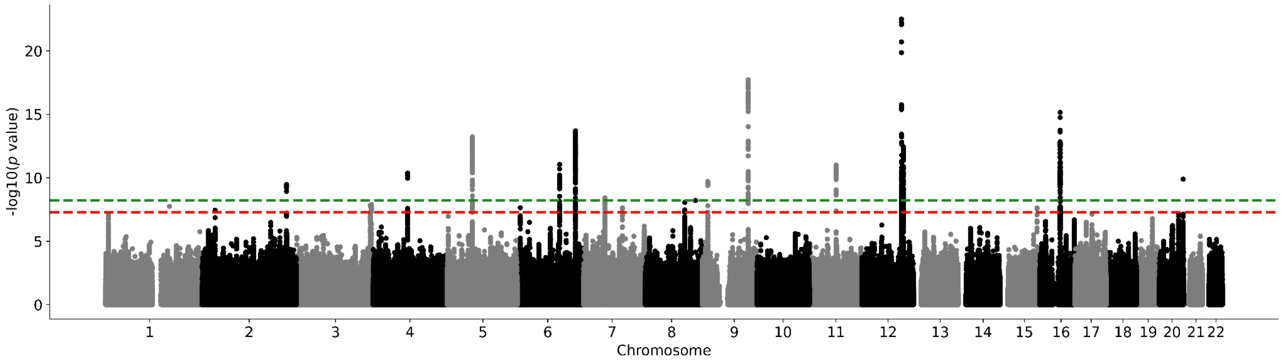

**Supplementary Figure 83. UK Biobank subsample 1 brainstem Manhattan plot.** Results for brainstem GWAS in the UK Biobank. Genome-wide significance is shown for the common threshold of *p*-value < $5x10^{-8}$ (red dashed line), and also for the multiple comparisons-corrected threshold of *p*-value < $6.25x10^{-9}$ (green dashed line). Two-sided P-values shown were derived from the non-infinitesimal mixed model association test p-value as implemented in BOLT-LMM.

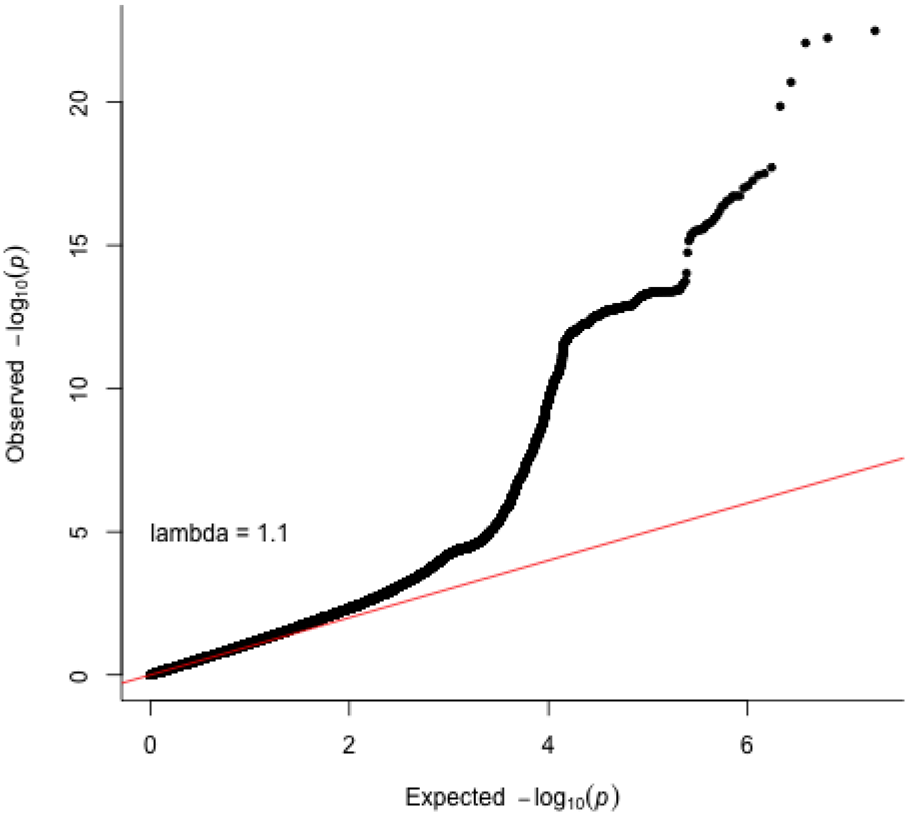

**Supplementary Figure 84. Brainstem QQ plot in the UK Biobank subsample 1.** Two-sided P-values shown were derived from the non-infinitesimal mixed model association test p-value as implemented in BOLT-LMM.

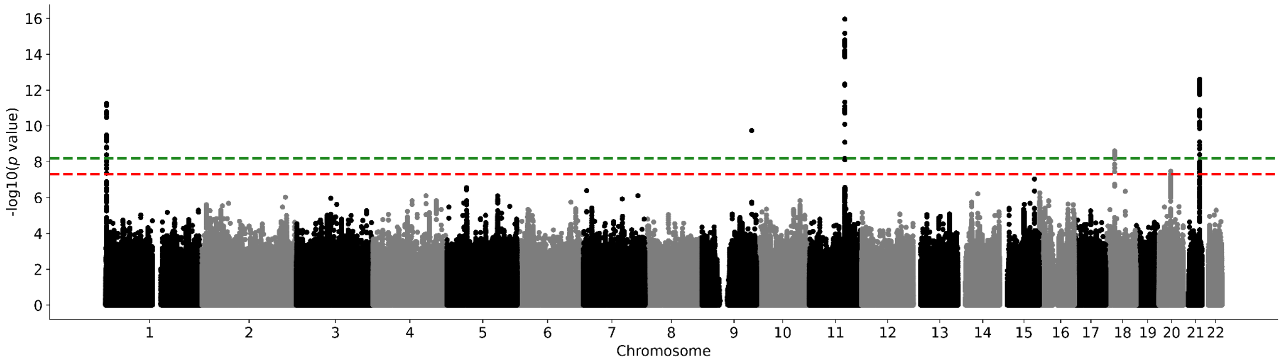

**Supplementary Figure 85. UK Biobank subsample 1 caudate nucleus Manhattan plot.** Results for caudate nucleus GWAS in the UK Biobank. Genome-wide significance is shown for the common threshold of *p*-value < $5x10^{-8}$ (red dashed line), and also for the multiple comparisons-corrected threshold of *p*-value < $6.25x10^{-9}$ (green dashed line). Two-sided P-values shown were derived from the non-infinitesimal mixed model association test p-value as implemented in BOLT-LMM.

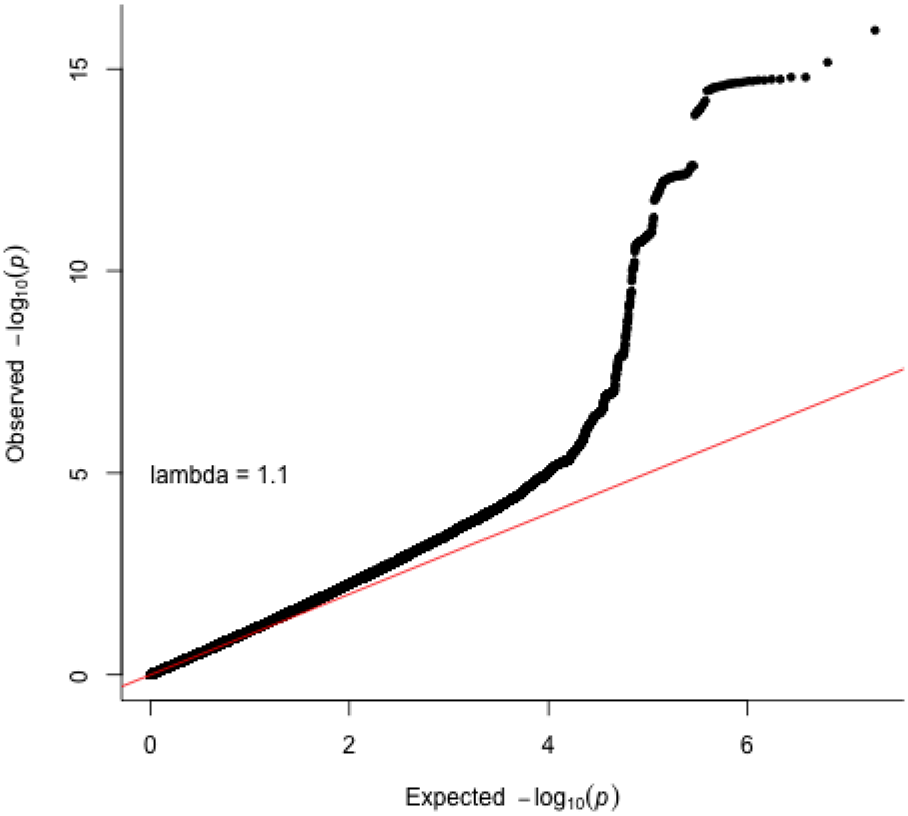

**Supplementary Figure 86. Caudate nucleus QQ plot in the UK Biobank subsample 1.** Two-sided P-values shown were derived from the non-infinitesimal mixed model association test p-value as implemented in BOLT-LMM.

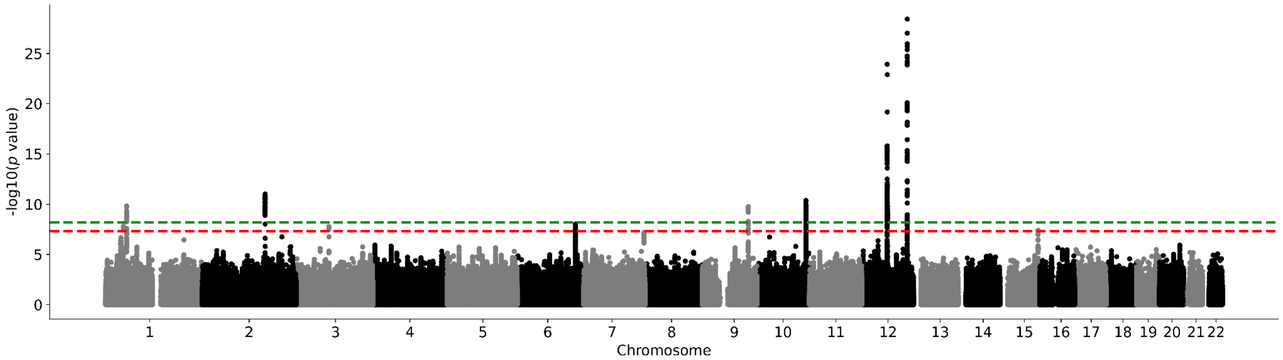

**Supplementary Figure 87. UK Biobank subsample 1 hippocampus Manhattan plot.** Results for hippocampus GWAS in the UK Biobank. Genome-wide significance is shown for the common threshold of *p*-value < $5x10^{-8}$ (red dashed line), and also for the multiple comparisons-corrected threshold of *p*-value < $6.25x10^{-9}$ (green dashed line). Two-sided P-values shown were derived from the non-infinitesimal mixed model association test p-value as implemented in BOLT-LMM.

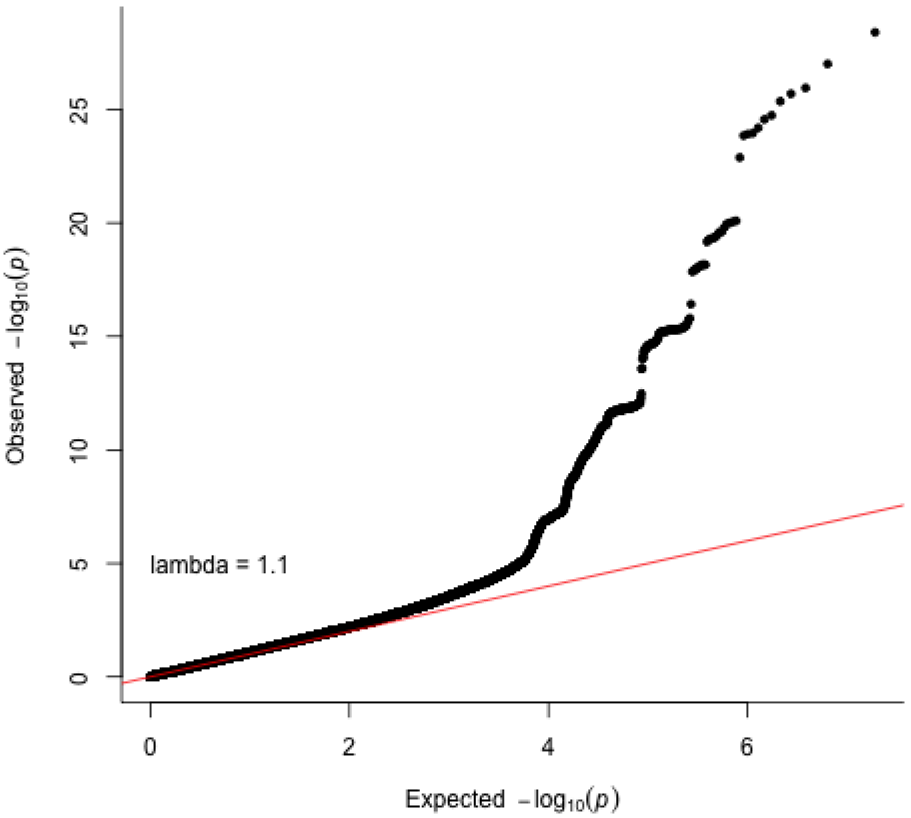

**Supplementary Figure 88. Hippocampus QQ plot in the UK Biobank subsample 1.** Two-sided P-values shown were derived from the non-infinitesimal mixed model association test p-value as implemented in BOLT-LMM.

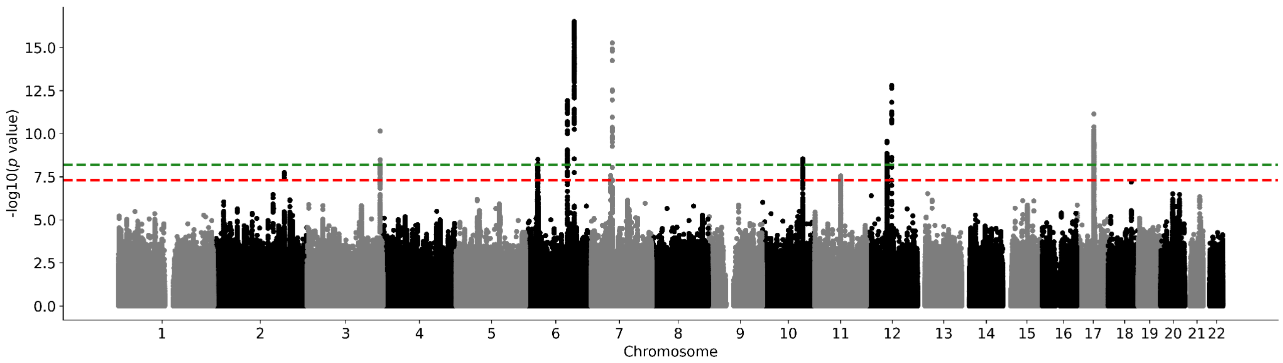

**Supplementary Figure 89. UK Biobank subsample 1 intracranial volume Manhattan plot.** Results for intracranial volume GWAS in the UK Biobank. Genome-wide significance is shown for the common threshold of $p$-value < $5x10^{-8}$ (red dashed line), and also for the multiple comparisons-corrected threshold of $p$-value < $6.25x10^{-9}$ (green dashed line). Two-sided P-values shown were derived from the non-infinitesimal mixed model association test p-value as implemented in BOLT-LMM.

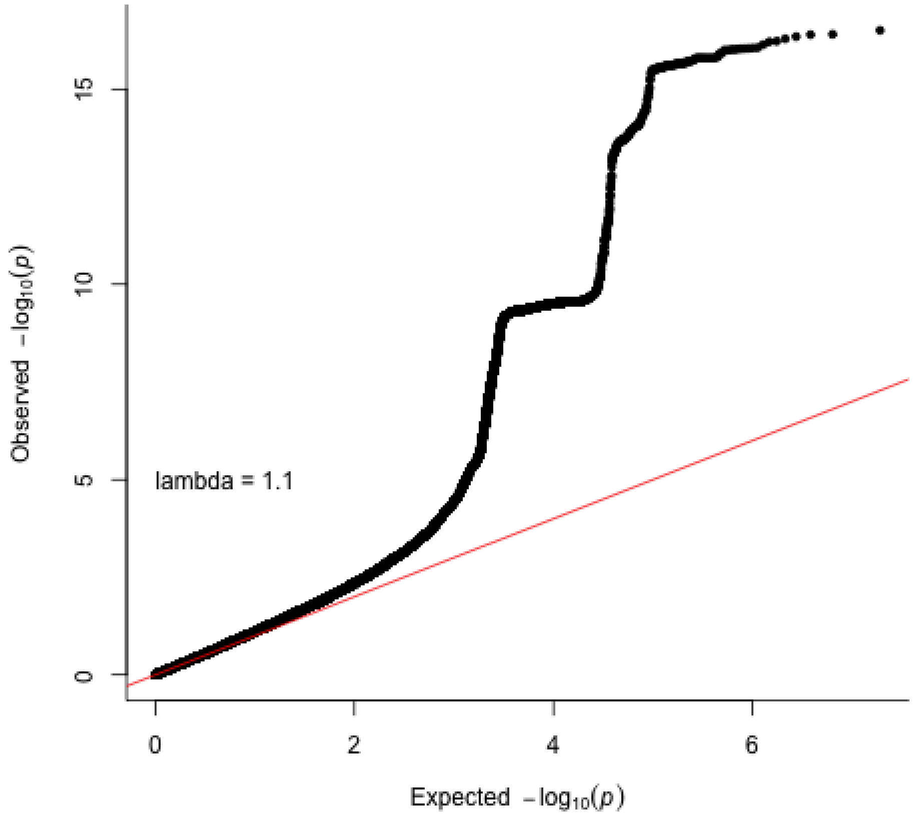

**Supplementary Figure 90. Intracranial volume QQ plot in the UK Biobank subsample 1.** Two-sided P-values shown were derived from the non-infinitesimal mixed model association test p-value as implemented in BOLT-LMM.

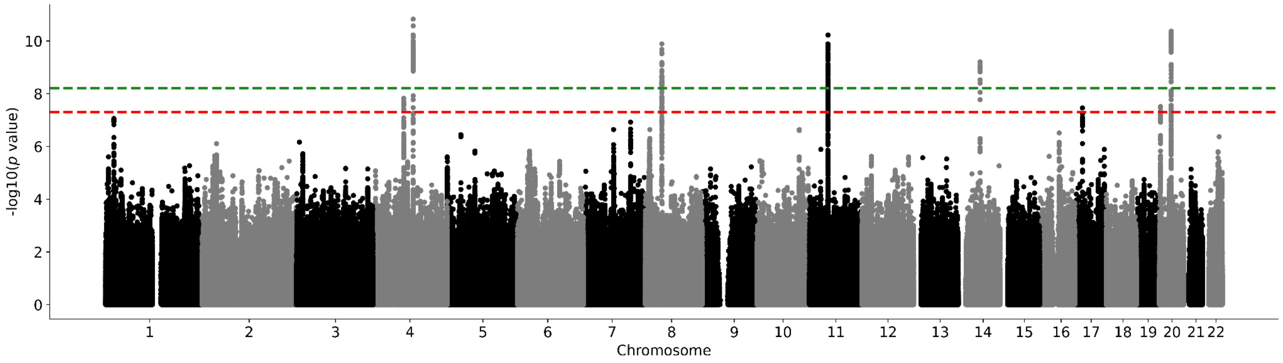

**Supplementary Figure 91. UK Biobank subsample 1 globus pallidus Manhattan plot.** Results for globus pallidus GWAS in the UK Biobank. Genome-wide significance is shown for the common threshold of *p*-value < $5x10^{-8}$ (red dashed line), and also for the multiple comparisons-corrected threshold of *p*-value < $6.25x10^{-9}$ (green dashed line). Two-sided P-values shown were derived from the non-infinitesimal mixed model association test p-value as implemented in BOLT-LMM.

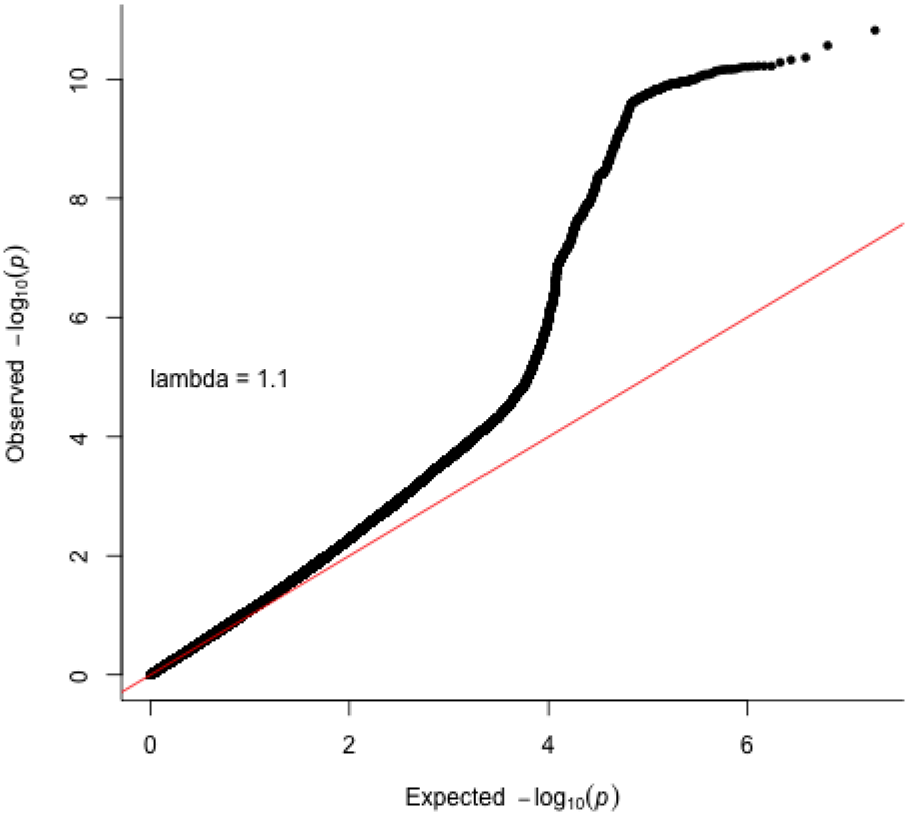

**Supplementary Figure 92. Globus pallidum QQ plot in the UK Biobank subsample 1.** Two-sided P-values shown were derived from the non-infinitesimal mixed model association test p-value as implemented in BOLT-LMM.

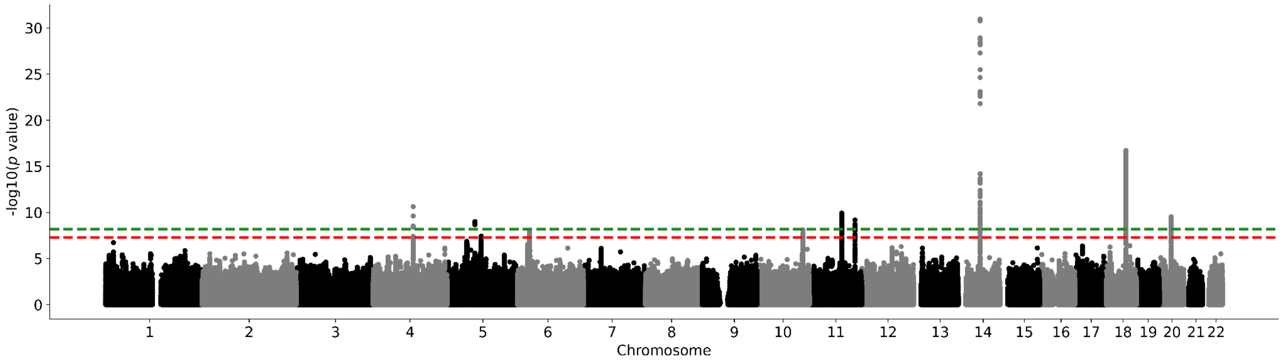

**Supplementary Figure 93. UK Biobank subsample 1 putamen Manhattan plot.** Results for putamen GWAS in the UK Biobank. Genome-wide significance is shown for the common threshold of *p*-value < $5x10^{-8}$ (red dashed line), and also for the multiple comparisons-corrected threshold of *p*-value < $6.25x10^{-9}$ (green dashed line). Two-sided P-values shown were derived from the non-infinitesimal mixed model association test p-value as implemented in BOLT-LMM.

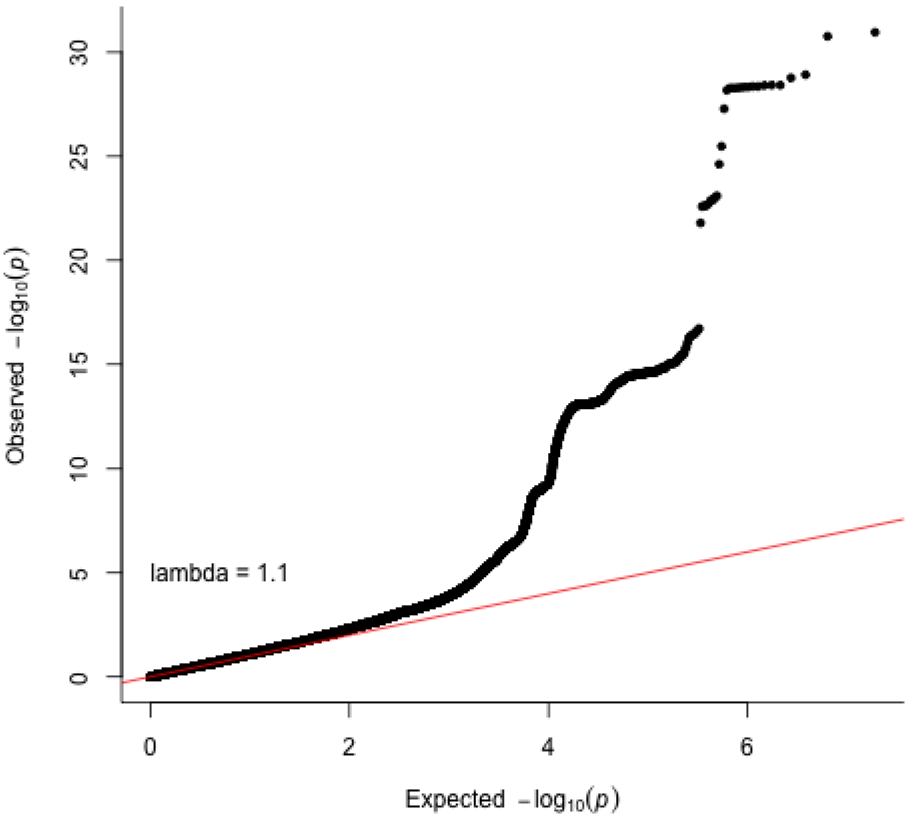

**Supplementary Figure 94. Putamen QQ plot in the UK Biobank subsample 1.** Two-sided P-values shown were derived from the non-infinitesimal mixed model association test p-value as implemented in BOLT-LMM.

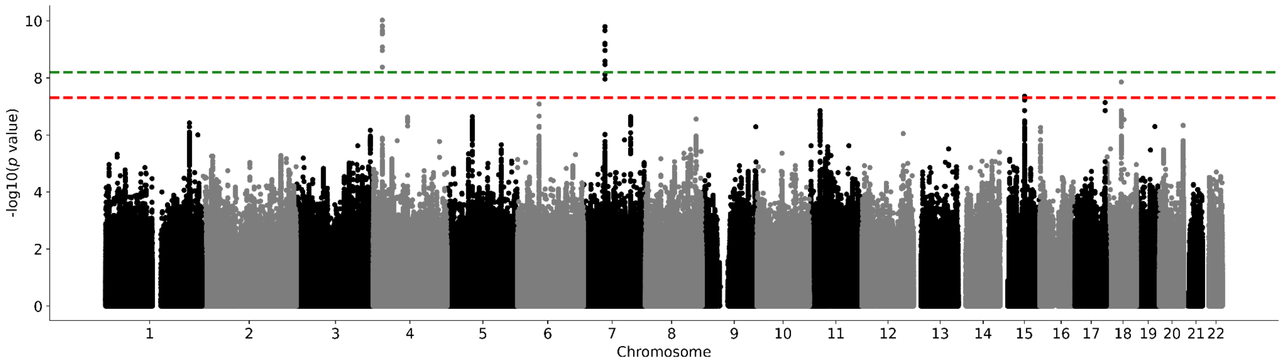

**Supplementary Figure 95. UK Biobank subsample 1 thalamus Manhattan plot.** Results for thalamus GWAS in the UK Biobank. Genome-wide significance is shown for the common threshold of *p*-value < $5x10^{-8}$ (red dashed line), and also for the multiple comparisons-corrected threshold of *p*-value < $6.25x10^{-9}$ (green dashed line). Two-sided P-values shown were derived from the non-infinitesimal mixed model association test p-value as implemented in BOLT-LMM.

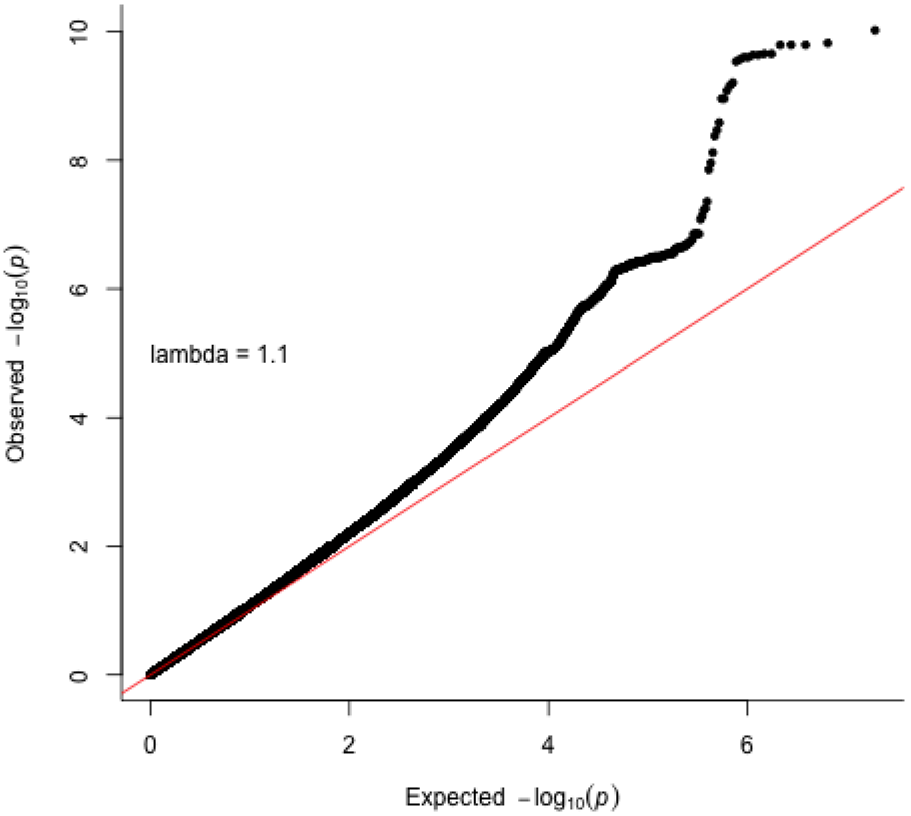

**Supplementary Figure 96. Thalamus QQ plot in the UK Biobank subsample 1.** Two-sided P-values shown were derived from the non-infinitesimal mixed model association test p-value as implemented in BOLT-LMM.

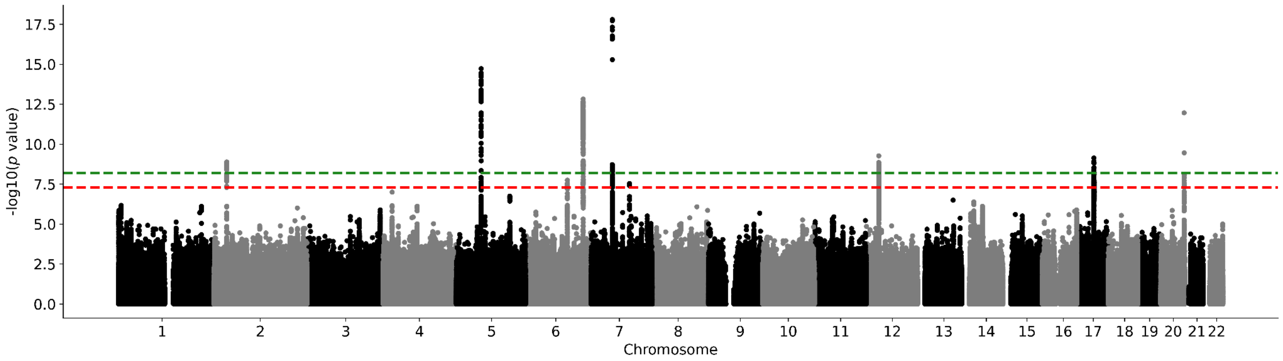

**Supplementary Figure 97. UK Biobank subsample 1 ventral diencephalon Manhattan plot.** Results for ventral diencephalon GWAS in the UK Biobank. Genome-wide significance is shown for the common threshold of *p*-value < $5x10^{-8}$ (red dashed line), and also for the multiple comparisons-corrected threshold of *p*-value < $6.25x10^{-9}$ (green dashed line). Two-sided P-values shown were derived from the non-infinitesimal mixed model association test p-value as implemented in BOLT-LMM.

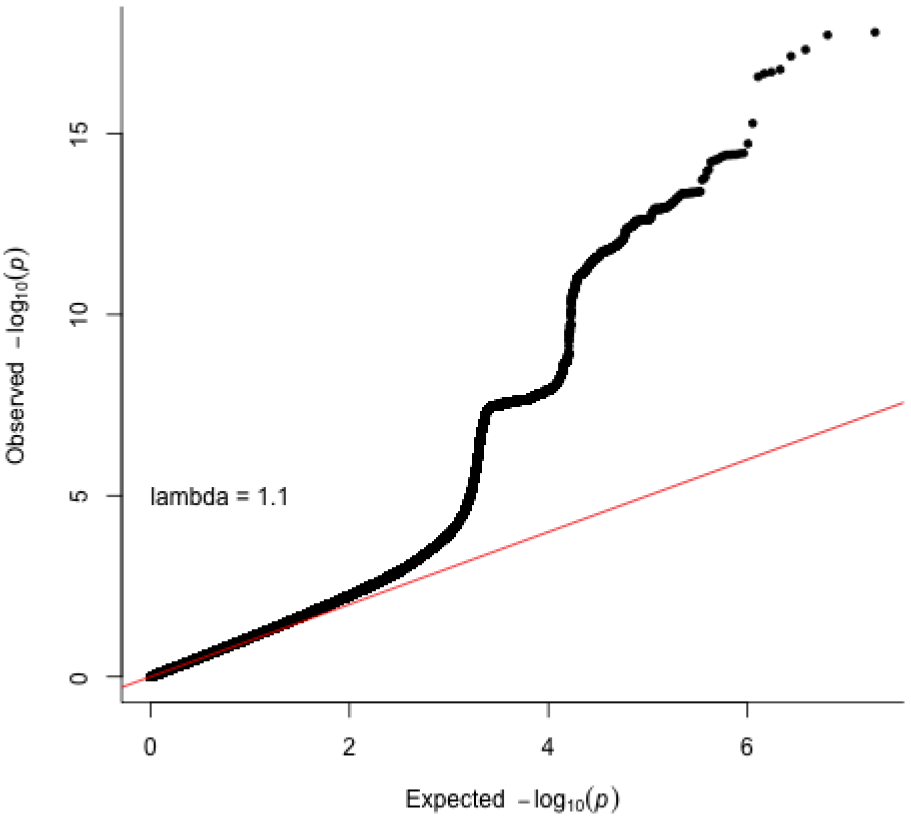

**Supplementary Figure 98. Ventral diencephalon QQ plot in the UK Biobank subsample 1.** Two-sided P-values shown were derived from the non-infinitesimal mixed model association test p-value as implemented in BOLT-LMM.

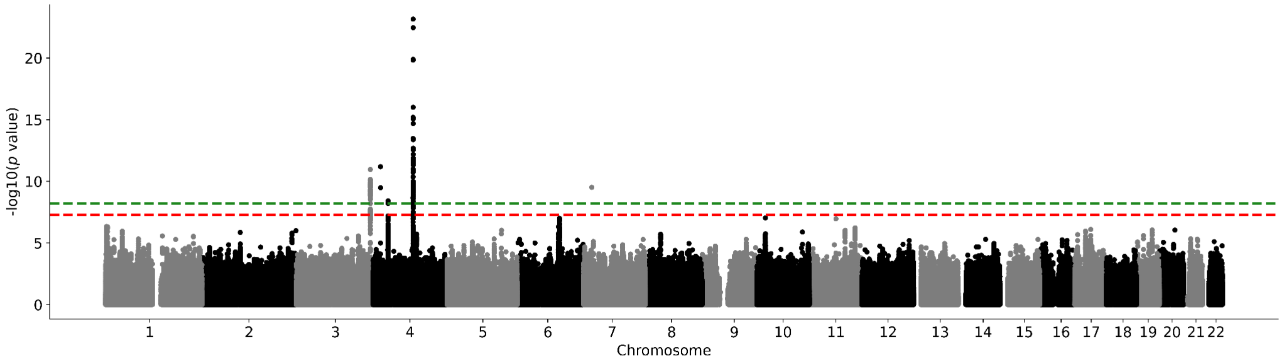

**Supplementary Figure 99. UK Biobank subsample 2 nucleus accumbens Manhattan plot.** Results for nucleus accumbens GWAS in the UK Biobank. Genome-wide significance is shown for the common threshold of *p*-value < $5x10^{-8}$ (red dashed line), and also for the multiple comparisons-corrected threshold of *p*-value < $6.25x10^{-9}$ (green dashed line). Two-sided P-values shown were derived from the non-infinitesimal mixed model association test p-value as implemented in BOLT-LMM.

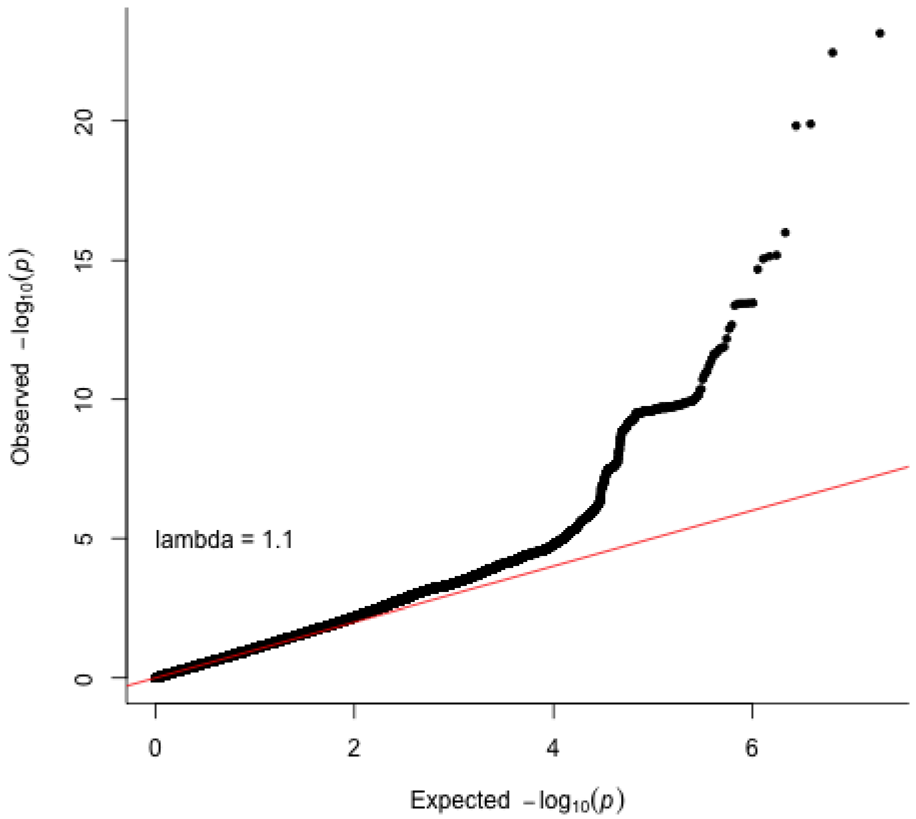

**Supplementary Figure 100. Nucleus accumbens QQ plot in the UK Biobank subsample 2.** Two-sided P-values shown were derived from the non-infinitesimal mixed model association test p-value as implemented in BOLT-LMM.

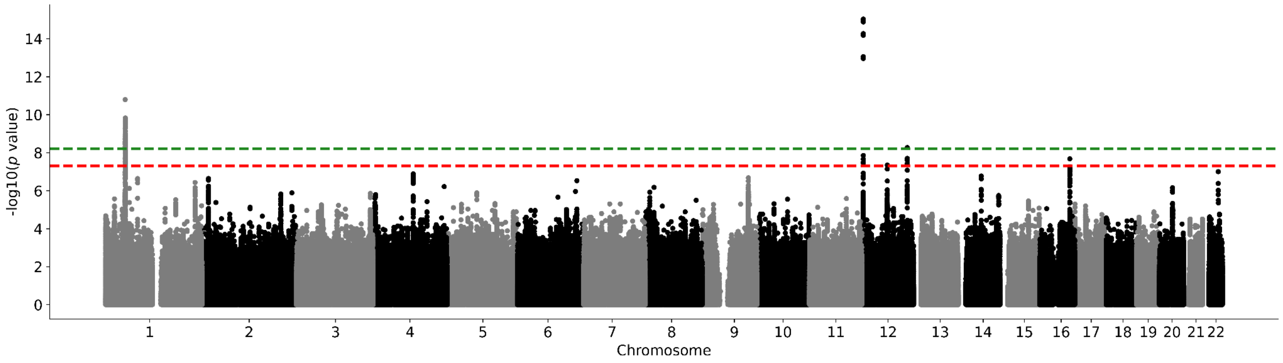

**Supplementary Figure 101. UK Biobank subsample 2 amygdala Manhattan plot.** Results for amygdala GWAS in the UK Biobank. Genome-wide significance is shown for the common threshold of *p*-value < $5 \times 10^{-8}$ (red dashed line), and also for the multiple comparisons-corrected threshold of *p*-value < $6.25 \times 10^{-9}$ (green dashed line). Two-sided P-values shown were derived from the non-infinitesimal mixed model association test p-value as implemented in BOLT-LMM.

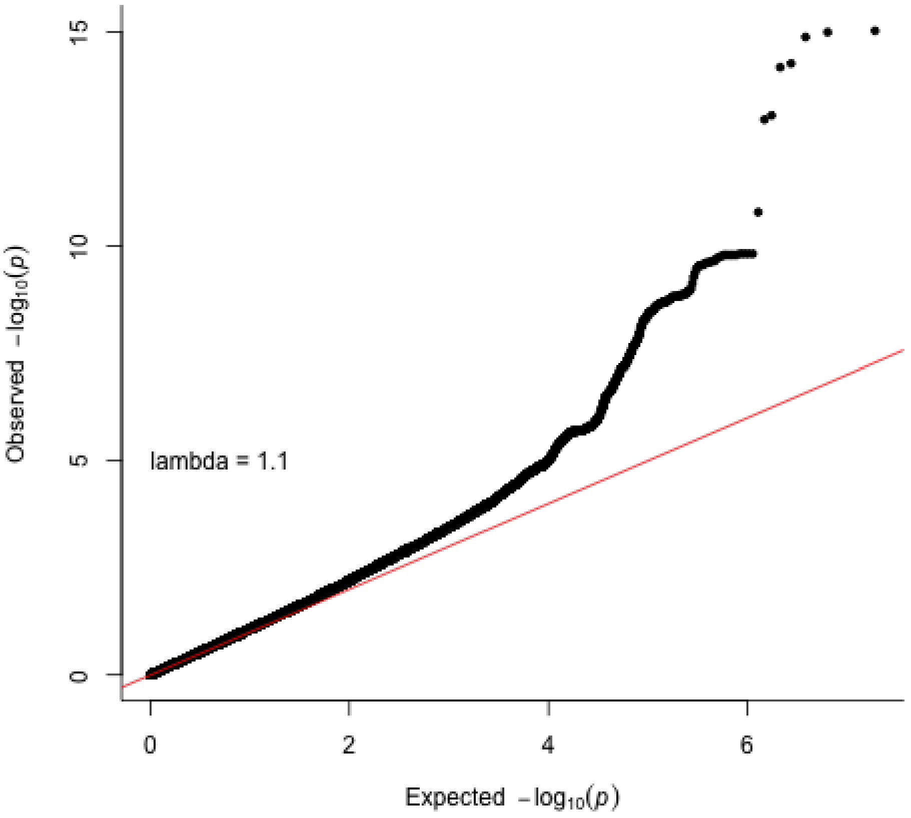

**Supplementary Figure 102. Amygdala QQ plot in the UK Biobank subsample 2.** Two-sided P-values shown were derived from the non-infinitesimal mixed model association test p-value as implemented in BOLT-LMM.

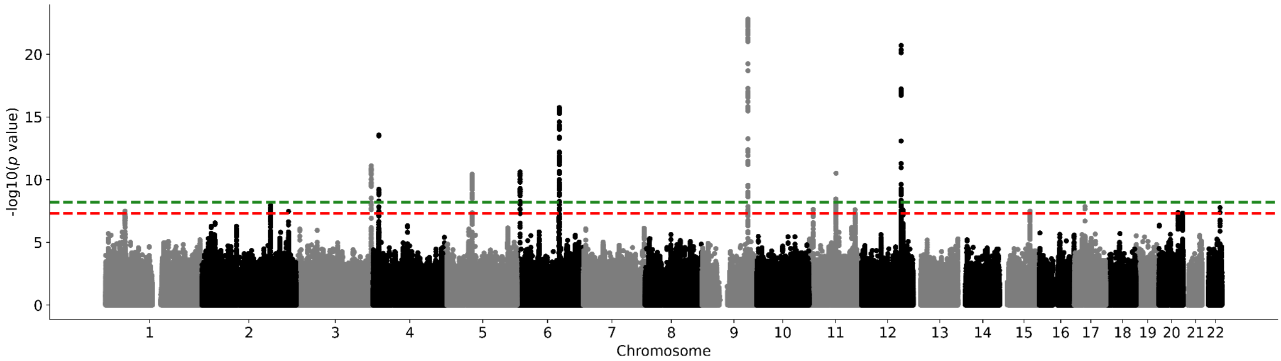

**Supplementary Figure 103. UK Biobank subsample 2 brainstem Manhattan plot.** Results for brainstem GWAS in the UK Biobank. Genome-wide significance is shown for the common threshold of *p*-value < 5x10$^{-8}$ (red dashed line), and also for the multiple comparisons-corrected threshold of *p*-value < 6.25x10$^{-9}$ (green dashed line). Two-sided P-values shown were derived from the non-infinitesimal mixed model association test p-value as implemented in BOLT-LMM.

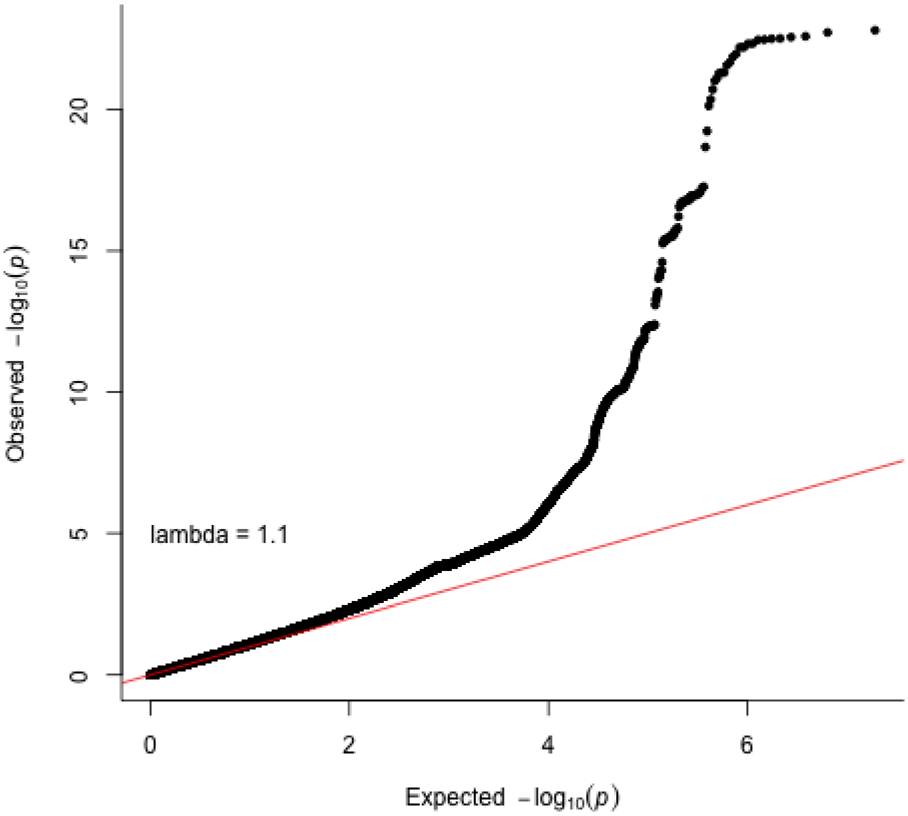

**Supplementary Figure 104. Brainstem QQ plot in the UK Biobank subsample 2.** Two-sided P-values shown were derived from the non-infinitesimal mixed model association test p-value as implemented in BOLT-LMM.

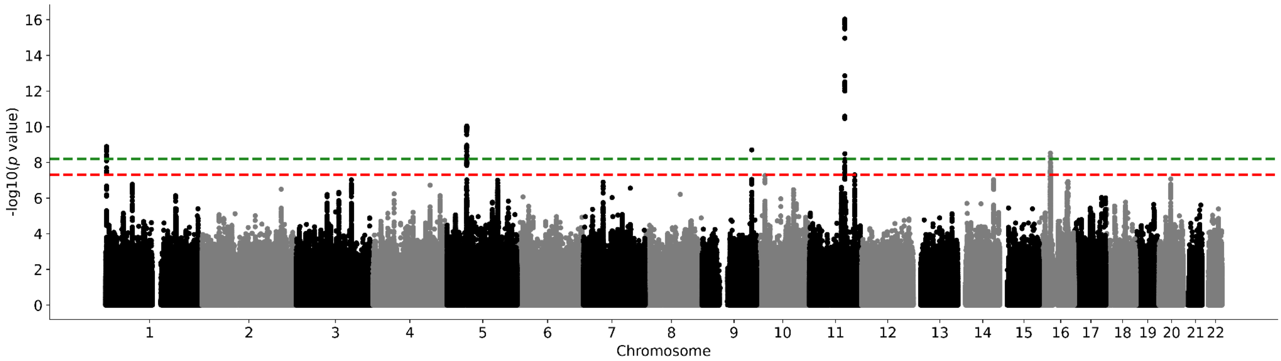

**Supplementary Figure 105. UK Biobank subsample 2 caudate nucleus Manhattan plot.** Results for caudate nucleus GWAS in the UK Biobank. Genome-wide significance is shown for the common threshold of *p*-value < $5x10^{-8}$ (red dashed line), and also for the multiple comparisons-corrected threshold of *p*-value < $6.25x10^{-9}$ (green dashed line). Two-sided P-values shown were derived from the non-infinitesimal mixed model association test p-value as implemented in BOLT-LMM.

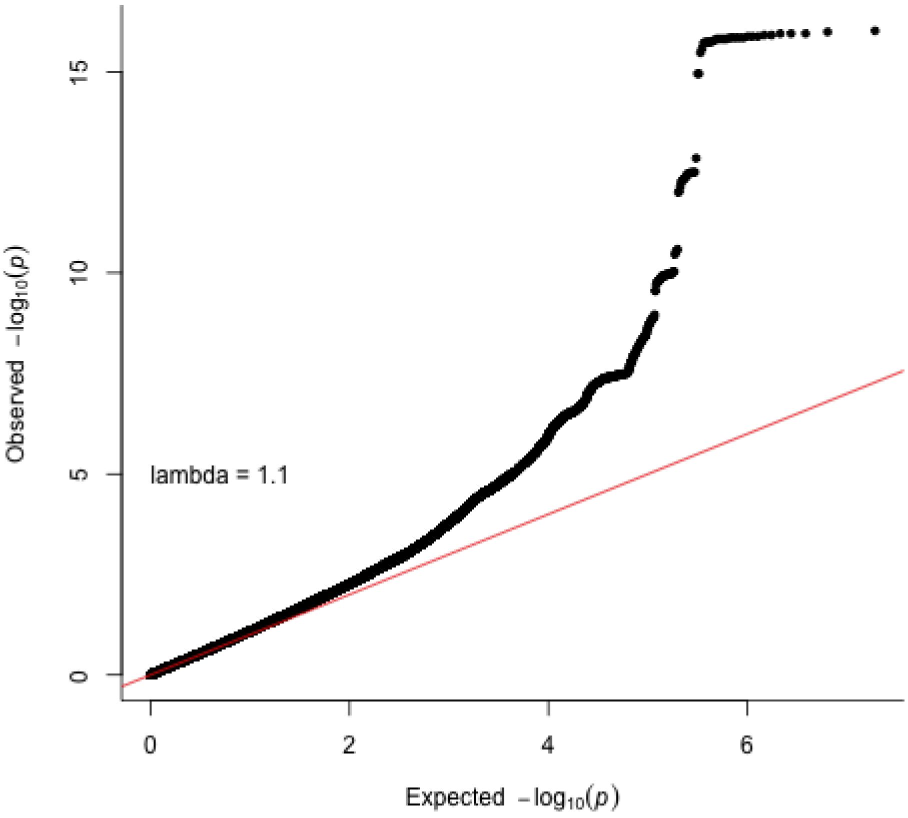

**Supplementary Figure 106. Caudate nucleus QQ plot in the UK Biobank subsample 2.** Two-sided P-values shown were derived from the non-infinitesimal mixed model association test p-value as implemented in BOLT-LMM.

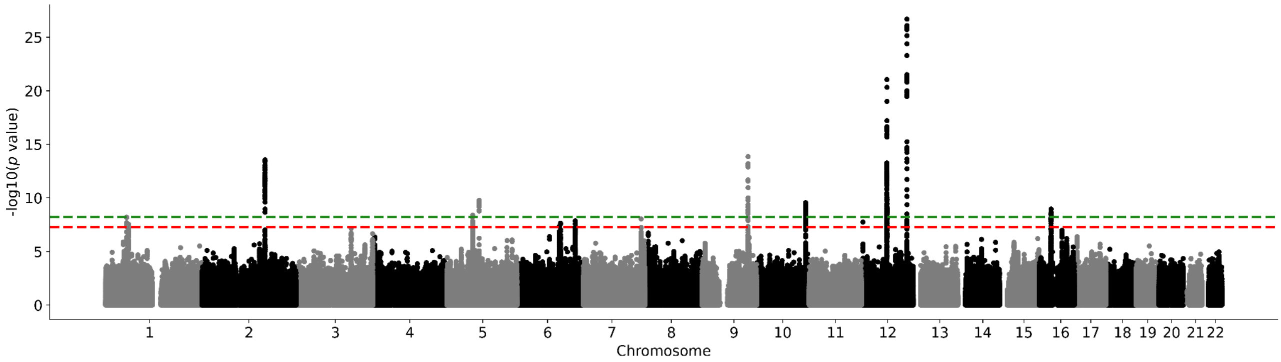

**Supplementary Figure 107. UK Biobank subsample 2 hippocampus Manhattan plot.** Results for hippocampus GWAS in the UK Biobank. Genome-wide significance is shown for the common threshold of *p*-value < 5x10$^{-8}$ (red dashed line), and also for the multiple comparisons-corrected threshold of *p*-value < 6.25x10$^{-9}$ (green dashed line). Two-sided P-values shown were derived from the non-infinitesimal mixed model association test p-value as implemented in BOLT-LMM.

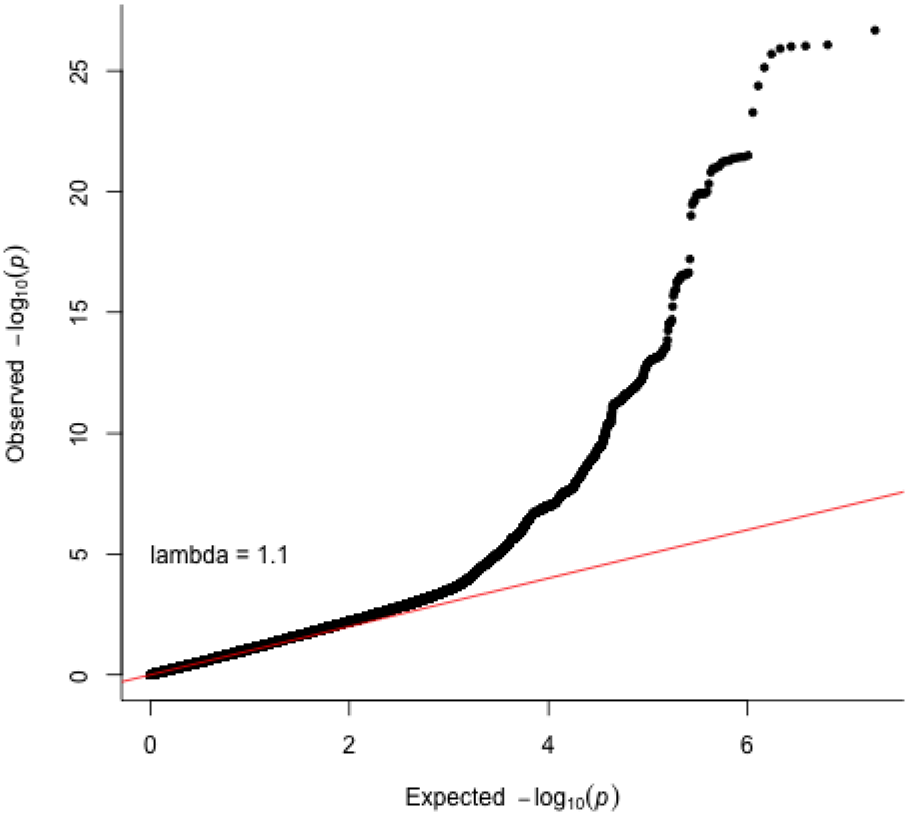

**Supplementary Figure 108. Hippocampus QQ plot in the UK Biobank subsample 2.** Two-sided P-values shown were derived from the non-infinitesimal mixed model association test p-value as implemented in BOLT-LMM.

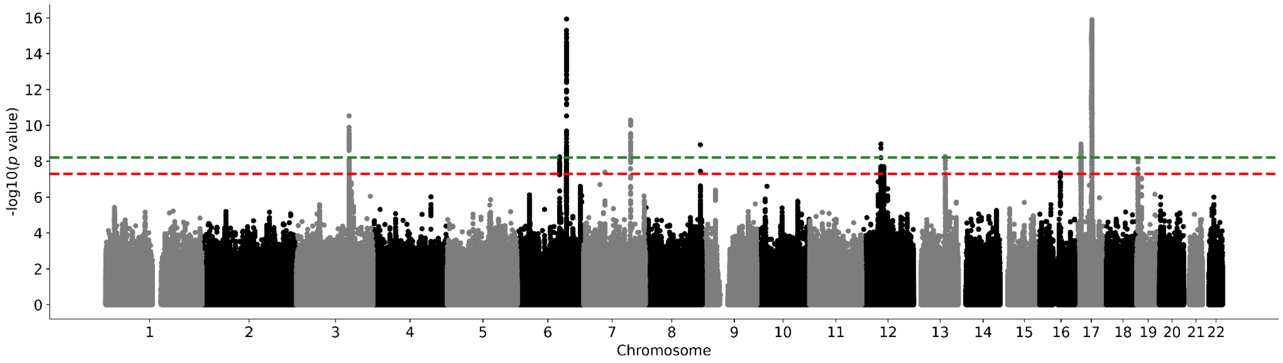

**Supplementary Figure 109. UK Biobank subsample 2 intracranial volume Manhattan plot.** Results for intracranial volume GWAS in the UK Biobank. Genome-wide significance is shown for the common threshold of $p$-value < $5x10^{-8}$ (red dashed line), and also for the multiple comparisons-corrected threshold of $p$-value < $6.25x10^{-9}$ (green dashed line). Two-sided P-values shown were derived from the non-infinitesimal mixed model association test p-value as implemented in BOLT-LMM.

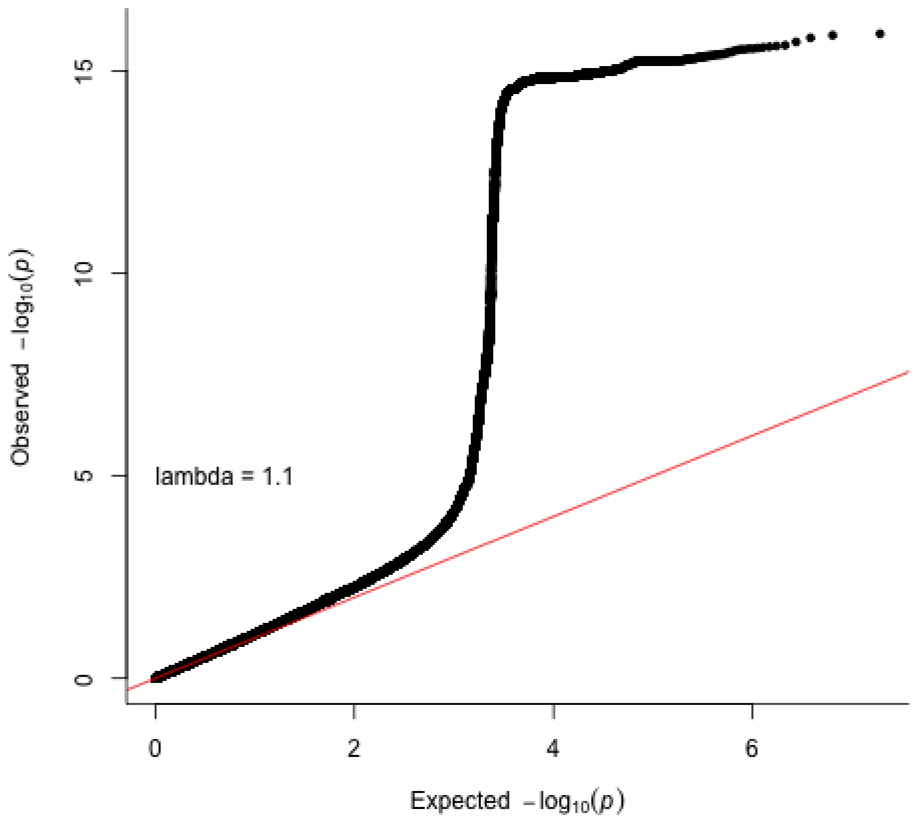

**Supplementary Figure 110. Intracranial volume QQ plot in the UK Biobank subsample 2.** Two-sided P-values shown were derived from the non-infinitesimal mixed model association test p-value as implemented in BOLT-LMM.

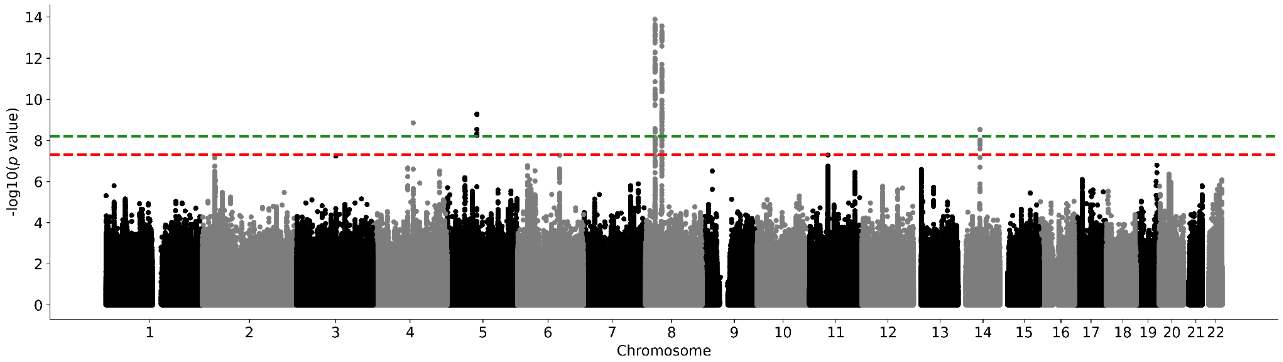

**Supplementary Figure 111. UK Biobank subsample 2 globus pallidus Manhattan plot.** Results for globus pallidus GWAS in the UK Biobank. Genome-wide significance is shown for the common threshold of $p$-value < $5x10^{-8}$ (red dashed line), and also for the multiple comparisons-corrected threshold of $p$-value < $6.25x10^{-9}$ (green dashed line). Two-sided P-values shown were derived from the non-infinitesimal mixed model association test p-value as implemented in BOLT-LMM.

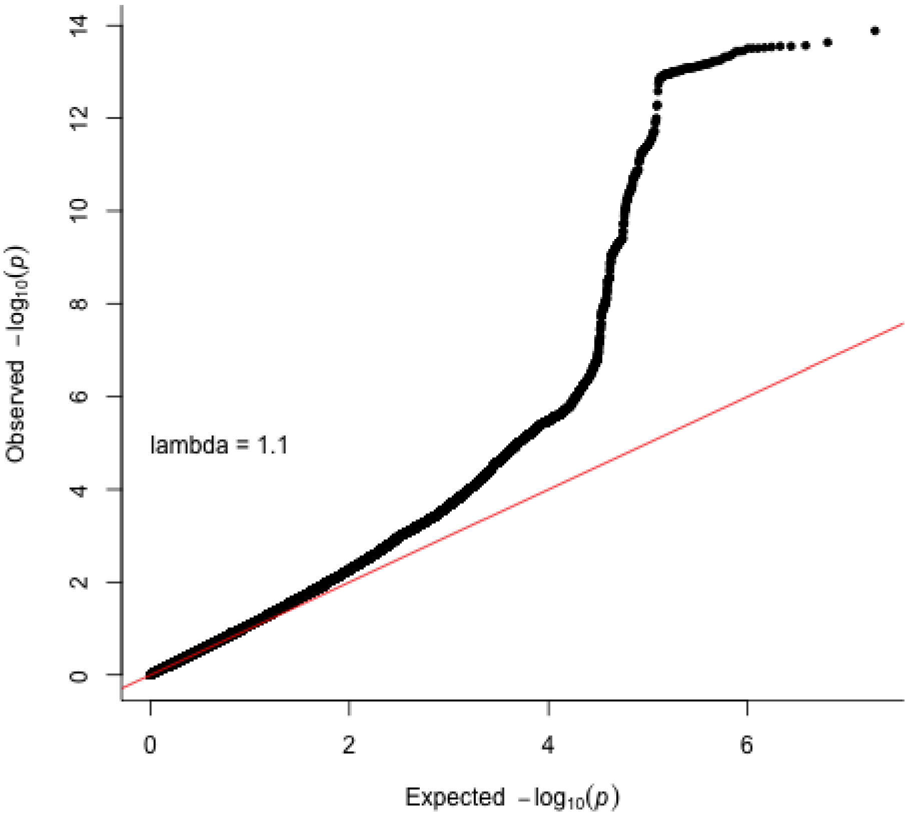

**Supplementary Figure 112. Globus pallidum QQ plot in the UK Biobank subsample 2.** Two-sided P-values shown were derived from the non-infinitesimal mixed model association test p-value as implemented in BOLT-LMM.

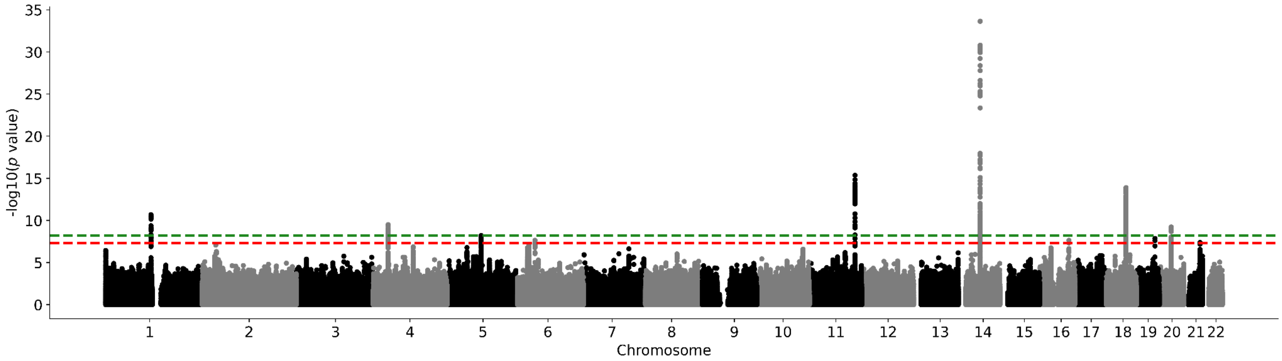

**Supplementary Figure 113. UK Biobank subsample 2 putamen Manhattan plot.** Results for putamen GWAS in the UK Biobank. Genome-wide significance is shown for the common threshold of *p*-value < 5x10$^{-8}$ (red dashed line), and also for the multiple comparisons-corrected threshold of *p*-value < 6.25x10$^{-9}$ (green dashed line). Two-sided P-values shown were derived from the non-infinitesimal mixed model association test p-value as implemented in BOLT-LMM.

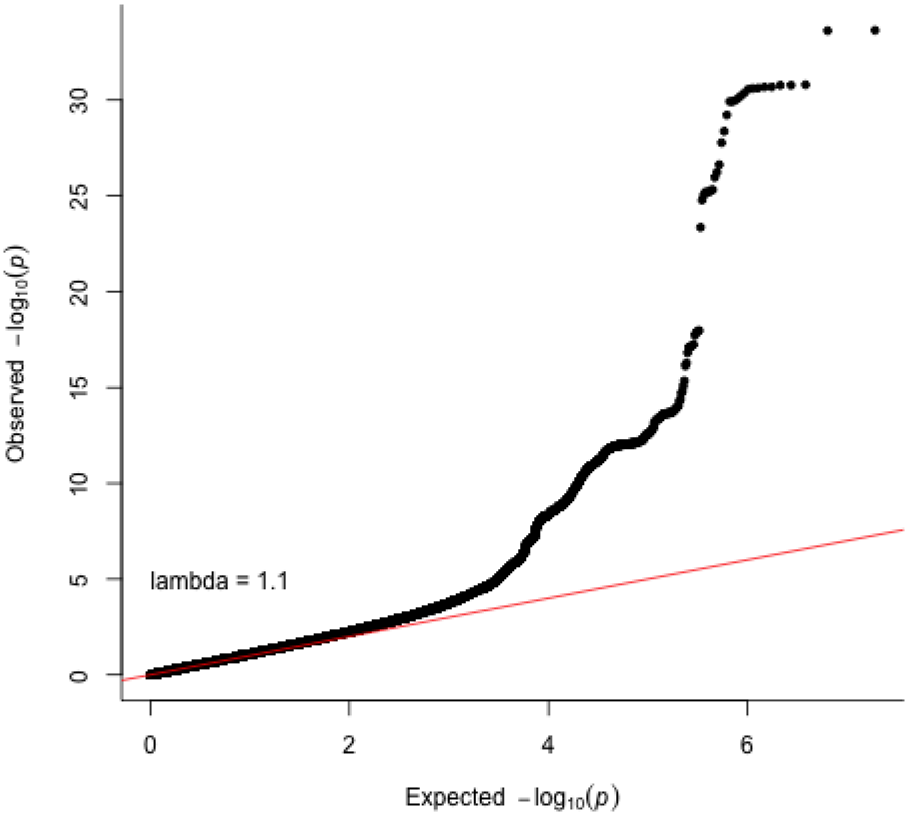

**Supplementary Figure 114. Putamen QQ plot in the UK Biobank subsample 2.** Two-sided P-values shown were derived from the non-infinitesimal mixed model association test p-value as implemented in BOLT-LMM.

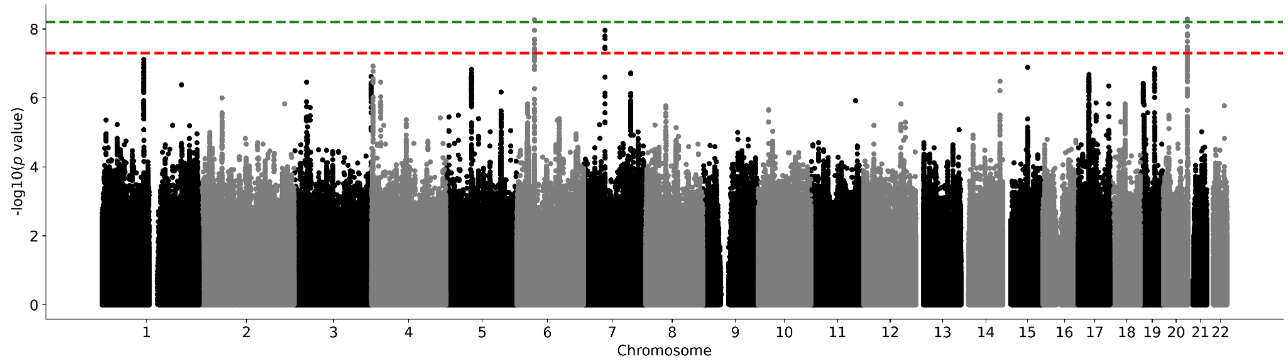

**Supplementary Figure 115. UK Biobank subsample 2 thalamus Manhattan plot.** Results for thalamus GWAS in the UK Biobank. Genome-wide significance is shown for the common threshold of *p*-value < $5x10^{-8}$ (red dashed line), and also for the multiple comparisons-corrected threshold of *p*-value < $6.25x10^{-9}$ (green dashed line). Two-sided P-values shown were derived from the non-infinitesimal mixed model association test p-value as implemented in BOLT-LMM.

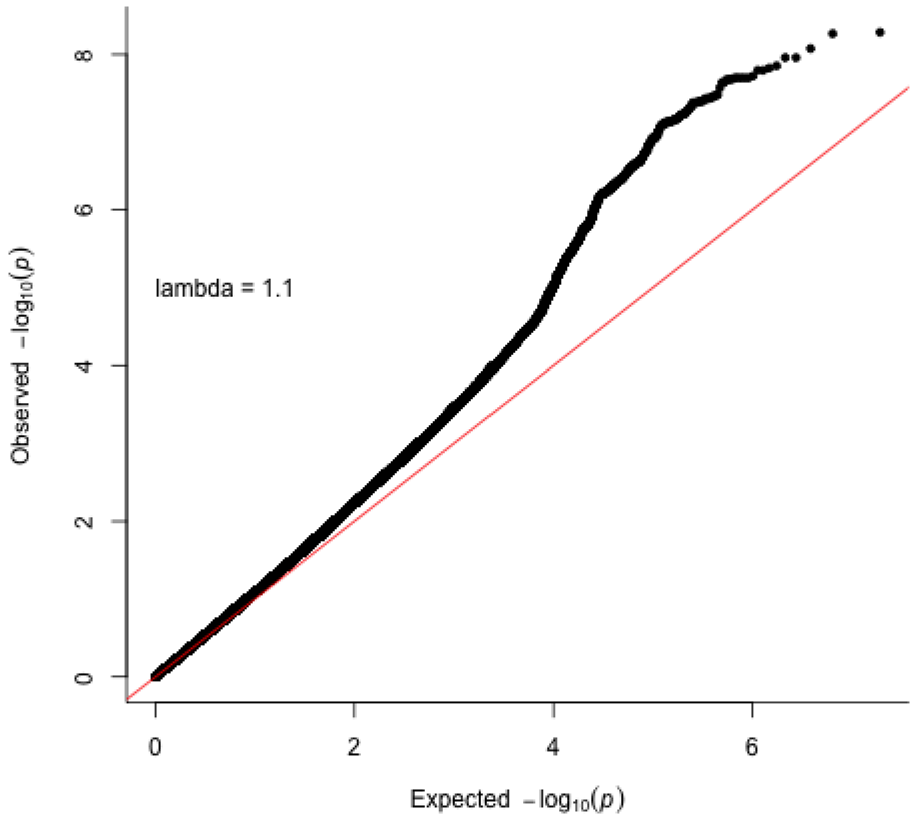

**Supplementary Figure 116. Thalamus QQ plot in the UK Biobank subsample 2.** Two-sided P-values shown were derived from the non-infinitesimal mixed model association test p-value as implemented in BOLT-LMM.

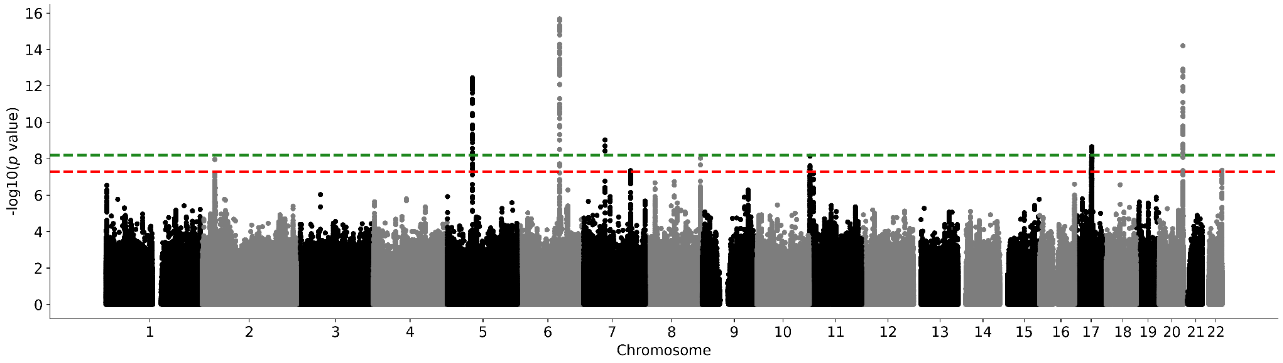

**Supplementary Figure 117. UK Biobank subsample 2 ventral diencephalon Manhattan plot.** Results for ventral diencephalon GWAS in the UK Biobank. Genome-wide significance is shown for the common threshold of *p*-value < $5x10^{-8}$ (red dashed line), and also for the multiple comparisons-corrected threshold of *p*-value < $6.25x10^{-9}$ (green dashed line). Two-sided P-values shown were derived from the non-infinitesimal mixed model association test p-value as implemented in BOLT-LMM.

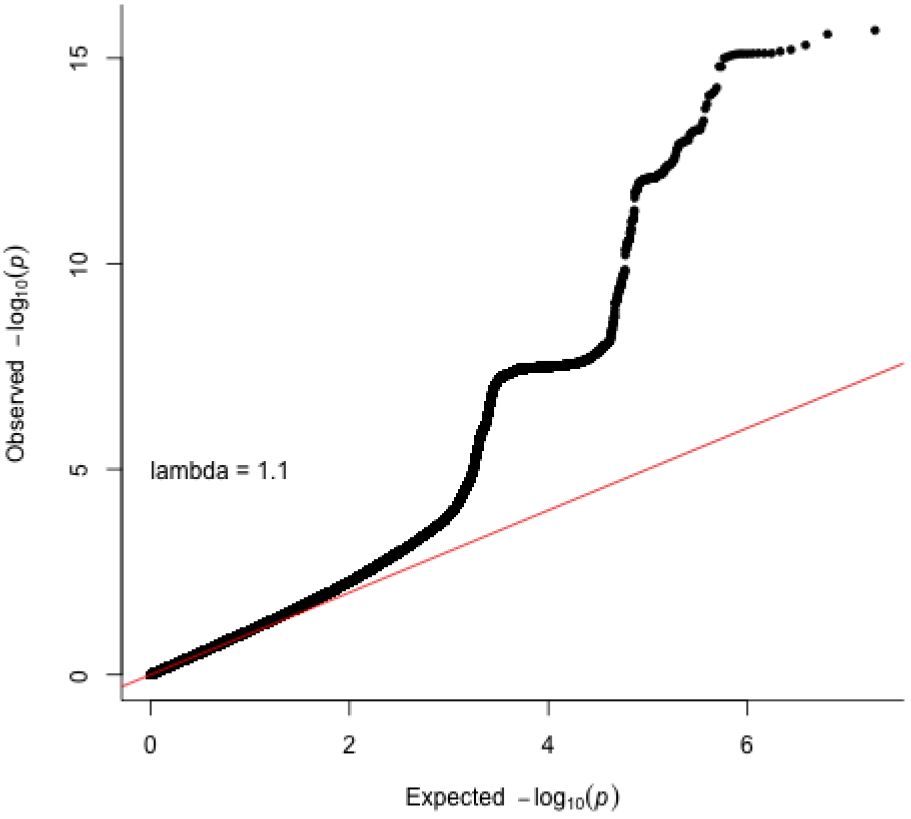

**Supplementary Figure 118. Ventral diencephalon QQ plot in the UK Biobank subsample 2.** Two-sided P-values shown were derived from the non-infinitesimal mixed model association test p-value as implemented in BOLT-LMM.

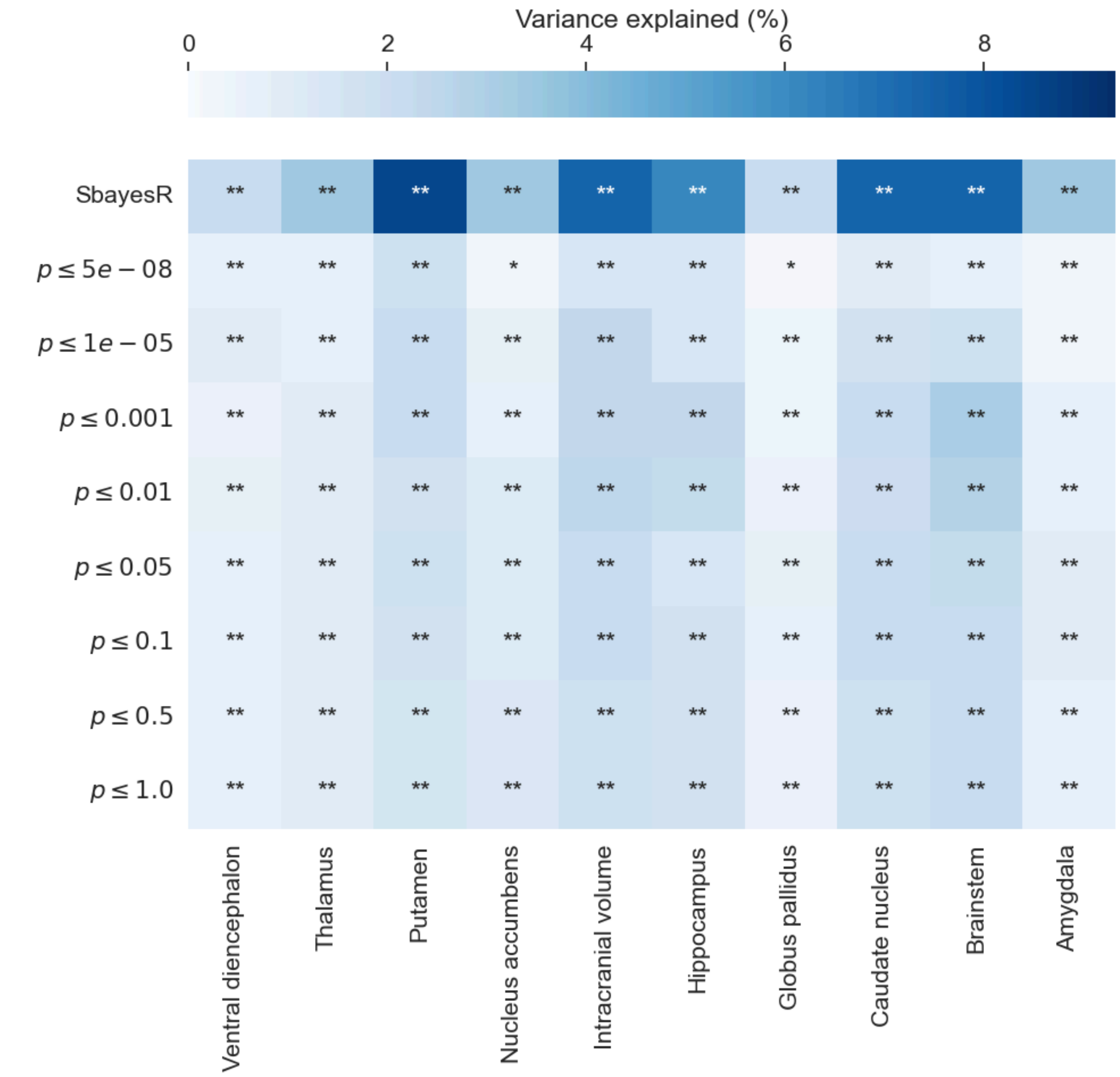

**Supplementary Figure 119.** Heatmap describing the variance explained by subcortical brain volume and intracranial volume PRS on Europeans using both SBayesR and a clumping and thresholding approach with a linear mixed effects model implemented in GCTA. P-values on the y-axis represent different thresholds for clumping and thresholding approach. Results with an asterisk (*) were nominally significant (p-value < 0.05), while those with two asterisks (**) were significant after a Bonferroni multiple testing correction [0.05 / 90 [total number of tests] = $5x10^{-5}$]. P-values in this figure correspond to two-sided wald-tests derived from the linear mixed model results

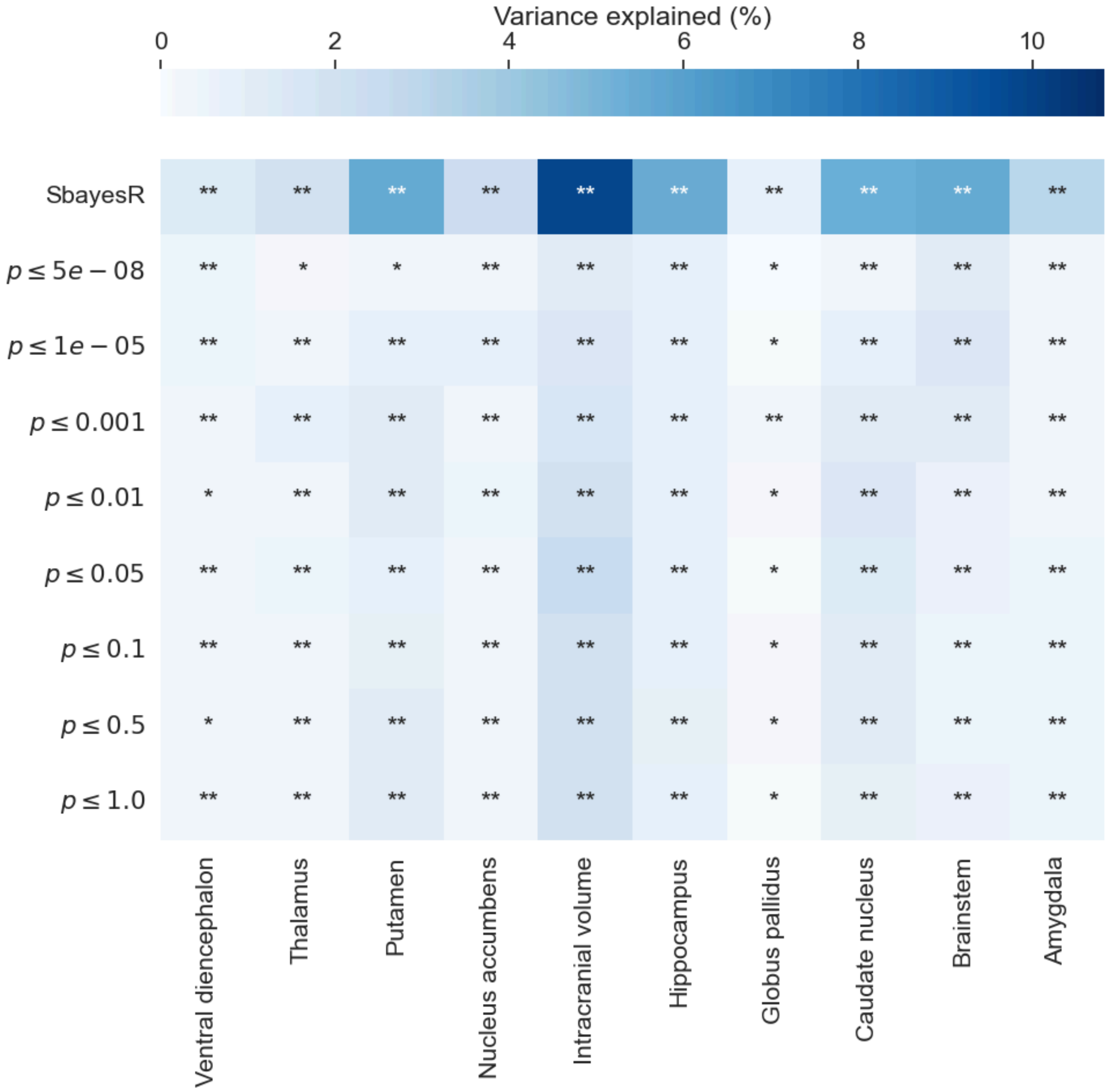

**Supplementary Figure 120.** Heatmap describing the variance explained by subcortical brain volume PRS on individuals of Non-European ancestry (African, Asian and admixed) using both SBayesR and a clumping and thresholding approach with a linear mixed effects model implemented in GCTA. P-values on the y-axis represent different thresholds for clumping and thresholding approach. Results with an asterisk (*) were nominally significant (p-value < 0.05), while those with two asterisks (**) were significant after a Bonferroni multiple testing correction [0.05 / 90 [total number of tests] = $5x10^{-5}$]. P-values in this figure correspond to two-sided wald-tests derived from the linear mixed model results

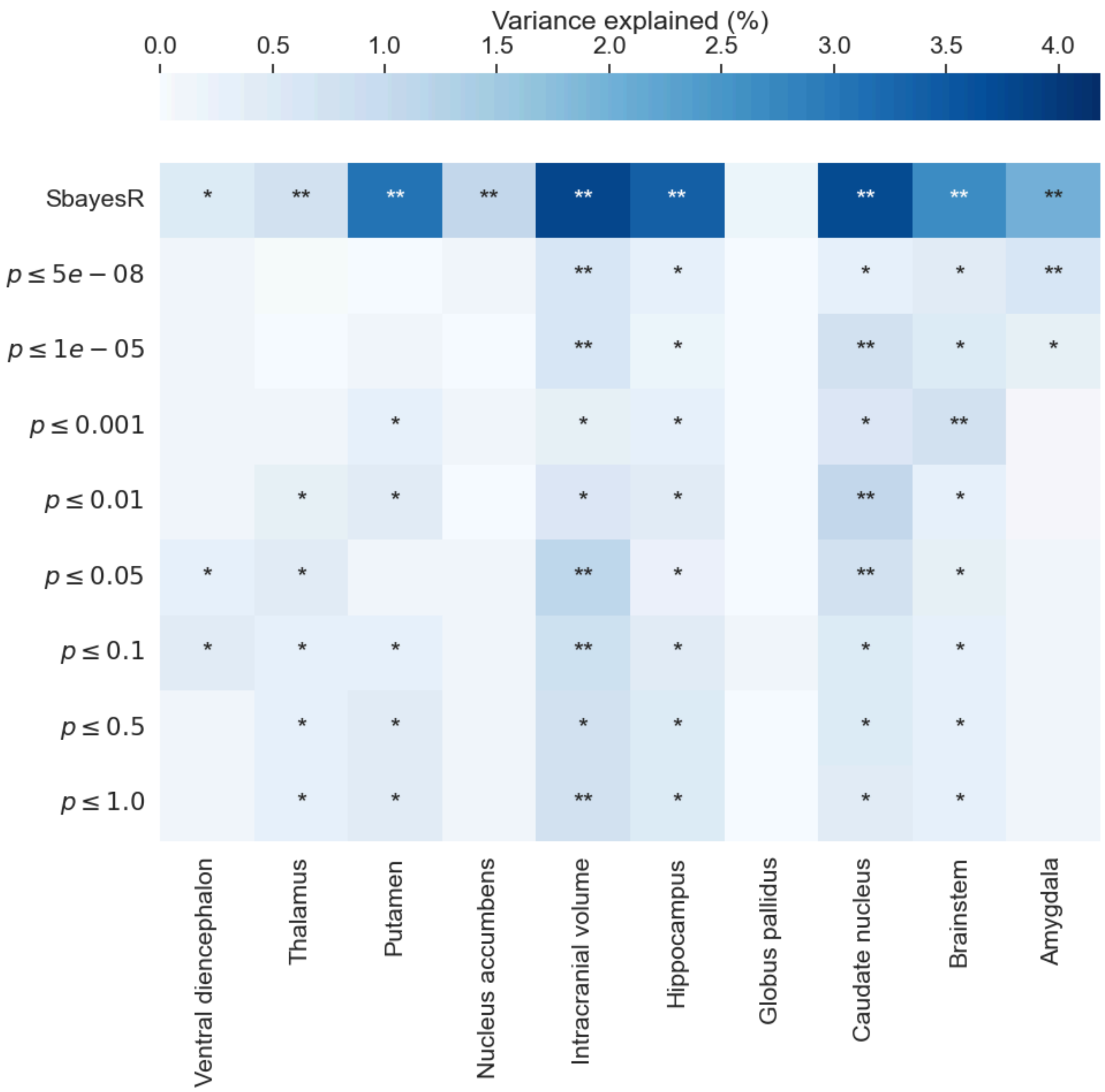

**Supplementary Figure 121.** Heatmap describing the variance explained by subcortical brain volume PRS on individuals of African ancestry using both SBayesR and a clumping and thresholding approach with a linear mixed effects model implemented in GCTA. P-values on the y-axis represent different thresholds for clumping and thresholding approach. Results with an asterisk (*) were nominally significant (p-value < 0.05), while those with two asterisks (**) were significant after a Bonferroni multiple testing correction [0.05 / 90 [total number of tests] = $5x10^{-5}$]. P-values in this figure correspond to two-sided wald-tests derived from the linear mixed model results

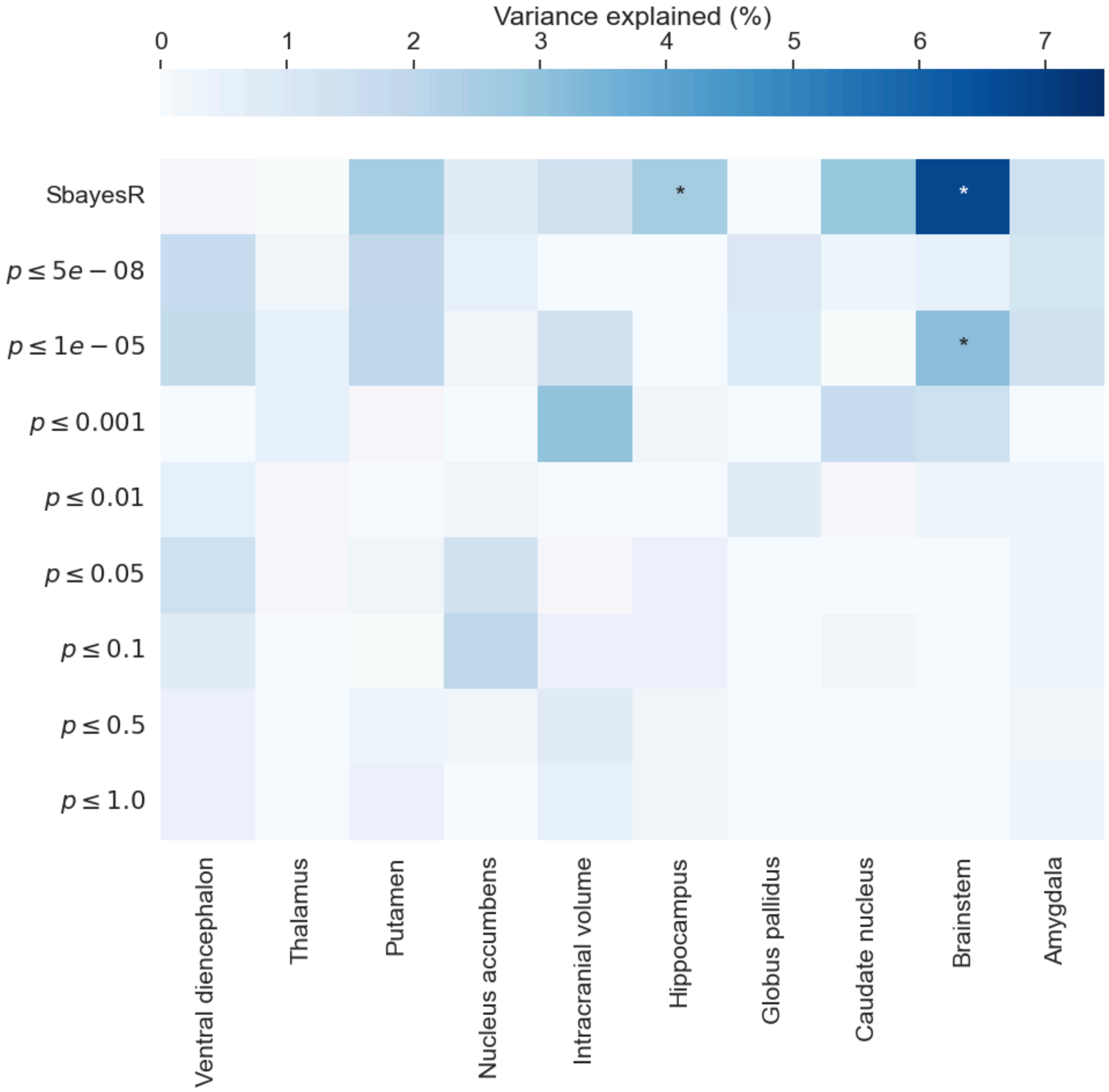

**Supplementary Figure 122.** Heatmap describing the variance explained by subcortical brain volume PRS on individuals of Asian ancestry using both SBayesR and a clumping and thresholding approach with a linear mixed effects model implemented in GCTA. P-values on the y-axis represent different thresholds for clumping and thresholding approach. Results with an asterisk (*) were nominally significant (p-value < 0.05), while those with two asterisks (**) were significant after a Bonferroni multiple testing correction [0.05 / 90 [total number of tests] = $5x10^{-5}$]. P-values in this figure correspond to two-sided wald-tests derived from the linear mixed model results

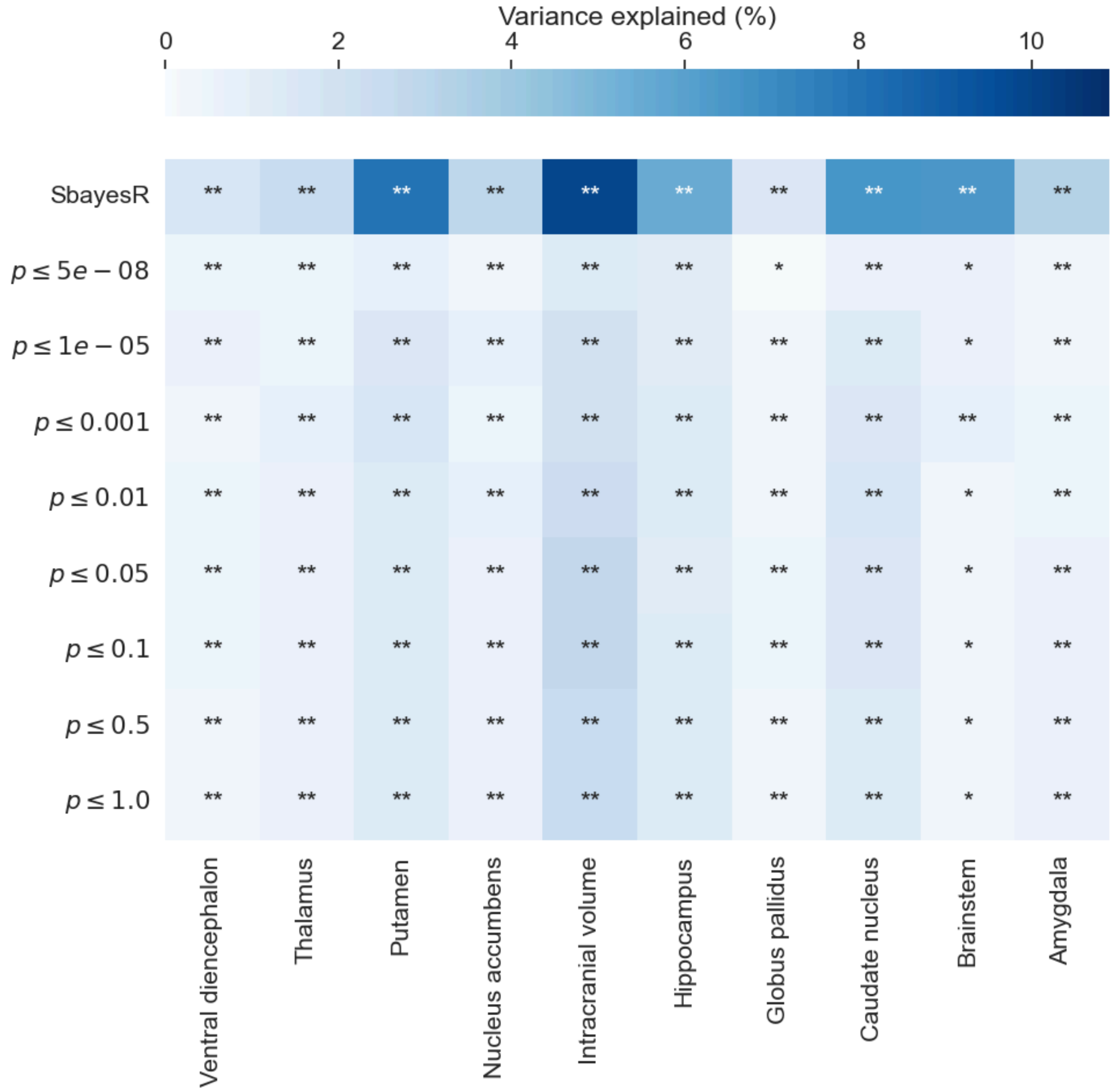

**Supplementary Figure 123.** Heatmap describing the variance explained by subcortical brain volume PRS on all ancestral samples (European, African, and Asian) using both SBayesR and a clumping and thresholding approach with a linear mixed effects model implemented in GCTA. P-values on the y-axis represent different thresholds for clumping and thresholding approach. Results with an asterisk (*) were nominally significant (p-value < 0.05), while those with two asterisks (**) were significant after a Bonferroni multiple testing correction [0.05 / 90 [total number of tests] = $5x10^{-5}$]. P-values in this figure correspond to two-sided wald-tests derived from the linear mixed model results.

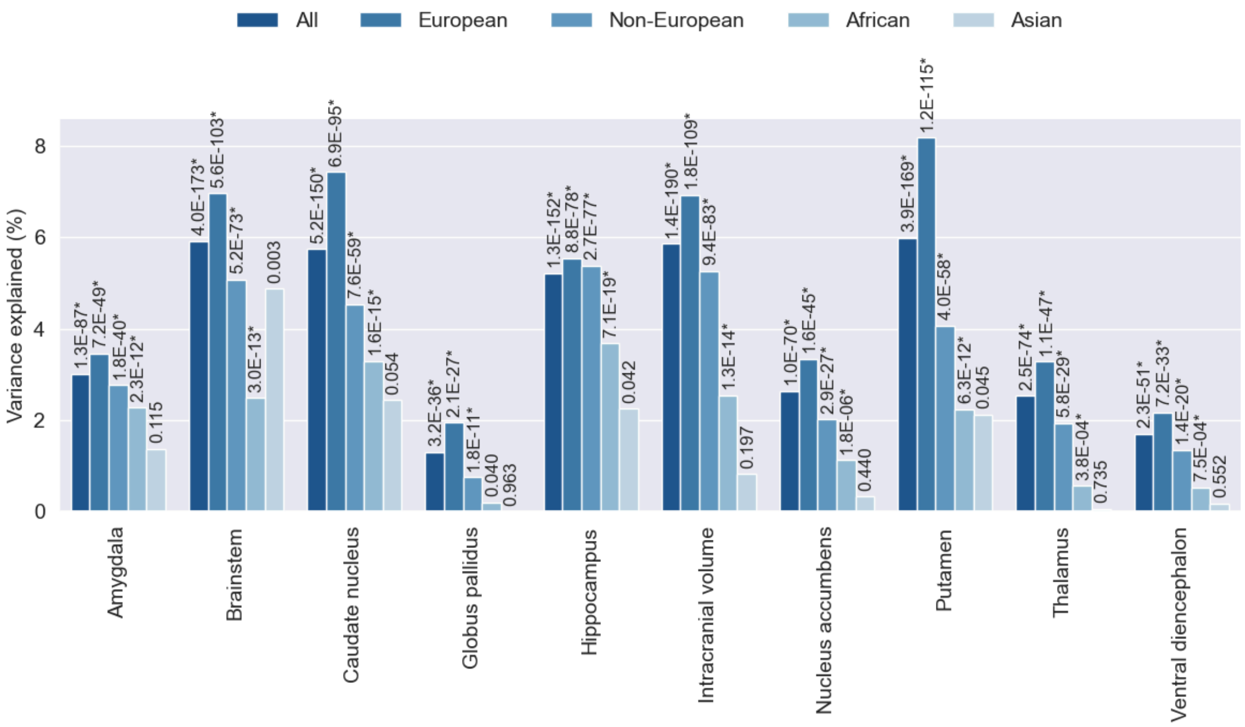

**Supplementary Figure 124. Polygenic prediction in the ABCD cohort**

Barplots describing the variance explained by intracranial and subcortical brain volume polygenic scores using the SBayesR approach with a multivariate linear regression model for the whole sample and for European, Non-European, African-only and Asian-only ancestral groups. The p-value of the association is shown at the top of each bar, those with an asterisk (*) were significant after a Bonferroni multiple testing correction [0.05 / 50 [total number of tests] = $1x10^{-3}$]. P-values in this figure correspond to two-sided wald-tests derived from the linear regression results.

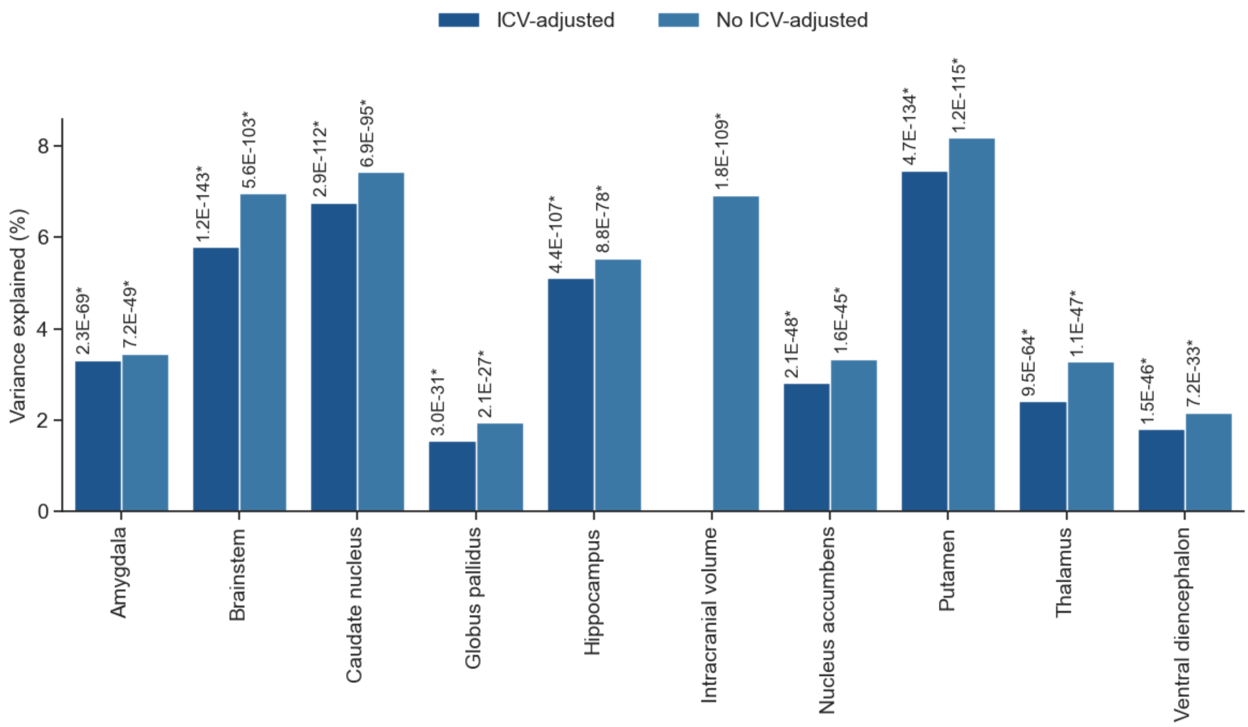

**Supplementary Figure 125. Variance explained for subcortical brain structures with and without adjustment for intracranial volume**

Barplots describing the variance explained by intracranial and subcortical brain volume polygenic scores using the SBayesR approach with a multivariate linear regression model for individuals of European ancestry only. The p-value of the association is shown at the top of each bar, those with an asterisk (*) were significant after a Bonferroni multiple testing correction [0.05 / 20 [total number of tests] = $2.5x10^{-3}$]. P-values in this figure correspond to two-sided wald-tests derived from the linear regression results.

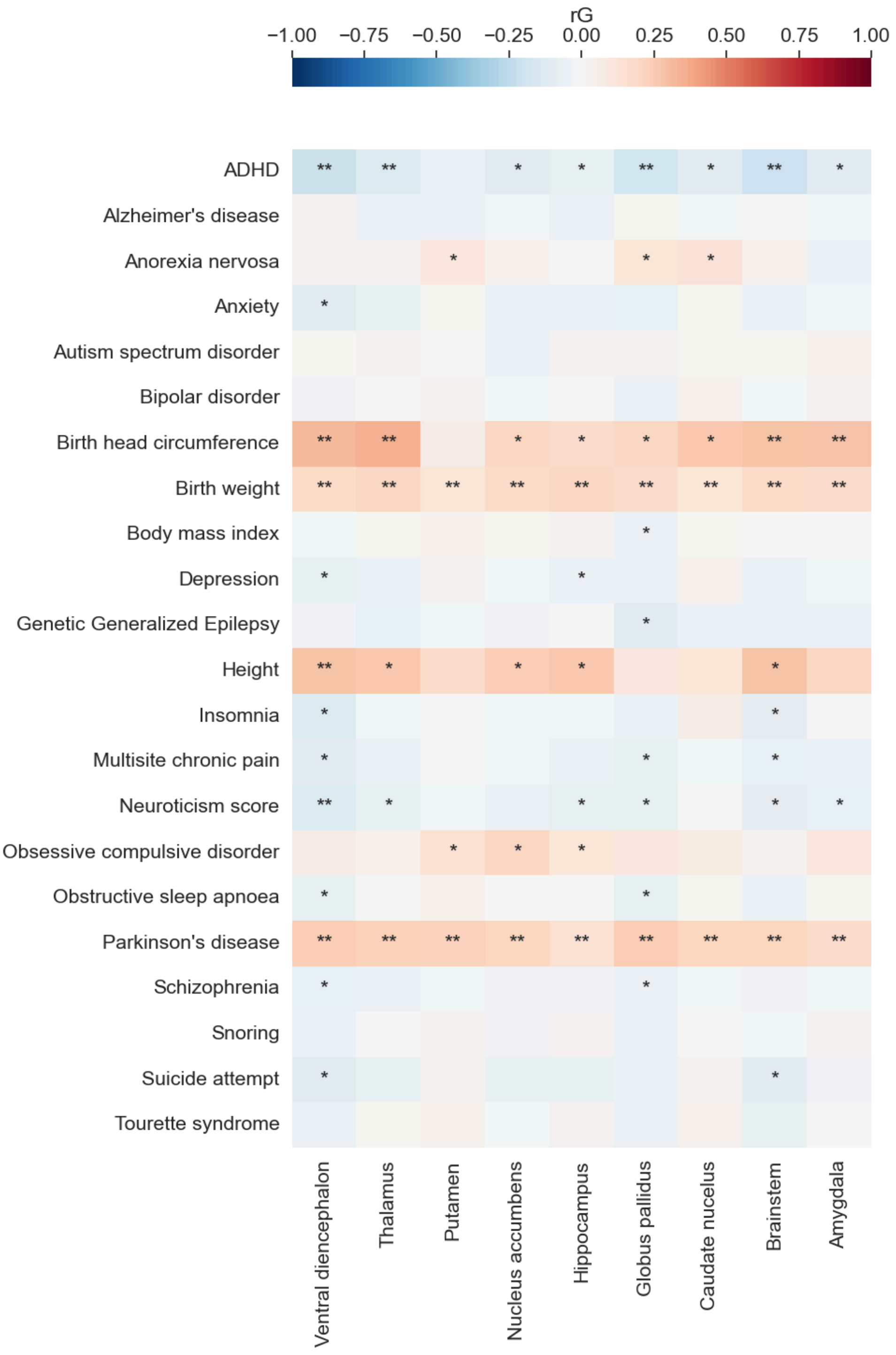

**Supplementary Figure 126. Genetic overlap with neuropsychiatric traits and disorders**

Heatmap depicting genetic correlations (rG) of subcortical brain volumes (without adjusting for ICV) from the UK Biobank with complex human phenotypes. *$p$-value < 0.05; **$p$-value significant after Bonferroni multiple testing correction (0.05 / 198 [total number of genetic correlation tests] = $2.53 \times 10^{-4}$). Genetic correlations were estimated using LD score regression. P-values correspond to chi-squared tests with one degree of freedom as implemented in LDSC regression.

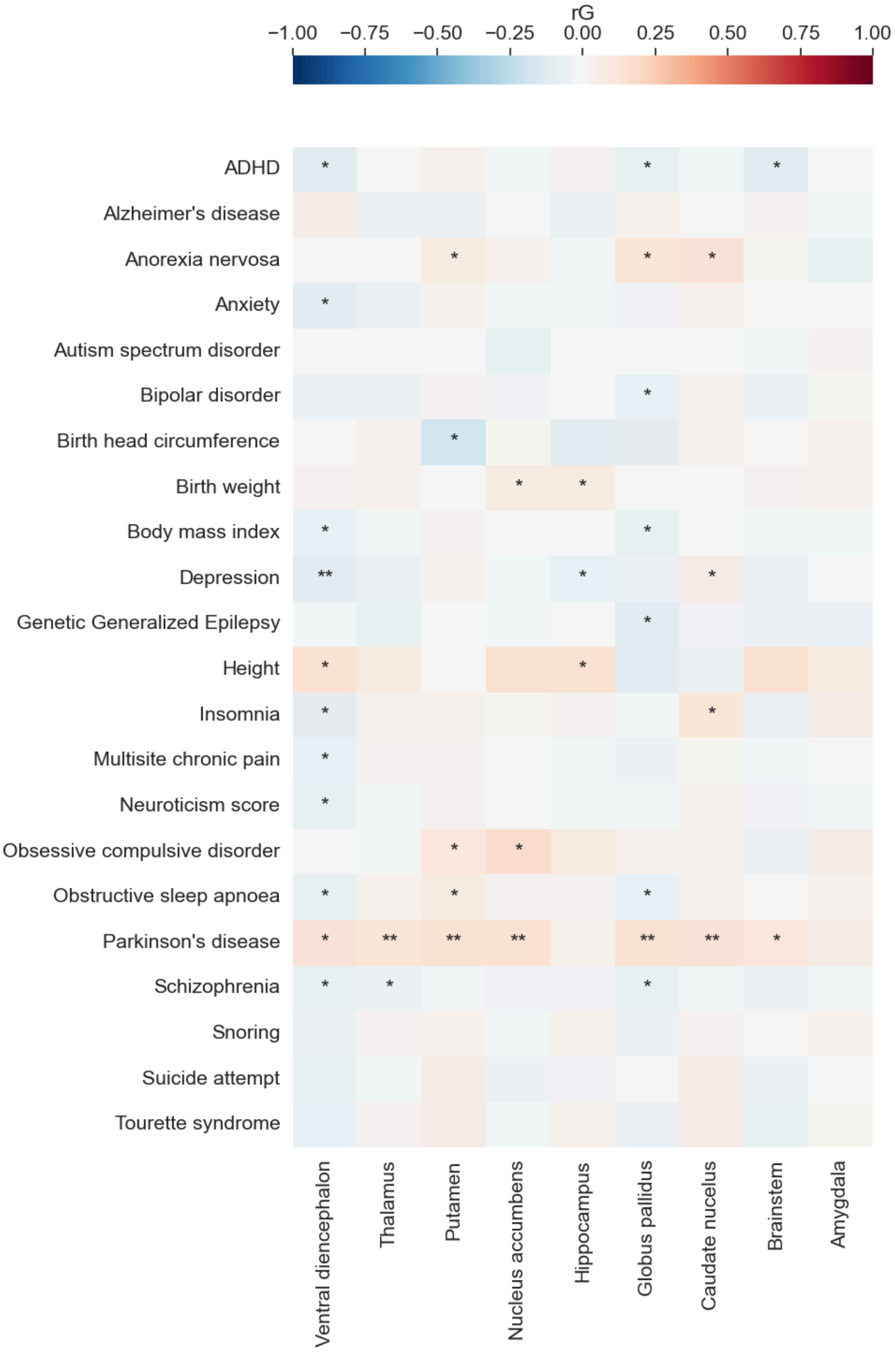

**Supplementary Figure 127. Genetic overlap with neuropsychiatric traits and disorders**

Heatmap depicting genetic correlations (rG) of subcortical brain volumes (adjusted for ICV) from the UK Biobank with complex human phenotypes. **p*-value < 0.05; ***p*-value significant after Bonferroni multiple testing correction (0.05 / 198 [total number of genetic correlation tests] = $2.53 \times 10^{-4}$). Genetic correlations were estimated using LD score regression. P-values correspond to chi-squared tests with one degree of freedom as implemented in LDSC regression.

**a)**

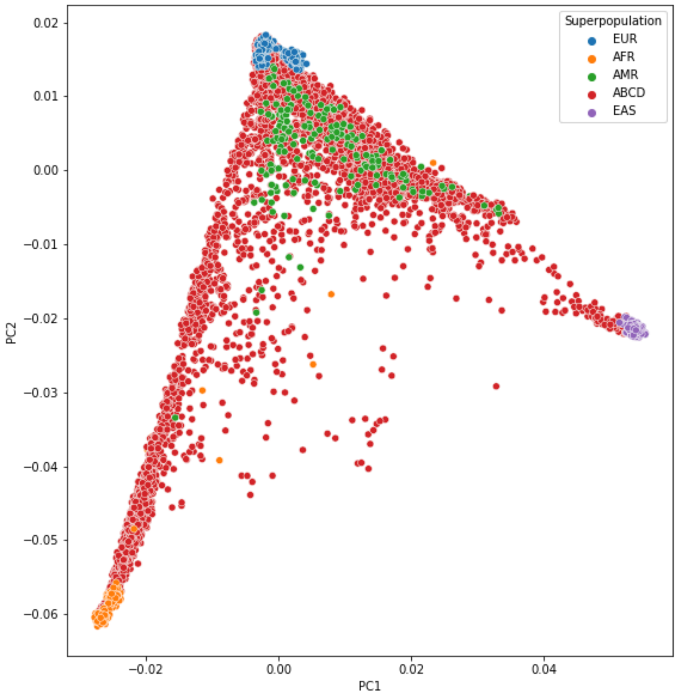

**b)**

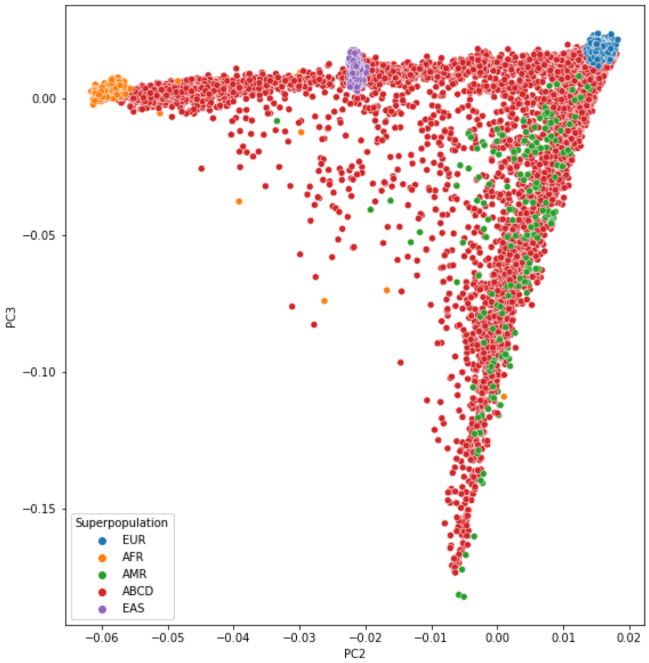

**Supplementary Figure 128. ABCD cohort ancestry principal components.** a) ABCD cohort first two principal components of the 1000Genomes project. b) ABCD cohort second and third principal components of the 1000Genomes project.

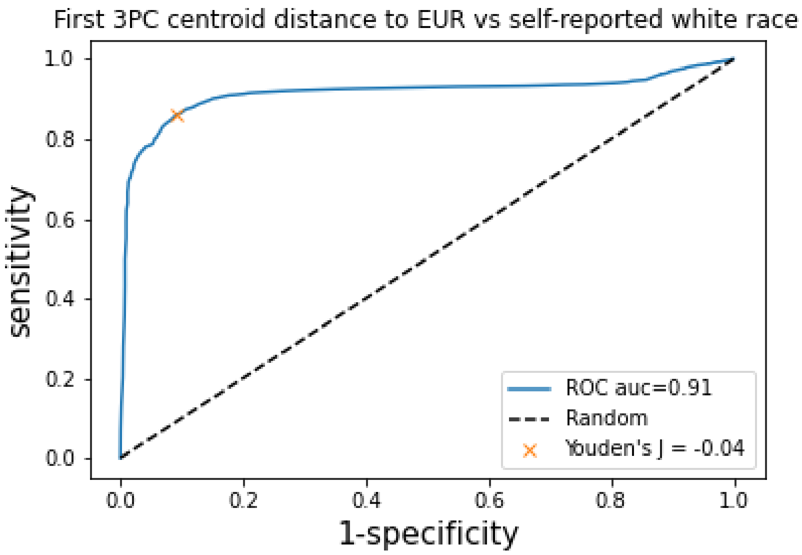

**Supplementary Figure 129. ABCD cohort ancestry classification.** Receiver operating characteristic (ROC) curve assessment of sample classification according to self-reported white race, which could be considered a proxy for European ancestry.

### Acknowledgments

This project has received funding from the European Union's Horizon 2020 Research and Innovation Programme under the Specific Grant Agreement 945539 (Human Brain Project SGA3; QTAB: National Health and Medical Research Council (NHMRC), Australia (Project Grant ID: 1078756). QTIM: NHMRC (Project Grant IDs: 486682, 1009064); This project is supported by a grant overseen by the French National Research Agency (ANR) as part of the “Investment for the Future Programme” ANR-18-RHUS-0002. It is also supported by an EU Joint Programme -Neurodegenerative Disease Research (JPND) project through the following funding organisations under the aegis of JPND -www.jpnd.eu: Australia, National Health and Medical Research Council, Austria, Federal Ministry of Science, Research and Economy; Canada, Canadian Institutes of Health Research; France, French National Research Agency; Germany, Federal Ministry of Education and Research; Netherlands, The Netherlands Organisation for Health Research and Development; United Kingdom, Medical Research Council. This project has received funding from the European Union’s Horizon 2020 research and innovation programme under grant agreement No 643417, No 640643, and No 667375 and 754517. The project also received funding from the French National Research Agency (ANR) through the VASCOGENE and SHIVA projects, and from the Initiative of Excellence of the University of Bordeaux (C-SMART project). Computations were performed on the Bordeaux Bioinformatics Center (CBiB) computer resources, University of Bordeaux. Funding support for additional computer resources has been provided to S.D. by the Fondation Claude Pompidou. Three City Study (3C-Dijon): We thank the staff and the participants of the 3C Study for their important contributions. The 3C Study is conducted under a partnership agreement between the Institut National de la Santé et de la Recherche Médicale (INSERM), the Victor Segalen–Bordeaux II University, and Sanofi-Aventis. The Fondation pour la Recherche Médicale funded the preparation and initiation of the study. The 3C Study is also supported by the Caisse Nationale Maladie des Travailleurs Salariés, Direction Générale de la Santé, Mutuelle Générale de l’Education Nationale (MGEN), Institut de la Longévité, Conseils Régionaux of Aquitaine and Bourgogne, Fondation de France, and Ministry of Research–INSERM Programme “Cohortes et collections de données biologiques.” We thank A. Boland (Centre National de Génotypage) for her technical help in preparing the DNA samples for analyses. This work was supported by the National Foundation for Alzheimer's Disease and Related Disorders, the Institut Pasteur de Lille and the Centre National de Génotypage and the LABEX (Laboratory of Excellence program investment for the future) DISTALZ - Development of Innovative Strategies for a Transdisciplinary approach to ALZheimer’s disease. Stéphanie Debette and Christophe Tzourio are recipients of grants from the French National Research Agency (ANR), a grant from the Fondation Leducq, from Joint Programme for Neurodegenerative Disease Research (JPND, BRIDGET). Stéphanie Debette is recipient of a grant from the European Research Council (ERC, SEGWAY). Marie-Gabrielle Duperron received a grant from the "Fondation Bettencourt Schueller". This work was supported by the National Foundation for Alzheimer's disease and related disorders, the Institut Pasteur de Lille, the labex DISTALZ and the Centre National de Génotypage. We thank Dr. Anne Boland (CNG) for her technical help in preparing the DNA samples for analyses.

IMAGEN was funded by the European Union-funded FP6 Integrated Project IMAGEN (LSHM-CT- 2007-037286). Further support was received from the following sources: the National Institutes of Health (NIH) (Consortium grant 5U54EB020403-05-'ENIGMA') and National Institute on Aging (NIA) 1R56AG058854-02-'ENIGMA World Aging Center'); the Medical Research Council and Medical Research Foundation ('ESTRA'- Neurobiological underpinning of eating disorders: integrative biopsychosocial longitudinal analyses in adolescents: grant MR/R00465X/; 'ESTRA' - Establishing causal relationships between biopsychosocial predictors and correlates of eating disorders and their mediation by neural pathways: grants MR/S020306/1), the Horizon 2020 funded ERC Advanced Grant 'STRATIFY' (Brain network based stratification of reinforcement-related disorders) (695313), the Medical Research Council (grant MR/W002418/1: 'Eating Disorders: Delineating illness and recovery trajectories to inform personalized prevention and early intervention in young people (EDIFY)', the National Institute for Health Research (NIHR) Biomedical Research Centre at South London and Maudsley NHS Foundation Trust and King's College London, the European Union (grant agreement no. 101057429-'environMENTAL') and Innovate UK (grant agreement no. 10038599-'environMENTAL') and the National Institute for Health and Care Research (NIHR) Maudsley Biomedical Research Centre (BRC).

The Austrian Stroke Prevention Study Family (ASPS-Fam) was funded by the Austrian Science Fund (FWF) grant number P20545-P05, P13180, PI904 the Austrian National Bank Anniversary Fund, P15435, the Austrain Federal Ministry of Science, Research and Economy under the aegis of the EU Joint Programme-Neurodegenerative Disease Research (JPND)-www.jpnd.eu and by the Austrian Science Fund P20545-B05. The Medical University of Graz supports the databank of the ASPS. The authors thank the staff and the participants for their valuable contributions. We thank Birgit Reinhart for her long-term administrative commitment, Elfi Hofer for the technical assistance at creating the DNA bank, Ing. Johann Semmler and Anita Harb for DNA sequencing and DNA analyses by TaqMan assays and

Irmgard Poelzl for supervising the quality management processes after ISO9001 at the biobankingand DNA analyses. The research reported in this article

The MCIC study was supported by the National Institutes of Health (NIH/NCRR P41RR14075 and R01EB005846 (to Vince D. Calhoun)), the Department of Energy (DE-FG02-99ER62764), the Mind Research Network, the Morphometry BIRN (1U24, RR021382A), the Function BIRN (U24RR021992-01, NIH.NCRR MO1 RR025758-01, NIMH 1RC1MH089257 to Vince D. Calhoun), the Deutsche Forschungsgemeinschaft (research fellowship to Stefan Ehrlich), and a NARSAD Young Investigator Award (to Stefan Ehrlich).

The Atherosclerosis Risk in Communities study was performed as a collaborative study supported by National Heart, Lung, and Blood Institute (NHLBI) contracts (HHSN268201100005C, HSN268201100006C, HSN268201100007C, HHSN268201100008C, HHSN268201100009C, HHSN268201100010C, HHSN268201100011C, and HHSN268201100012C), R01HL70825, R01HL087641, R01HL59367, and R01HL086694; National Human Genome Research Institute contract U01HG004402; and National Institutes of Health (NIH) contract HHSN268200625226C. Infrastructure was partly supported by grant No. UL1RR025005, a component of the NIH and NIH Roadmap for Medical Research. This project was partially supported by National Institutes of Health R01 grants HL084099 and NS087541 to MF. The authors thank the staff and participants of the ARIC study for their important contributions.

NeuroIMAGE: This project was supported by grants from National Institutes of Health (grant R01MH62873 to SV Faraone) for initial sample recruitment, and from NWO Large Investment (grant 1750102007010 to JK Buitelaar), NWO Brain & Cognition (grant 433-09-242 to JK Buitelaar), and grants from Radboud University Medical Center, University Medical Center Groningen, Accare, and VU University Amsterdam for subsequent assessment waves. NeuroIMAGE also received funding from the European Community's Seventh Framework Programme (FP7/2007 – 2013) under grant agreements n° 602805 (Aggressotype), n° 278948 (TACTICS), and n° 602450 (IMAGEMEND), and from the European Community's Horizon 2020 Programme (H2020/2014 – 2020) under grant agreements n° 643051 (MiND), n° 667302 (CoCA), and n° 728018 (Eat2beNICE).

BIG: This study used the BIG database, which was established in Nijmegen in 2007. This resource is now part of Cognomics, a joint initiative by researchers of the Donders Centre for Cognitive Neuroimaging, the Human Genetics and Cognitive Neuroscience departments of the Radboud university medical centre, and the Max Planck Institute for Psycholinguistics. The Cognomics Initiative is supported by the participating departments and centres and by

external grants, including grants from the Biobanking and Biomolecular Resources Research Infrastructure (Netherlands) (BBMRI-NL) and the Hersenstichting Nederland. In particular, the authors would also like to acknowledge grants supporting their work from the Netherlands Organization for Scientific Research (NWO), i.e. the NWO Brain & Cognition Excellence Program (grant 433-09- 229) and the Vici Innovation Program (grant 016–130-669 to BF). Additional support is received from the European Community's Seventh Framework Programme (FP7/2007 – 2013) under grant agreements nº 602805 (Aggressotype), nº 603016 (MATRICS), nº 602450 (IMAGEMEND), and nº 278948 (TACTICS), and from the European Community's Horizon 2020 Programme (H2020/2014 – 2020) under grant agreements nº 643051 (MiND) and nº 667302 (CoCA).
IMpACT: We acknowledge funding from the Netherlands Organization for Scientific Research (NWO), i.e. the Veni Innovation Program (grant 016-196-115 to MH) and the Vici Innovation Program (grant 016–130-669 to BF). The work was also supported by grant U54 EB020403 to the ENIGMA Consortium from the BD2K Initiative, a cross-NIH partnership, and by the European College of Neuropsychopharmacology (ECNP) Network "ADHD Across the Lifespan".
Munich Morphometry Sample (MPIP): The MPIP comprises images acquired as part of the Munich Antidepressant Response Signature Study and the Recurrent Unipolar Depression (RUD) Case-Control study performed at the MPIP, and control subjects acquired at the LudwigMaximilians-University, Munich, Department of Psychiatry. We thank Eva Meisenzahl and Dan Rujescu for providing MRI and genetical data for inclusion into the MPIP Munich Morphometry sample. We wish to acknowledge Anna Olynyik and radiographers Rosa Schirmer, Elke Schreiter, Reinhold Borschke and Ines Eidner for image acquisition and data preparation. We thank Dorothee P. Auer for local study management in the initial phase of the RUD study. We are grateful to GlaxoSmithKline for providing the genotypes of the Recurrent Unipolar Depression Case-Control Satizabal et al. 131 Sample. We thank the staff of the Center of Applied Genotyping (CAGT) for generating the genotypes of the MARS cohort. The study is supported by a grant of the Exzellenz-Stiftung of the Max Planck Society. This work has also been funded by the Federal Ministry of Education and Research (BMBF) in the framework of the National Genome Research Network (NGFN), FKZ 01GS0481.
SHIP is part of the Community Medicine Research net of the University of Greifswald, Germany, which is funded by the Federal Ministry of Education and Research (grants no. 01ZZ9603, 01ZZ0103, and 01ZZ0403), the Ministry of Cultural Affairs and the Social Ministry of the Federal State of Mecklenburg-West Pomerania. Genome-wide SNP typing in SHIP and MRI scans in SHIP and SHIP-TREND have been supported by a joint grant from Siemens

Healthcare, Erlangen, Germany and the Federal State of Mecklenburg-West Pomerania.
The DCHS cohort is funded by the Bill & Melinda Gates Foundation [OPP 1017641].
The Age, Gene/Environment Susceptibility-Reykjavik Study has been funded by NIH contract N01-AG-1-2100, the NIA Intramural Research Program, Hjartavernd (the Icelandic Heart Association), and the Althingi (the Icelandic Parliament). The study is approved by the Icelandic National Bioethics Committee, VSN: 00–063. The researchers are indebted to the participants for their willingness to participate in the study.
The HUNT Study is a collaboration between HUNT Research Centre (Faculty of Medicine and Movement Sciences, NTNU – Norwegian University of Science and Technology), Nord-Trøndelag County Council, Central Norway Health Authority, and the Norwegian Institute of Public Health. HUNT-MRI was funded by the Liaison Committee between the Central Norway Regional Health Authority and the Norwegian University of Science and Technology, and the Norwegian National Advisory Unit for functional MRI. Role of the Funder/Sponsor: The funding sources had no involvement in the study design, data collection, analysis, and interpretation of data; writing of the manuscript; or the decision to submit the manuscript for publication.
Cardiovascular Health Study (CHS): This CHS research was supported by NHLBI contracts HHSN268201200036C, HHSN268200800007C, HHSN268201800001C, N01HC55222, N01HC85079, N01HC85080, N01HC85081, N01HC85082, N01HC85083, N01HC85086, 75N92021D00006; and NHLBI grants U01HL080295, R01HL087652, R01HL105756, R01HL103612, R01HL120393, and U01HL130114 with additional contribution from the National Institute of Neurological Disorders and Stroke (NINDS). Additional support was provided through R01AG023629 from the National Institute on Aging (NIA). A full list of principal CHS investigators and institutions can be found at CHS-NHLBI.org. The provision of genotyping data was supported in part by the National Center for Advancing Translational Sciences, CTSI grant UL1TR001881, and the National Institute of Diabetes and Digestive and Kidney Disease Diabetes Research Center (DRC) grant DK063491 to the Southern California Diabetes Endocrinology Research Center.
Erasmus Rucphen family study (ERF): The ERF study as a part of EUROSPAN (European Special Populations Research Network) was supported by European Commission FP6 STRP grant number 018947 (LSHG-CT-2006-01947) and also received funding from the European Community's Seventh Framework Programme (FP7/2007-2013)/grant agreement HEALTH-F4-2007-201413 by the European Commission under the programme "Quality of Life and Management of the Living Resources" of 5th Framework Programme (no.

QLG2-CT-2002-01254). High-throughput analysis of the ERF data was supported by a joint grant from the Netherlands Organization for Scientific Research and the Russian Foundation for Basic Research (NWO-RFBR 047.017.043). Najaf Amin is supported by the Netherlands Brain Foundation (project number F2013(1)-28). We are grateful to all study participants and their relatives, general practitioners and neurologists for their contributions and to P. Veraart for her help in genealogy, J. Vergeer for the supervision of the laboratory work and P. Snijders for his help in data collection.

Framingham Heart Study (FHS): This work was supported by the National Heart, Lung and Blood Institute's Framingham Heart Study (Contract No. N01-HC-25195 and No. HHSN268201500001I) and its contract with Affymetrix, Inc. for genotyping services (Contract No. N02-HL-6-4278). A portion of this research utilized the Linux Cluster for Genetic Analysis (LinGA-II) funded by the Robert Dawson Evans Endowment of the Department of Medicine at Boston University School of Medicine and Boston Medical Center. This study was also supported by grants from the National Institute of Aging (R01s AG033040, AG033193, AG054076, AG049607, AG008122, AG016495; and U01-AG049505) and the National Institute of Neurological Disorders and Stroke (R01-NS017950). We would like to thank the dedication of the Framingham Study participants, as well as the Framingham Study team, especially investigators and staff from the Neurology group, for their contributions to data collection. Dr. DeCarli is supported by the Alzheimer's Disease Center (P30 AG 010129). The views expressed in this manuscript are those of the authors and do not necessarily represent the views of the National Heart, Lung, and Blood Institute; the National Institutes of Health; or the U.S. Department of Health and Human Services.

Genetic Study of Atherosclerosis Risk (GeneSTAR): is supported by grants from the National Institutes of Health National Institute of Neurological Disorders and Stroke (R01NS062059), the National Institutes of Health National Heart, Lung, and Blood Institute (U01 HL72518, HL087698) and the National Institutes of Health/National Center for Research Resources (M01-RR000052) to the Johns Hopkins General Clinical Research Center. We would like to thank the participants and families of GeneSTAR and our dedicated staff for all their sacrifices.

The Sydney Memory and Ageing Study (Sydney MAS) was funded by three National Health & Medical Research Council (NHMRC) Program Grants (ID No. ID350833, ID568969, and APP1093083).DNA samples were extracted by Genetic Repositories Australia, an Enabling Facility, which was supported by an NHMRC Grant (ID No. 401184). MRI scans were processed with the support of NHMRC Project Grants (510175 and 1025243) and an ARC Discovery Project Grant (DP0774213) and John Holden Family Foundation.

The Older Australian Twins Study (OATS) was funded by a National Health & Medical Research Council (NHMRC) and Australian Research Council (ARC) Strategic Award Grant of the Ageing Well, Ageing Productively Program (ID No. 401162); NHMRC Project (seed) Grants (ID No. 1024224 and 1025243); NHMRC Project Grants (ID No. 1045325 and 1085606); and NHMRC Program Grants (ID No. 568969 and 1093083). DNA was extracted by Genetic Repositories Australia, which was funded by the NHMRC Enabling Grant 401184. This research study was facilitated through access to Twins Research Australia, a national resource supported by a Centre of Research Excellence Grant (ID No. 1079102) from the National Health and Medical Research Council. We would like to acknowledge the contributions of the Sydney MAS and OATS research teams and thank the study participants for their time and generosity in supporting this research.

The Lothian Birth Cohort (LBC) -1936: The work was undertaken as part of the Cross Council and University of Edinburgh Centre for Cognitive Ageing and Cognitive Epidemiology (CCACE; http://www.ccace.ed.ac.uk).). This work was supported by a Research into Ageing programme grant (to I.J.D.) and the Age UK-funded Disconnected Mind project (http://www.disconnectedmind.ed.ac.uk; to I.J.D. and J.M.W.), with additional funding from the UK Medical Research Council (MRC; to I.J.D., J.M.W. and M.E.B.). The whole genome association part of this study was funded by the Biotechnology and Biological Sciences Research Council (BBSRC; Ref. BB/F019394/1). J.M.W. is supported by the Scottish Funding Council through the SINAPSE Collaboration (http://www.sinapse.ac.uk), the UK Dementia Research Institute, the Row Fogo Charitable Trust, and the Fondation Leducq. CCACE (MRC MR/K026992/1) is funded by the BBSRC and MRC. The image acquisition and analysis was performed at the Brain Research Imaging Centre, University of Edinburgh (http://www.bric.ed.ac.uk), partially funded by Row Fogo Charitable Trust (Grant no. BRO-D.FID3668413)

LIFE-Adult: LIFE-Adult is funded by the Leipzig Research Center for Civilization Diseases (LIFE). LIFE is an organizational unit affiliated to the Medical Faculty of the University of Leipzig. LIFE is funded by means of the European Union, by the European Regional Development Fund (ERDF) and by funds of the Free State of Saxony within the framework of the excellence initiative (project numbers 713-241202, 713-241202, 14505/2470, 14575/2470), and by the German Research Foundation (CRC1052 Obesity mechanisms Project A01 A. Villringer/M. Stumvoll). The authors would like to thank Matthias L. Schroeter, Leonie Lampe and Frauke Beyer for help with data acquisition and analysis and all participants and the staff at the LIFE study center.

NTR: The NTR cohort was supported by the Netherlands Organization for Scientific Research (NWO) and The Netherlands Organisation for Health Research and Development (ZonMW) grants 904-61-090, 985-10-002, 912-10-020, 904-61-193, 480-04-004,463-06-001, 451-04-034, 400-05-717, Addiction-31160008, 016-115-035, 481-08-011, 056-32-010, Middelgroot-911-09-032, OCW_NWO Gravity programme—024.001.003, NWO-Groot 480-15-001/674, Center for Medical Systems Biology (CSMB, NWO Genomics), NBIC/BioAssist/RK(2008.024), Biobanking and Biomolecular Resources Research Infrastructure (BBMRI-NL, 184.021.007 and 184.033.111); Spinozapremie (NWO-56-464-14192), KNAW Academy Professor Award (PAH/6635) and University Research Fellow grant (URF) to Dorret I. Boomsma; Amsterdam Public Health research institute (former EMGO+), Neuroscience Amsterdam research institute (former NCA); the European Science Foundation (ESF, EU/QLRT-2001-01254), the European Community's Seventh Framework Programme (FP7- HEALTH-F4-2007-2013, grant 01413: ENGAGE and grant 602768: ACTION); the European Research Council (ERC Starting 284167, ERC Consolidator 771057, ERC Advanced 230374), Rutgers University Cell and DNA Repository (NIMH U24 MH068457-06), the National Institutes of Health (NIH, R01D0042157-01A1, R01MH58799-03, MH081802, DA018673, R01 DK092127-04, Grand Opportunity grants 1RC2 MH089951 and 1RC2 MH089995); the Avera Institute for Human Genetics, Sioux Falls, South Dakota (USA). Part of the genotyping and analyses were funded by the Genetic Association Information Network (GAIN) of the Foundation for the National Institutes of Health. Computing was supported by NWO through grant 2018/EW/00408559, BiG Grid, the Dutch e-Science Grid and SURFSARA.

Religious Orders Study and Memory and Aging Project (ROSMAP): The clinical, genomic, and neuroimaging data for the Religious Orders Study and the Rush Memory and Aging Project was funded by NIH grants P30AG10161, RF1AG15819, R01AG17917, R01AG30146, R01AG40039, and the Translational Genomics Research Institute.

Rotterdam Study (RSI, RSII, RSIII): The Rotterdam Study is funded by Erasmus Medical Center and Erasmus University, Rotterdam, Netherlands Organization for the Health Research and Development (ZonMw), the Research Institute for Diseases in the Elderly (RIDE), the Ministry of Education, Culture and Science, the Ministry for Health, Welfare and Sports, the European Commission (DG XII), and the Municipality of Rotterdam. The authors are grateful to the study participants, the staff from the Rotterdam Study and the participating general practitioners and pharmacists. The generation and management of GWAS genotype data for the Rotterdam Study (RS I, RS II, RS III) were executed by the Human Genotyping Facility of the Genetic

Laboratory of the Department of Internal Medicine, Erasmus MC, Rotterdam, The Netherlands. The GWAS datasets are supported by the Netherlands Organisation of Scientific Research NWO Investments (nr. 175.010.2005.011, 911-03-012), the Genetic Laboratory of the Department of Internal Medicine, Erasmus MC, the Research Institute for Diseases in the Elderly (014-93-015; RIDE2), the Netherlands Genomics Initiative (NGI)/Netherlands Organisation for Scientific Research (NWO) Netherlands Consortium for Healthy Aging (NCHA), project nr. 050-060-810. We thank Pascal Arp, Mila Jhamai, Marijn Verkerk, Lizbeth Herrera and Marjolein Peters, and Carolina Medina-Gomez, for their help in creating the GWAS database, and Karol Estrada, Yurii Aulchenko, and Carolina Medina-Gomez, for the creation and analysis of imputed data. This work has been performed as part of the CoSTREAM project (www.costream.eu) and has received funding from the European Union's Horizon 2020 research and innovation programme under grant agreement No 667375.
SHIP is part of the Community Medicine Research Network of the University Medicine Greifswald, which is supported by the German Federal State of Mecklenburg- West Pomerania.

LMGM is supported by a UQ Research Training Scholarship from The University of Queensland (UQ); IA is supported by Research Council of Norway (grant numbers 223273, 274359), K.G. Jebsen Foundation (grant number SKGJ-MED-008). Swedish Research Council (grant numbers K2012-61X-15078-09-3, K2015-62X-15077-12-3, 2017-00949); OAA is supported by Research Council of Norway (#324499,324252,223273), NordForsk (#164218), KG Jebsen Stiftelsen (#SKGJ-MED-021), South East Norway Health Authority, NIH 1R01MH129742 - 01; KA is supported by R01AG064233, R01AG052200, UF1NS100599; AA is supported by European's Union Horizon 2020 research and innovation programme, grant agreement 728018, project Eat2beNICE; LA is supported by Research Council of Norway (grant number: 223273 and 273446); MEB is supported by UK MRC; DAB is supported by P30AG10161, RF1AG15819, R01AG17917, R01AG30146, R01AG40039, and the Translational Genomics Research Institute; JCB is supported by Infrastructure for the CHARGE Consortium is supported in part by the National Heart, Lung, and Blood Institute grant R01HL105756; MPMB is supported by R01 MH090553; HB is supported by the NHMRC, Australia; JKB has been supported by the EU-AIMS (European Autism Interventions) and AIMS-2-TRIALS programmes which receive support from Innovative Medicines Initiative Joint Undertaking Grant No. 115300 and 777394, the resources of which are composed of financial contributions from the European Union’s FP7 and Horizon2020 Programmes, and from the European Federation of Pharmaceutical Industries and Associations (EFPIA) companies’ in-kind contributions, and AUTISM SPEAKS, Autistica and SFARI; and by the Horizon2020 supported programme CANDY Grant No. 847818). The funders had no role in the design of the study; in the collection, analyses, or interpretation of data; in the writing of the manuscript, or in the decision to publish the results. Any views expressed are those of the author(s) and not necessarily those of the funders; VC is supported by NIH R01MH118695 and NSF 2112455; OTC is supported by NIH grants R01AG078533, U19AG078558, R01AG074258, R01AG077497, R01AG077000, R01AG067765, R01AG041200, R01AG062309, R01AG062200, R01AG069476; CRKC is supported by R01 MH129742-01, R56 AG058854, R01 MH116147, U54 EB020403, Baszucki Brain Research Fund and the Milken Institute's Center for Strategic Philanthropy grant; AMD is supported by U24DA041123, U24DA041147; GDS works within the MRC Integrative Epidemiology Unit at the University of Bristol, which is supported by the Medical Research Council (MC_UU_00032/01); PLDJ is supported by U01 AG061356; CDeC is supported by R01 NS17050, R01AG054076, P30 AG072972; SEh and HW is supported by German Federal Ministry of Education and Research (BMBF) grants NGFNplus MooDS (Systematic Investigation of the Molecular Causes of Major Mood Disorders and Schizophrenia) and the Integrated Network IntegraMent (Integrated Understanding of Causes and Mechanisms in Mental Disorders) under the auspices of the e:Med program (grant numbers O1ZX1314B and O1ZX1314G); SEF is supported by Max Planck Society; CF is supported by Max Planck Society (Germany); BF's contribution was supported by funding from the European Community's Horizon 2020 Programme (H2020/2014 – 2020) under grant agreement n° 847879 (PRIME). She also received relevant funding from the Netherlands Organization for Scientific Research (NWO) for the GUTS project (grant 024.005.011); KLG is supported by APP1173025; AH received support from the following sources: the Bundesministerium für Bildung und Forschung (BMBF grants 01GS08152; 01EV0711; Forschungsnetz AERIAL 01EE1406A, 01EE1406B; Forschungsnetz IMAC-Mind 01GL1745B), the Deutsche Forschungsgemeinschaft (DFG grants SM 80/7-2, SFB 940, TRR 265, NE 1383/14-1); JJH is supported by 1P30AG066546-01A1, AG062531 and an endowment from the William Castella family as William Castella Distinguished University Chair for Alzheimer's Disease Research; AJH is supported by R01MH123245; R01MH120080; NJ is supported by R01AG059874, R01MH117601; SLH is supported by Trond Mohn Research Foundation; PHL is supported by R01 MH119243, R01 MH124694, R01 GM148494, R01 MH116037; HL is supported by U01AG068221; JM is supported by grants from: - the ANR (ANR-12-SAMA-0004, AAPG2019 - GeBra), the Eranet Neuron (AF12-NEUR0008–01 - WM2NA; and ANR-18-NEUR00002–01 - ADORe), the Fondation de France (00081242), the Fondation pour la Recherche Médicale (DPA20140629802), the Mission Interministérielle de Lutte-contre-les-Drogues-et-les-Conduites-Addictives (MILDECA), the Assistance-Publique-Hôpitaux-de-Paris and INSERM (interface grant), Paris Sud University IDEX 2012, the Fondation de l'Avenir (grant AP-RM-17–013), the Fédération pour la Recherche sur le Cerveau; IM is supported by Research Council of Norway #223273/F50; BLM is supported by the Australian National Health and Medical Research Council Investigator grant scheme (APP2017176); THM is supported in part by NIH (75N92022D00004, U01HL096812, and RF1NS135615); SM is supported by Age UK, Medical Research Council and Biological Sciences Research Council; KN is supported by NLM R01 LM012535; MMN is supported by BMBF; PAN is supported by National Institute of Neurological Disorders and Stroke, R01NS062059; ZP is supported by CIHR; GP is supported by CAIP Chair in Healthy Brain Ageing, Canadian Institute for Health Research FDN-143290; MDR is supported by IntegraMent 01ZX1314G; SLR is supported by R01 AG061788; RR is supported by R.R.G was supported by EMERGIA Junta de Andalucía program (EMERGIA20_00139), Plan Propio IV, University of Seville and Plan de Generación de Conocimiento (PID2021-122853OA-I00), Ministry of Science and Innovation, Spain; GVR is supported by the ZonMw Veni grant (Veni, 1936320); CLS is supported by the National Institute on Aging (P30 AG066546, R01 AG059727, R01 AG082360), the National Institute of Neurological Disorders and Stroke (UF1 NS125513), and TARCC (2020-58-81-CR). ; AJS is supported by NIH Grants U19 AG024904, P30 AG010133, P30 AG072976, R01 AG019771; LSc is supported by NHMRC Investigator Grant 2017962, NIH RO1 MH129832; HS is supported by Austrian National Bank Anniversary Fund, P15435, City Graz, Graz, Austria and, the Austrian Ministry of Science under the aegis of the EU Joint Programme—Neurodegenerative Disease Research—www. jpnd.eu; PRS is supported by National Health and Medical Research Council; GS is supported by European Union funded Horizon Europe project 'environMENTAL' (101057429), the Horizon 2020 funded ERC Advanced Grant 'STRATIFY' (695313), the National Natural Science Foundation of China (82202093); SS is supported by grants from the NIH: P30 AG066546, U01 AG052409, RF1 AG059421, R01 AG066524, UF1 AG054076, AG058589, NS125513 and NHLBI contract 75N92019D00031-0-75920220001; LSh is supported by NIH R01 AG058854, U01 AG068057, U01 AG066833, R01 AG071470; Work from the London Cohort was supported by research grants from the Wellcome Trust (grant 084730 to S.M.S.), University College London (UCL)/University College London Hospitals (UCLH) NIHR Biomedical Research Centre/Specialist Biomedical Research Centres (CBRC/SBRC); (grant 114 to S.M.S.), the European Union Marie Curie Reintegration (to M. Matarin and S.M.S.), the UK NIHR

(08-08-SCC), the Comprehensive Local Research Network (CLRN) Flexibility and Sustainability Funding (FSF) (grant CEL1300 to S.M.S.), The Big Lottery Fund, the Wolfson Trust and the Epilepsy Society. This work was undertaken at UCLH/UCL, which received a proportion of funding from the UK Department of Health's NIHR Biomedical Research Centres funding scheme; JLS is supported by R01MH120125, R01MH118349, R56MH122819, R01MH121433; PMT and SIT are supported in part by NIH grants R01MH123163, R01MH121246, and R01MH116147. Core funding for ENIGMA was provided by the NIH Big Data to Knowledge (BD2K) program under consortium grant U54 EB020403 to PMT; DTT is supported by the Instituto de Salud Carlos III ( 00/3095, 01/3129, PI020499, PI14/00639, PI17/01056 and PI14/00918), SENY Fundació Research Grant CI2005 0308007 and Fundación Marqués de Valdecilla. Instituto de investigación sanitaria Valdecilla (A/02/07, NCT0235832 and NCT02534363); MCV is supported by Row Fogo Charitable Trust, grant no. BROD.FID3668413; DvdM is supported by the Research Council of Norway #324252 (PleioMent); JV-B has been supported by funding from ISCIII (ref.: INT22/00029) and IDIVAL (ref.: INT/A21/10 and INT/A20/04); AV is supported by the State of Saxony; JMW is supported by UK Dementia Research Institute (award no. UKDRI – Edin002, DRIEdi17/18, and MRC MC_PC_17113) which receives its funding from DRI Ltd, funded by the UK Medical Research Council, Alzheimer's Society and Alzheimer's Research UK; LBC1936 MRI brain imaging was supported by Medical Research Council (MRC) grants G0701120, G1001245, MR/M013111/1 and MR/R024065/1; Image analysis by the Row Fogo Charitable Trust; MWW is supported by U19AG024904; LTW is supported by The European Research Council under the European Union's Horizon 2020 research and Innovation program (ERC StG, Grant 802998); TW is supported by Intramural Research Program of the National Institutes of Health/; AW is supported by European Union, the European Regional Development Fund, the Free State of Saxony within the framework of the excellence initiative, the LIFE-Leipzig Research Center for Civilization Diseases, University of Leipzig [project numbers: 713-241202, 14505/2470, 14575/2470] and by grants of the German Research Foundation (DFG), contract grant numbers 209933838 CRC1052-03 A1 (AVW); JNT is supported by an NHMRC Leadership Fellowship GNT2009771, the Australian Government Department of Health and Aged Care, and the NSW Ministry of Health; JY is supported by P30AG10161, RF1AG15819, R01AG17917, R01AG30146, R01AG40039, and the Translational Genomics Research Institute; SEM is supported by grants from the Australian NHMRC APP1158127 and APP1172917. MER thanks support from Australia's National Health and Medical Research Council (GNT1102821) and the Rebecca L Cooper Medical Research Foundation (F20231230).

Data used in the preparation from the Adolescent Brain Cognitive Development (ABCD) Study (https://abcdstudy.org), held in the NIMH Data Archive (NDA). This is a multisite, longitudinal study designed to recruit more than 10 000 children age 9–10 and follow them over 10 years into early adulthood. A full list of supporters is available at https://abcdstudy.org/federal-partners.html. A listing of participating sites and a complete listing of the study investigators can be found at https://abcdstudy.org/consortium_members/. ABCD consortium investigators designed and implemented the study and/or provided data but did not necessarily participate in the analysis or writing of this report. This manuscript reflects the views of the authors and may not reflect the opinions or views of the NIH or ABCD consortium investigators. The ABCD data repository grows and changes over time. The ABCD data used in this report came from DOI:10.15154/hmjn-g821.

### Supplementary note references
